# MultiSuSiE improves multi-ancestry fine-mapping in All of Us whole-genome sequencing data

**DOI:** 10.1101/2024.05.13.24307291

**Authors:** Jordan Rossen, Huwenbo Shi, Benjamin J Strober, Martin Jinye Zhang, Masahiro Kanai, Zachary R. McCaw, Liming Liang, Omer Weissbrod, Alkes L. Price

## Abstract

Leveraging data from multiple ancestries can greatly improve fine-mapping power due to differences in linkage disequilibrium and allele frequencies. We propose MultiSuSiE, an extension of the sum of single effects model (SuSiE) to multiple ancestries that allows causal effect sizes to vary across ancestries based on a multivariate normal prior informed by empirical data. We evaluated MultiSuSiE via simulations and analyses of 14 quantitative traits leveraging whole-genome sequencing data in 47k African-ancestry and 94k European-ancestry individuals from All of Us. In simulations, MultiSuSiE applied to Afr47k+Eur47k was well-calibrated and attained higher power than SuSiE applied to Eur94k; interestingly, higher causal variant PIPs in Afr47k compared to Eur47k were entirely explained by differences in the extent of LD quantified by LD 4th moments. Compared to very recently proposed multi-ancestry fine-mapping methods, MultiSuSiE attained higher power and/or much lower computational costs, making the analysis of large-scale All of Us data feasible. In real trait analyses, MultiSuSiE applied to Afr47k+Eur94k identified 579 fine-mapped variants with PIP > 0.5, and MultiSuSiE applied to Afr47k+Eur47k identified 44% more fine-mapped variants with PIP > 0.5 than SuSiE applied to Eur94k. We validated MultiSuSiE results for real traits via functional enrichment of fine-mapped variants. We highlight several examples where MultiSuSiE implicates well-studied or biologically plausible fine-mapped variants that were not implicated by other methods.

## Introduction

Genome-wide association studies (GWAS) have provided valuable insights about human diseases and complex traits^1,2^, but statistical fine-mapping of GWAS loci often fails to identify causal variants^3,4^. Leveraging data from multiple ancestries can greatly improve fine-mapping power due to differences in linkage disequilibrium (LD), allele frequencies, and causal variant effect sizes^3,5–9^. Existing multi-ancestry fine-mapping methods that model multiple causal variants search the extremely large space of potential causal variant configurations via exhaustion^6^ or Markov Chain Monte Carlo^6,10,11^, resulting in prohibitively high running times and/or suboptimal solutions^12,13^; methods that assume a single causal variant^14,15^ are faster but suffer reduced power at loci with multiple causal variants^12,16,17^. The sum of single effects model (SuSiE) is a powerful, versatile, and fast approach to fine-mapping loci with multiple causal variants in a single ancestry^13,18^.

Here, we propose MultiSuSiE, an extension of SuSiE to multiple ancestries. Like SuSiE, MultiSuSiE efficiently and accurately searches the space of potential causal configurations using iterative Bayesian stepwise selection. Unlike SuSiE, MultiSuSiE allows causal effect sizes to vary across ancestries based on a multivariate normal prior informed by empirical data. Using All of Us^19,20^ whole-genome sequencing data in simulations and analyses of 14 quantitative traits, we compare MultiSuSiE to several existing fine-mapping methods, including two recently proposed approaches that also extend SuSiE to accommodate multiple ancestries, SuSiEx^21^ and MESuSiE^22^.

## Results

### Overview of methods

MultiSuSiE analyzes multi-ancestry genetic and phenotypic data to estimate the posterior inclusion probability (PIP, the probability of having a non-zero causal effect in at least one ancestry) and posterior mean causal effect size for each variant at a GWAS locus. MultiSuSiE extends an existing fine-mapping method, SuSiE^13,18^, to multiple ancestries. In analyses of a single ancestry, SuSiE sums across multiple single effect models, each with a single (unknown) causal variant. Each single effect model is fit and iteratively updated by sequentially residualizing the phenotype with respect to all other single effects. In analyses of multiple ancestries, MultiSuSiE likewise sums across multiple single effect models, each with a single (unknown) causal variant that is shared across ancestries, but allows causal effect sizes to vary across ancestries based on a multivariate normal prior informed by empirical data.

In detail, MultiSuSiE fits the following model:

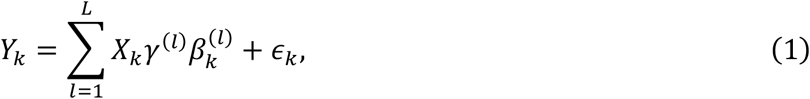

where *Y*_*k*_ is a vector of phenotypes (for ancestry *k*), *L* is the number of single effect regression models (i.e. maximum number of causal variants at the locus), *X*_*k*_ is a matrix of genotypes (for ancestry *k*), *γ*^(*l*)^ is an indicator vector with a single nonzero entry indicating which variant is causal (for single effect model *l*), 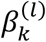 is the scalar per-allele effect size of the causal variant (for single effect model *l* in ancestry *k*), and 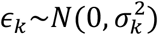 denotes residual noise with variance 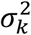(for ancestry *k*). Like SuSiE, MultiSuSiE uses iterative Bayesian stepwise regression to efficiently search the extremely large space of potential causal configurations while accounting for uncertainty in variable selection by iteratively fitting and residualizing phenotypes for the *L* single effect regression models in turn, until convergence to a stable solution. Unlike SuSiE, MultiSuSiE specifies a multivariate normal prior for 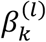 (across ancestries *k*) informed by cross-ancestry genetic correlations, under the assumption that causal variants are shared across ancestries with ancestry-specific effect sizes. MultiSuSiE outputs posterior distributions for *γ*^(*l*)^ (yielding PIPs when combined across single effect models) and 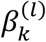 (yielding posterior mean causal effect sizes). MultiSuSiE allows for functionally informed prior distributions on *γ*^(*l*)^ (see ref.^12,23^), but incorporation of functionally informed priors is outside the scope of this study. MultiSuSiE can analyze either individual-level genetic and phenotypic data^13^ or summary association statistics and in-sample LD^18,24^. All analyses in this study use summary association statistics and in-sample LD, to minimize computational costs. We have publicly released open-source software implementing MultiSuSiE (see Code Availability).

We applied MultiSuSiE to All of Us (AoU)^19,20^ whole genome sequencing (WGS) data for 47,041 U.S. individuals of predominantly African ancestry^25^ and 94,082 U.S. individuals of European ancestry^25^ (19 million variants with MAF > 0.01 in at least one ancestry), analyzing 14 approximately independent (absolute phenotypic correlation < 0.2) quantitative traits derived from manually collected physical measurements or electronic health records (EHR) (Table 1; see Data Availability). The proportion of missing phenotypes was 23-38% higher in African ancestry individuals than in European ancestry individuals for EHR traits, likely reflecting widespread disparities in access to health care. Our analysis of WGS data avoids ancestry-based differences in imputation quality of genotyping chip data^26^, which may produce false-positive discoveries^27,28^. We have publicly released MultiSuSiE fine-mapping results and All of Us GWAS summary statistics for the 14 analyzed traits (see Data Availability).

**Table 1:** Overview of 14 quantitative traits analyzed. Missingness in Eur and Afr refer to the proportion of individuals with missing phenotypes in Eur94k and Afr47k, respectively. *N* available in Eur is different from *N* used in Eur because the sample size of Eur94k was downsampled to equal exactly twice the sample size of Afr47k for each EHR trait.

| Trait type | Trait | Missingness in Eur | Missingness in Afr | *N* in Afr | *N* available in Eur | *N* used in Eur |
| --- | --- | --- | --- | --- | --- | --- |
| Manually collected physical measurements | Resting heart rate | 1% | 1% | 47k | 93k | 93k |
|  | Height | 2% | 1% | 47k | 92k | 92k |
|  | Body mass index (BMI) | 3% | 1% | 47k | 92k | 92k |
|  | Waist-to-hip ratio, BMI adjusted | 10% | 7% | 43k | 84k | 84k |
| Electronic health record | Blood sodium concentration | 40% | 50% | 23k | 57k | 47k |
|  | Blood creatinine concentration | 41% | 52% | 23k | 56k | 45k |
|  | Blood glucose concentration | 40% | 53% | 23k | 56k | 46k |
|  | Blood potassium concentration | 41% | 54% | 22k | 56k | 43k |
|  | Mean corpuscular volume (MCV) | 40% | 54% | 22k | 57k | 43k |
|  | Platelet count | 40% | 55% | 21k | 56k | 42k |
|  | Blood total protein concentration | 43% | 57% | 20k | 54k | 40k |
|  | Alkaline phosphatase activity | 44% | 57% | 20k | 52k | 40k |
|  | Blood bilirubin concentration | 47% | 58% | 20k | 50k | 39k |
|  | Aspartate aminotransferase activity | 46% | 59% | 19k | 51k | 38k |

### Simulations

We performed simulations to evaluate fine-mapping performance across ancestries, sample sizes, and methods. We simulated 10 3Mb quantitative trait GWAS loci on chromosome 11 using empirical African-ancestry and European-ancestry AoU LD (17,262-23,634 variants per locus; Supplementary Table 1), similar to previous work^12^. In our primary simulations, each locus contained 5 randomly selected causal variants with an average per-variant *h*^2^ such that, at *N=*94k, the expected chi-squared statistic for a causal variant corresponds to a p-value of 5*10^−8^. The cross-ancestry genetic correlation of per-allele causal effect sizes was set to 0.75 (consistent with ref.^29^). Cross-ancestry differences in effect sizes were due to differences in both causal variant identity and per-allele effect sizes at shared causal variants. We simulated summary statistics directly from empirical LD using the RSS likelihood^30^, due to the high computational cost of individual-level simulations on the AoU Researcher Workbench and challenges with data egress. Further details of the simulation framework are provided in the Methods section.

First, we varied the ancestry and sample size of the input data, fine-mapping using MultiSuSiE for multi-ancestry cohorts and SuSiE for single-ancestry cohorts. We compared the empirical false-discovery rate (FDR) at different PIP thresholds (*P*(*β*_*i*,1_ = *β*_*i*,2_ = 0|*PIP*_*i*_ > α), where *β*_*i,k*_ is the true causal effect size of variant *i* in population *k*, e.g. α = 0.5 or 0.9) across five cohorts: Afr47k, Eur47k, Eur94k, Afr23k+Eur23k, and Afr47k+Eur47k (Figure 1a and Supplementary Table 2). To assess calibration, we compared the empirical FDR to (1 – PIP threshold), a conservative FDR upper bound (as in ref. ^12^), as well as (1 – mean PIP), the expected FDR (which has been reported to be slightly mis-calibrated in previous fine-mapping simulations^12,31^). All cohorts were well-calibrated with respect to the conservative FDR upper bound, but slightly mis-calibrated with respect to the expected FDR (also see Supplementary Figure 1), consistent with previous reports of imperfect calibration of SuSiE PIPs^12,31^. We generally observed slightly larger FDR for cohorts with lower sample size per ancestry, particularly Afr23k+Eur23k; we observed similarly greater empirical FDR when performing fine-mapping in Afr23k or Eur23k cohorts (Supplementary Figure 2), implying that greater empirical FDR are a consequence of sample size, and not specific to MultiSuSiE.

**Figure 1:**
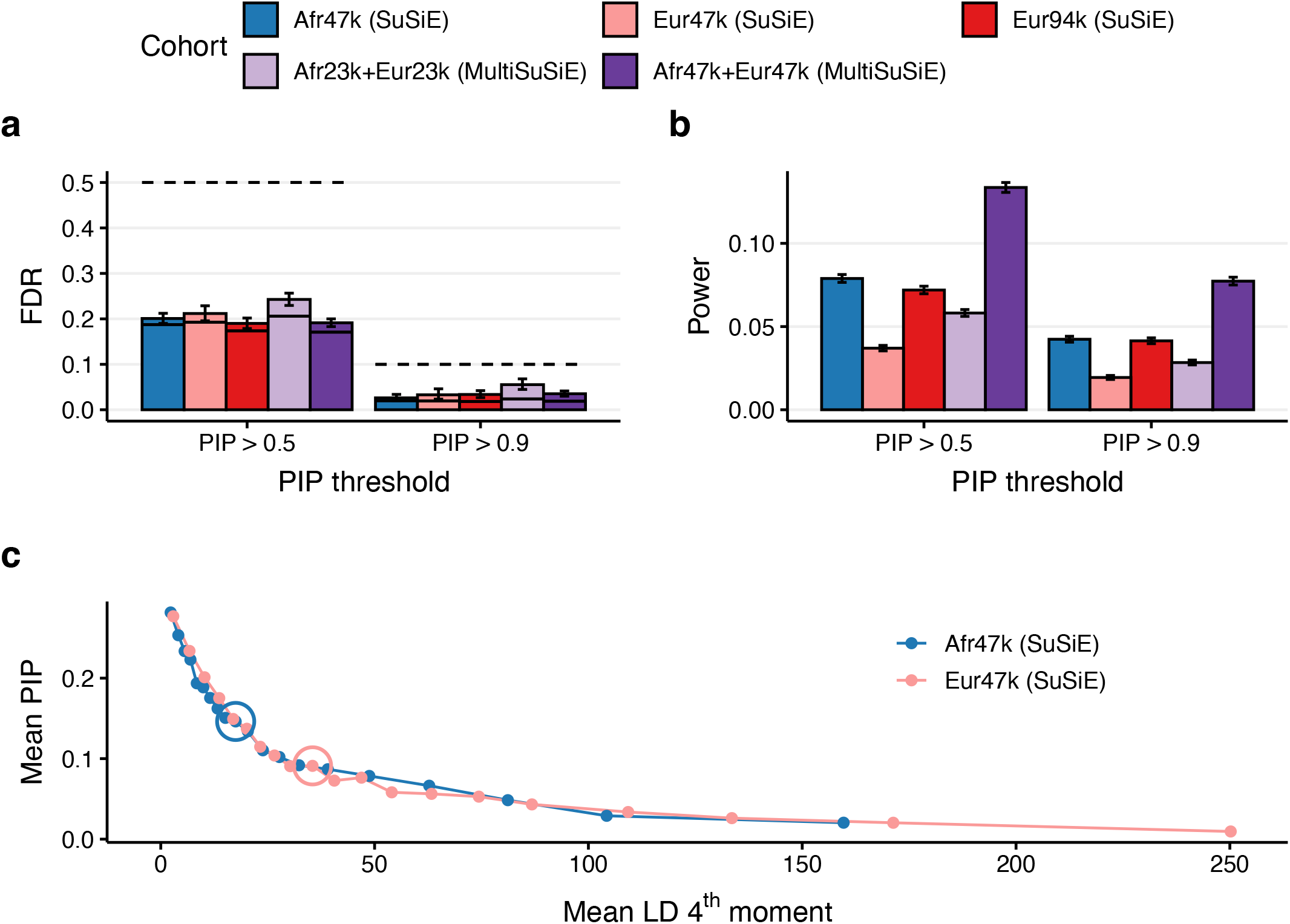
Simulation results across ancestries and sample sizes analyzed. We report **(a)** the FDR (bars), conservative FDR upper bound (dashed line), and expected FDR (solid bars) at PIP > 0.5 and PIP > 0.9, **(b)** power at PIP > 0.5 and PIP > 0.9, **(c)** mean PIP of causal variants with MAF > .05 within 20 equally sized LD 4^th^ moment bins in Afr47k and Eur47k fine-mapping. Error bars denote 95% confidence intervals. Circled dots denote the 10^th^ LD 4^th^ moment bin for each cohort. Numerical results are reported in Supplementary Tables 2-3.

We also compared the power at different PIP thresholds (*P*(*PIP*_*i*_ > α|*β*_*i*,1_ ≠ 0 *or β*_*i*,2_ ≠ 0), e.g. α = 0.5 or 0.9) across the 5 cohorts (Figure 1b and Supplementary Table 2). Multi-ancestry fine-mapping outperformed European-ancestry fine-mapping at matched sample sizes (e.g. 91% improvement at *N*=94k and 57% improvement at *N*=47k for PIP > 0.5). The improvement in power for Afr47k+Eur47k vs. Eur94k was greater at variants with higher African MAF than European MAF (Supplementary Figure 3) and larger African effect sizes (Supplementary Figure 4). Surprisingly, single-ancestry fine-mapping using Afr47k outperformed multi-ancestry fine-mapping using Afr23k+Eur23k, (36% improvement for PIP > 0.5), in contrast to previously reported simulations^5^. Fine-mapping with Afr47k additionally outperformed single-ancestry fine-mapping using Eur47k (e.g. 113% improvement for PIP > 0.5) and Eur94k (9.6% improvement for PIP > 0.5).

The larger PIPs for simulated causal variants when fine-mapping in African-ancestry samples can be attributed to lower levels of LD, which can be quantified by causal variant LD 4^th^ moments. For Afr47k and Eur47k, for each simulated common causal variant with in-sample MAF > 0.05, we calculated the in-sample LD 4^th^ moment^32^ (defined for variant *i* as 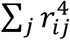, where *j* indexes all other variants at the locus and *r*_*ij*_ is the correlation between variants *i* and *j*). We constructed 20 equally sized bins based on LD 4^th^ moments and determined that, conditional on the mean LD 4^th^ moment, the mean PIPs when fine-mapping in Afr47k or Eur47k were roughly equal (Figure 1c and Supplementary Table 3). Regressing causal variant PIPs on population labels and (log-transformed) LD 4^th^ moments confirmed that the impact of ancestry on fine-mapping PIPs is entirely explained by LD 4^th^ moments (Supplementary Table 4), such that the smaller values of LD 4^th^ moments in Afr47k compared to Eur47k (Supplementary Figure 5) explain the higher fine-mapping power in Afr47k. On the other hand, LD scores (LD 2nd moments), MAF, and true causal effect sizes failed to explain the impact of ancestry on causal variant PIPs (Supplementary Figures 6-8, Supplementary Table 4); compared to LD scores, LD 4^th^ moments heavily weight correlations close to 1, which pose particular challenges for fine-mapping.

Next, we compared the performance of 6 multi-ancestry fine-mapping methods: MultiSuSiE; SuSiEx, an unpublished multi-ancestry fine-mapping method that also extends the SuSiE model to multiple ancestries, but assumes that the effects of causal variants across ancestries are uncorrelated and applies several filters to the estimated single effect regressions^21^; MESuSiE, a recently published multi-ancestry fine-mapping method that also extends the SuSiE model to multiple ancestries, but does not assume that causal variants are shared across ancestries^22^; MCVmeta, defined as the application of SuSiE (allowing for multiple causal variants per locus) to a fixed-effect meta-analysis across ancestries (using meta-analyzed LD)^33,34^; SCVmeta, defined as the application of SuSiE (allowing only a single causal variant per locus) to a fixed-effect meta-analysis across ancestries (which does not require LD)^15,35,36^; and PIPmeta, a post-hoc approach for combining PIPs from SuSiE applied to each ancestry separately 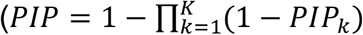, where *PIP*_*k*_ is estimated using SuSiE in ancestry *k*). All methods were run on Afr47k+Eur47k. A list of methods with dedicated software packages is provided in Table 2. We did not include two previously published multi-ancestry fine-mapping methods, PAINTOR^6,23,37^ and MGfm^11^, in our comparisons due to prohibitively high running times for loci with tens of thousands of variants (Table 2); we note that SuSiEx^21^ and MESuSiE^22^ have previously been reported to outperform PAINTOR in simulations with fewer variants.

**Table 2:** Overview of multi-ancestry fine-mapping software packages. Each row describes a multi-ancestry fine-mapping method with a dedicated software package. Running times and memory requirements refer to analyses of a single 3Mb window, averaged across 100 simulations for each of 10 3Mb windows using the main simulation settings, except for PAINTOR and MGfm which were applied to a single simulation and locus. MGfm did not complete in 48 hours (2880min).

|  | Individual-level data input | Summary statistics LD input | Supports functional priors | Software type | Reference | Average running time (min) | Max memory required (GB) |
| --- | --- | --- | --- | --- | --- | --- | --- |
| MultiSuSiE | X | X | X | Python | This work | 2.2 | 5.0 |
| SuSiEx |  | X |  | Command-line | <sup>21</sup> | 6.0 | 4.3 |
| MESuSiE |  | X | X | R | <sup>22</sup> | 41 | 71 |
| PAINTOR |  | X | X | Command-line | <sup>6,23,37</sup> | 314 | 30 |
| MGfm |  | X |  | R | <sup>11</sup> | did not complete | 68 |

We compared the empirical false-discovery rate at different PIP thresholds of the 6 methods (Figure 2a and Supplementary Table 5). All methods were well-calibrated with respect to the conservative FDR upper bound, except for MCVmeta (FDR = 0.13 for PIP > 0.9). All methods were miscalibrated with respect to expected FDR (MultiSuSiE, SuSiEx and MESuSiE were only slightly miscalibrated), consistent with previous reports^12,31^.

**Figure 2:**
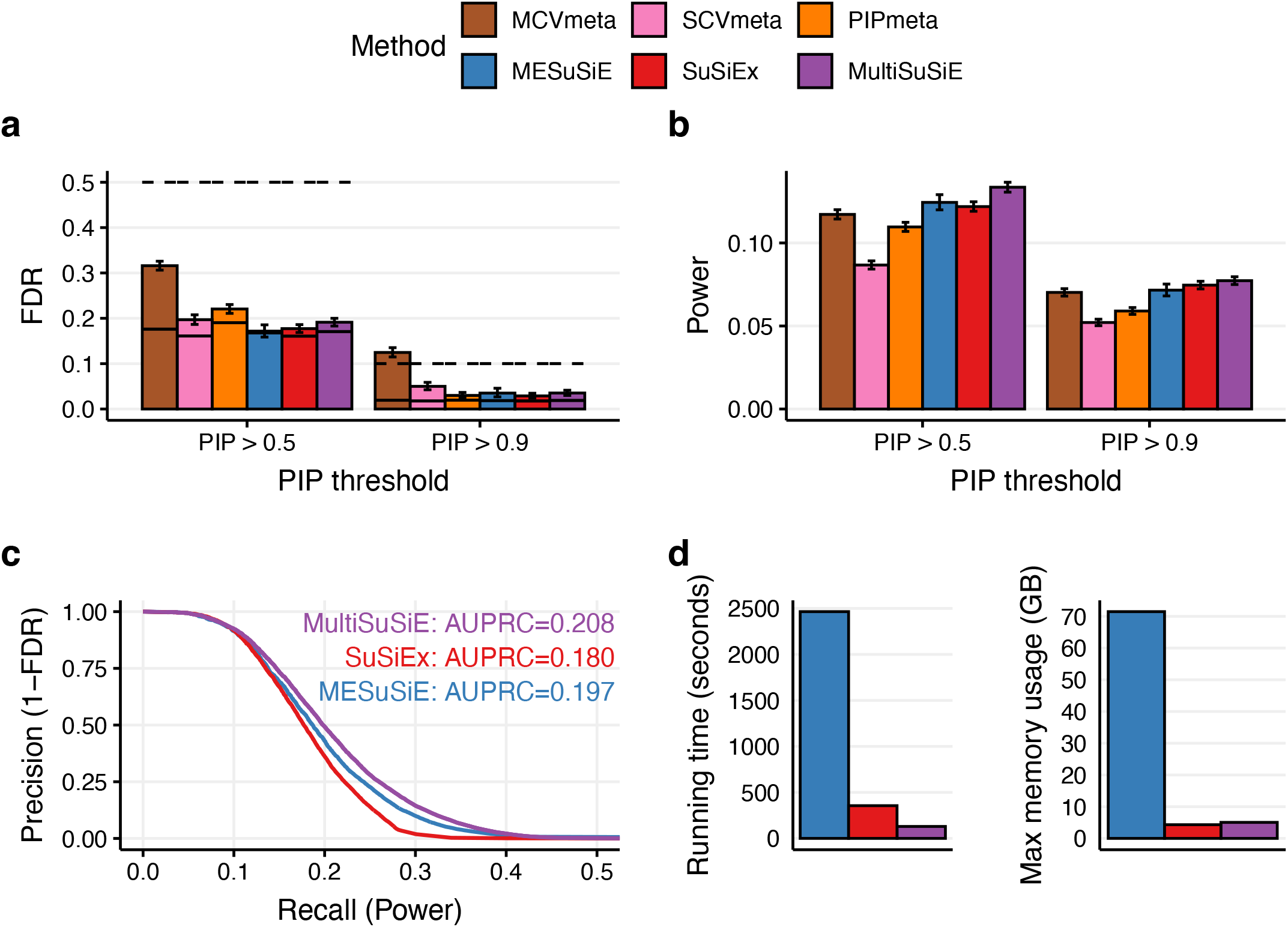
Simulation results for Afr47k+Eur47k across fine-mapping methods. We report **(a)** the FDR (bars), conservative FDR upper bound (dashed line), and expected FDR (solid bars) at PIP > 0.5 and PIP > 0.9, **(b)** power at PIP > 0.5 and PIP > 0.9, **(c)** precision-recall curves varying PIP threshold for methods with dedicated software packages, **(d)** Running times and memory requirements for methods with dedicated software packages. Error bars denote 95% confidence intervals. Numerical results are reported in Supplementary Tables 5-6.

We also compared the power at different PIP thresholds of the 6 methods (Figure 2b and Supplementary Table 5). MultiSuSiE attained 8-54% higher power than the 5 other methods at PIP > 0.5, and 4-48% higher power at PIP > 0.9 (Figure 2b and Supplementary Table 5). To compare the overall performance of the three methods with dedicated software packages (MultiSuSiE, SuSiEx, and MESuSiE), we calculated precision and recall across PIP thresholds (Figure 2c and Supplementary Table 6) and found that MultiSuSiE attained 16% and 6% higher area under the precision recall curve (AUPRC) than SuSiEx and MESuSiE, respectively. This was driven by higher power of MultiSuSiE at moderate FDR; the three methods attained similar power at low FDR. Finally, we compared the running time and memory usage of these three methods (Figure 2d and Table 2). MultiSuSiE was 2.7x faster than SuSiEx (with 1.2x higher memory usage) and 19x faster than MESuSiE (with 14x lower memory usage).

We ran SuSiEx at non-default parameters settings to better understand the difference in performance between MultiSuSiE and SuSiEx. We were able to improve the performance of SuSiEx by disabling two filters that remove single effect regression models with 95% credible sets that either (i) contain pairs of variants with absolute correlation less than 0.5 (purity filter^13^) or (ii) do not contain a variant with marginal GWAS p-value less than 10^−5^ (*p*-value filter). We refer to this method as SuSiEx-unfiltered. We determined that SuSiEx-unfiltered attained nearly identical power and FDR as MultiSuSiE, but was 6.4x slower (with 1.2x lower memory usage) (Supplementary Figure 9).

We also ran MESuSiE restricting to variants with MAF > 0.001 in both populations, as in ref.^22^. We refer to this method as MESuSiE-intersection. MESuSiE-intersection had 1.3x lower power and 1.7x higher FDR at PIP > 0.5 than MultiSuSiE due to causal variants that did not pass the MAF filter (Supplementary Figure 9).

We performed 9 secondary analyses; we excluded MESuSiE from all secondary analyses (except as noted) due to its high running time and memory requirements. First, we compared fine-mapping performance across ancestries, sample sizes, and methods under alternative genetic architectures with increased or decreased mean heritability per variant (Supplementary Figures 10-13). We consistently observed lower FDR and higher power at higher heritability and higher FDR and lower power at lower heritability. We reached similar conclusions as in our main simulations, with four exceptions: (i) Afr23k+Eur23k failed to satisfy the conservative FDR upper bound at low heritability, suggesting that particular care must be taken when fine-mapping weak GWAS signals with small sample sizes; (ii) fine-mapping with Afr23k+Eur23k attained lower power than Eur47k at low heritability (power=0.0063 vs 0.0041; p=2.19*10^−7^ for difference), potentially due to the larger number of parameters estimated by MultiSuSiE; (iii) LD 4^th^ moments did not fully explain the impact of ancestry on causal variant PIPs at low heritability; and (iv) SCVmeta performed nearly as well as MultiSuSiE at low heritability (but substantially worse at high heritability). Second, we considered alternative generative models with the same cross-ancestry genetic correlation of effect sizes, but where cross-ancestry differences in causal effect sizes were either (i) entirely due to differences in causal variant identity, or (ii) entirely due to differences in per-allele effect sizes at shared causal variants; we included MESuSiE in comparisons under the first alternative generative model, because it does not assume that all causal variants are shared across ancestries. For both (i) and (ii), we reached similar conclusions as in our main simulations (Supplementary Figures 14-17). Third, we restricted our simulations to a subset of variants with similar MAF distributions in Afr47k and Eur47k, resulting in similar mean *h*^2^ across ancestries. We reached similar conclusions as in our main simulations (Supplementary Figures 18-19), with one exception: Eur94k attained 29% higher power than Afr47k, compared to 9% lower power in our main simulations. Fourth, we varied the cross-ancestry genetic correlation hyperparameter (*ρ*) used by MultiSuSiE, and observed very little difference in performance (Supplementary Figure 20). This is consistent with the observation that MultiSuSiE and SuSiEx-unfiltered performed similarly (Supplementary Figure 9), because SuSiEx-unfiltered is similar to MultiSuSiE with *ρ* set to 0. Fifth, we fine-mapped Afr23k+Eur23k using reference LD matrices from Afr47k+Eur47k, which is a superset of Afr23k+Eur23k, to mimic the effect of excluding samples with missing phenotypes while computing summary statistics but not while computing in-sample reference LD matrices. We confirmed that FDR and power were not impacted by this change (Supplementary Figure 21-22). Sixth, we performed simulations using UK Biobank (UKBB) genotypes (see Data Availability) (*N*=7K European samples and *N*=7K African samples) to assess the effects of long-range admixture LD^38^ on FDR. We selected causal variants throughout chromosome 11 based on allele frequency differences between Africans and Europeans (as larger allele frequency differences generate more admixture LD^39^), simulated individual-level phenotypes to ensure accurate modeling of long-range admixture LD, and fine-mapped 3Mb windows with MultiSuSiE using in-sample LD and PC1-adjusted summary statistics. We used UKBB to avoid the high cost of individual-level simulations and challenges with data egress on the AoU Researcher Workbench. Including PC1 as a covariate was sufficient to control FDR in all but the most extreme simulations, where all causal variants had absolute allele frequency difference between Africans and Europeans greater than 0.3 (Supplementary Figure 23). Seventh, we performed additional simulations using UK Biobank genotypes (*N*=7K African samples and *N*=7K European samples) to assess robustness to population stratification (environmental effect based on PC1, reflecting African vs. European ancestry); we confirmed that including PC1 as a covariate while computing summary association statistics (but not while computing in-sample LD) was sufficient to avert population stratification effects (Supplementary Figure 24). Eighth, we ran MCVmeta and SCVmeta on Afr47k+Eur47k restricting to variants with MAF > 0.01 in both populations (instead of just one population). These methods suffered decreased power and increased FDR due to the effects of causal variants that did not pass the MAF filter (Supplementary Figure 25); this is consistent with the poor performance of MESuSiE-intersection, which applies a similar MAF filter (Supplementary Figure 9). Ninth, we compared the performance of PIPmeta to alternative methods for combining PIPs across cohorts. Combining PIPs using the maximum PIP^22,40^ performed comparably to PIPmeta, but combining PIPs using the minimum PIP^22^ or mean PIP decreased power substantially (Supplementary Figure 26).

### Multi-ancestry fine-mapping of 14 quantitative traits in the All of Us Research Program

We performed multi-ancestry fine-mapping of 14 quantitative traits in AoU (Table 1; WGS data with sample sizes up to European *N*=94k, African *N*=47k), varying ancestry, sample size and method. We analyzed 6 cohorts: Afr47k, Eur47k, Eur94k, Afr23k+Eur23k, Afr47k+47k (as in simulations) and Afr47k+Eur94k. We performed GWAS using Plink2^41^ and fine-mapped 1,758 unique overlapping 3Mb windows^12^ (2,862 window-trait pairs, Supplementary Table 7) that contained a variant in the central 1Mb with *p* < 5*10^−6^ for at least one of Afr47k, Eur94k, or Afr47k+Eur47k (using inverse variance based fixed-effect meta-analysis^42^), excluding three regions with long-range LD^43^ and restricting to variants with MAF > 0.01 in Afr47k or Eur47k and missingness < 0.05 in both Afr47k and Eur47k (2,313-48,867 variants per window, Supplementary Table 7). We chose to fine-map the same set of loci across cohorts to focus comparisons on differences in fine-mapping performance, rather than differences in GWAS discovery power. To account for differences in trait missingness rates across ancestries for EHR-based traits (Table 1), we subselected individuals of European ancestry to match missingness in individuals of African ancestry. We excluded MESuSiE from real-trait analyses due to its high running time and memory requirements (Figure 2d), and excluded MCVmeta due to its miscalibrated FDR in simulations (Figure 2a). We have publicly released GWAS summary statistics for all variants (censored as required by AoU) and fine-mapping results for all variants with PIP > 0.01 (see Data Availability).

First, we varied the ancestry and sample size of the input data, fine-mapping using MultiSuSiE for multi-ancestry cohorts and SuSiE for single-ancestry cohorts (Figure 3 and Supplementary Table 8). Across the 14 traits, MultiSuSiE applied to Afr47k+Eur94k identified 579 variants with PIP > 0.5 and 197 variants with PIP > 0.9. At total *N*=94k, MultiSuSiE applied to Afr47k+Eur47k identified 44% more variants at PIP > 0.5 than SuSiE applied to Eur94k (*p*=1*10^−6^ for difference; 44% more variants at PIP > 0.9, *p*=0.01 for difference). At total *N*=47k, MultiSuSiE applied to Afr23k+Eur23k identified 26% more variants at PIP > 0.5 than SuSiE applied to Eur47k (*p*=0.04 for difference) and 19% fewer variants than SuSiE applied to Afr47k (*p*=0.02 for difference). We observed similar relative results (with fewer fine-mapped variants in total) when restricting a given cohort to window-trait pairs that contained a variant in the central 1Mb with *p* < 5*10^−6^ (resp. *p* < 5*10^−8^) in that cohort (instead of at least one of Afr47k, Eur94k, or Afr47k+Eur94k) (Supplementary Figure 27-28).

**Figure 3:**
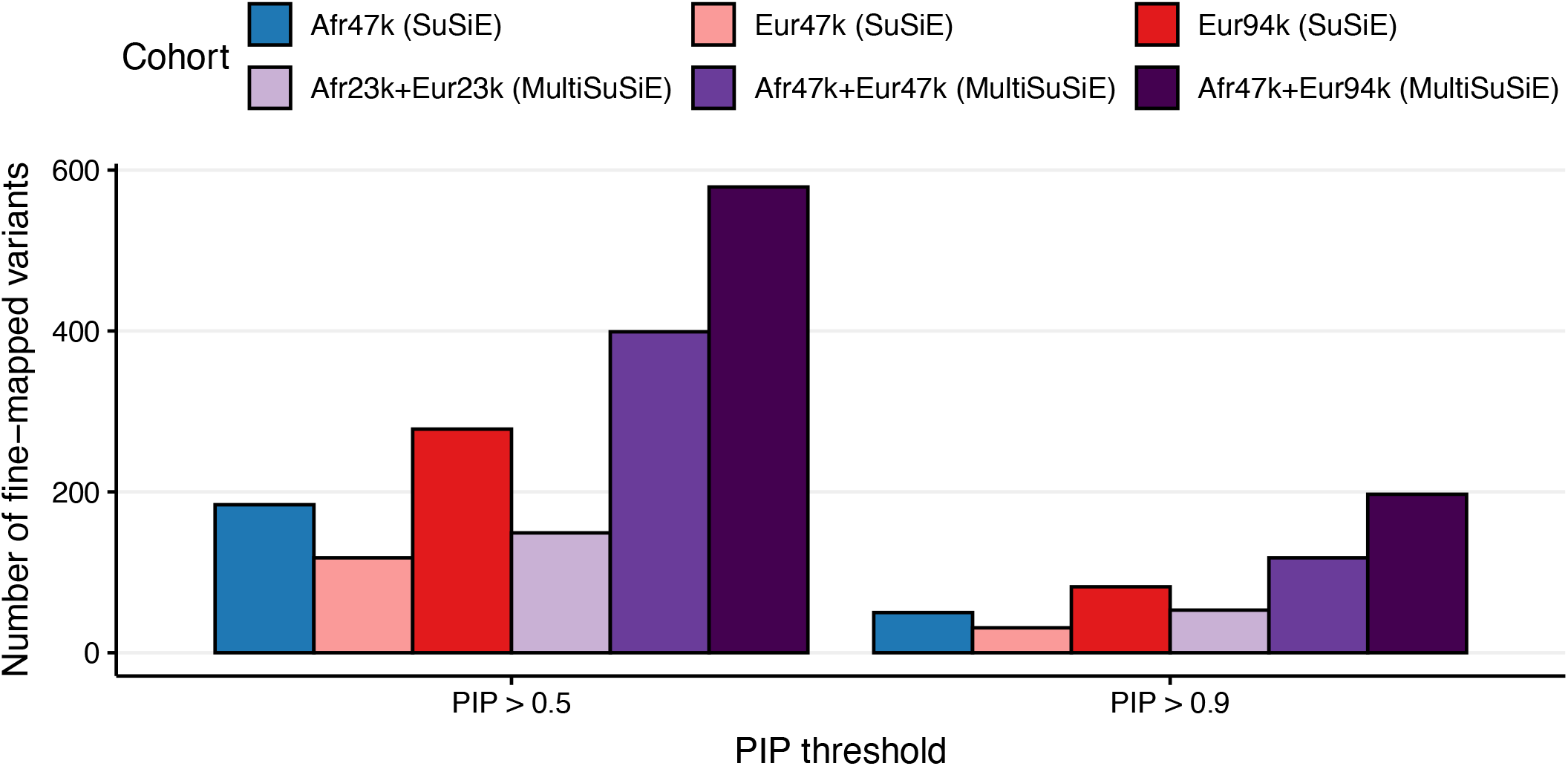
Real trait results across ancestries and sample sizes analyzed. We report the number of variant-trait pairs fine-mapped by MultiSuSiE (for multi-ancestry cohorts) or SuSiE (for single-ancestry cohorts) at PIP > 0.5 and PIP > 0.9 across 14 quantitative traits. Numerical results are reported in Supplementary Table 8.

Next, we compared the performance of 4 multi-ancestry fine-mapping methods: MultiSuSIE, SuSiEx, SCVmeta, and PIPmeta, applied to the Afr47k+Eur47k cohort (Figure 4 and Supplementary Table 9). MultiSuSiE identified 32-94% more variants at PIP > 0.5 than the other three methods (17-76% more variants at PIP > 0.9). We additionally fine-mapped all window-trait pairs using SuSiEx-unfiltered. Consistent with simulations, MultiSuSiE identified only 5% more variants with PIP > 0.5 than SuSiEx-unfiltered (2% more variants with PIP > 0.9) (Supplementary Figure 29). These results suggest that filters by SuSiEx may be overly conservative; however, we have not investigated the setting of fine-mapping with reference LD (see Discussion), which may yield different conclusions.

**Figure 4:**
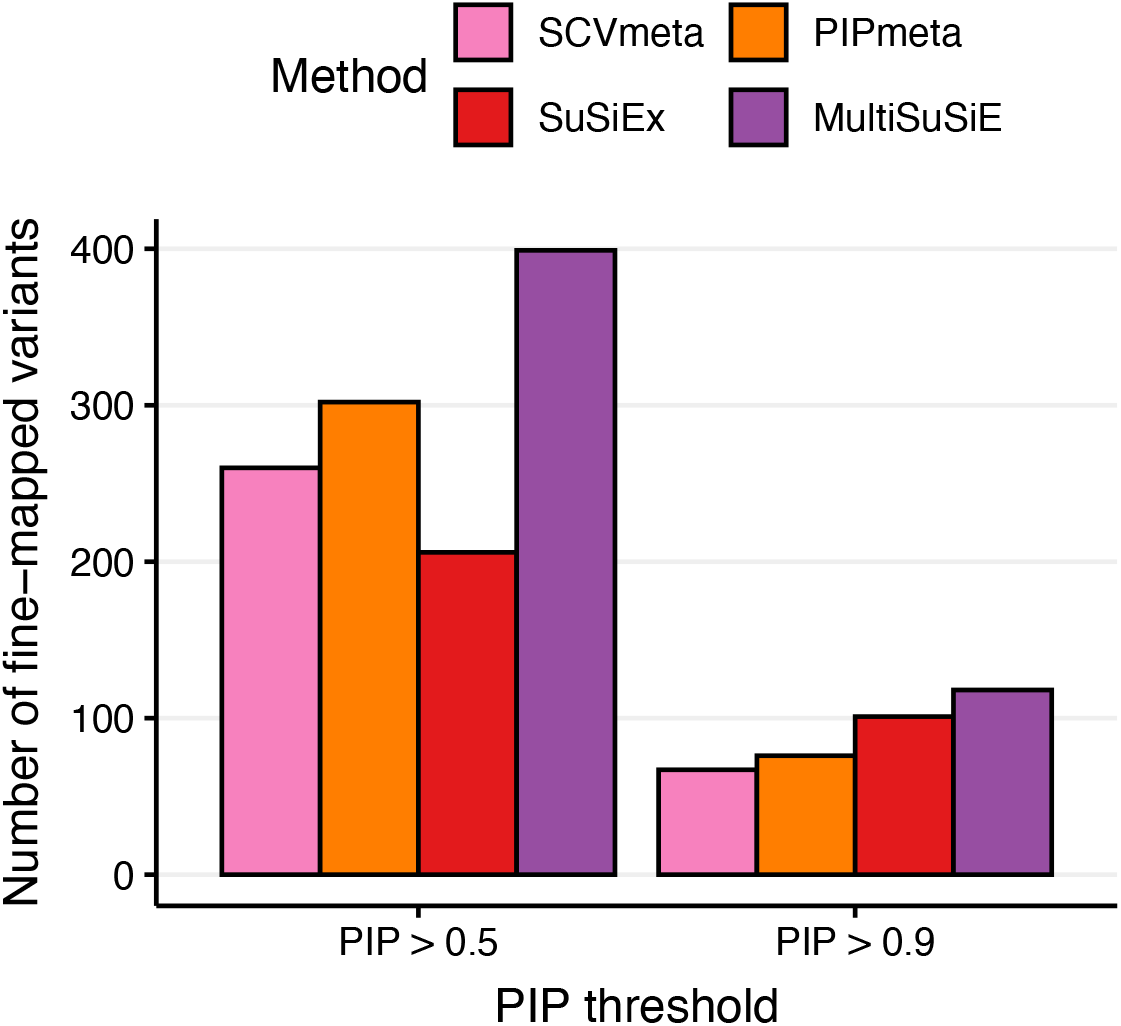
Real trait results for Afr47k+Eur47k across fine-mapping methods. We report the number of variant-trait pairs fine-mapped at PIP > 0.5 and PIP > 0.9 across 14 quantitative traits. Numerical results are reported in Supplementary Table 9.

We validated our fine-mapping results by assessing the functional enrichment of fine-mapped variants^12,44,45^. We assessed the enrichment of PIP > 0.5 variants for 11 approximately independent (absolute correlation < 0.2) binary functional annotations from the baseline-LF or baseline-LD models^46–48^ (Supplementary Table 10; see Methods). First, we varied the ancestry and sample size of the input data, fine-mapping using MultiSuSiE for multi-ancestry cohorts and SuSiE for single-ancestry cohorts (Figure 5 and Supplementary Table 11). We determined that cohorts with African ancestry attained lower functional enrichment (e.g. 2.86 vs. 3.87 for Afr47k+Eur47k vs. Eur94k, *p*=1*10^−4^ for difference; 2.36 vs. 4.34 for Afr47k vs. Eur47k, *p*=3*10^−5^ for difference). However, cohorts with higher power may have lower functional enrichment (due to identification of variants with weaker effects). To correct for this, we compared the enrichment of the top x variants with highest PIP, where x is the minimum number of variants with PIP > 0.5 across cohorts compared (Methods and Supplementary Figure 30). We continued to observe that cohorts with African ancestry attained lower functional enrichment (e.g. 4.07 vs. 4.98 for Afr47k+Eur47k vs. Eur94k, *p*=0.14 for difference; 2.53 vs. 4.34 for Afr47k vs. Eur47k, *p*=1*10^−3^ for difference). Next, we compared the functional enrichment of 4 multi-ancestry fine-mapping methods: MultiSuSIE, SuSiEx, SCVmeta, and PIPmeta, applied to the Afr47k+Eur47k cohort (Supplementary Figure 31). Correcting for differences in power via enrichment of the top x variants with highest PIP, we determined that MultiSuSiE attained similar (non-significantly larger) functional enrichment compared to the other 3 methods. The lower functional enrichment of fine-mapped variants in cohorts with African ancestry is a topic for future investigation, but is also observed when directly quantifying functional enrichment of *χ*^2^ association statistics, hence unlikely to be an artifact of fine-mapping (Supplementary Note, Supplementary Figures 32-36).

**Figure 5:**
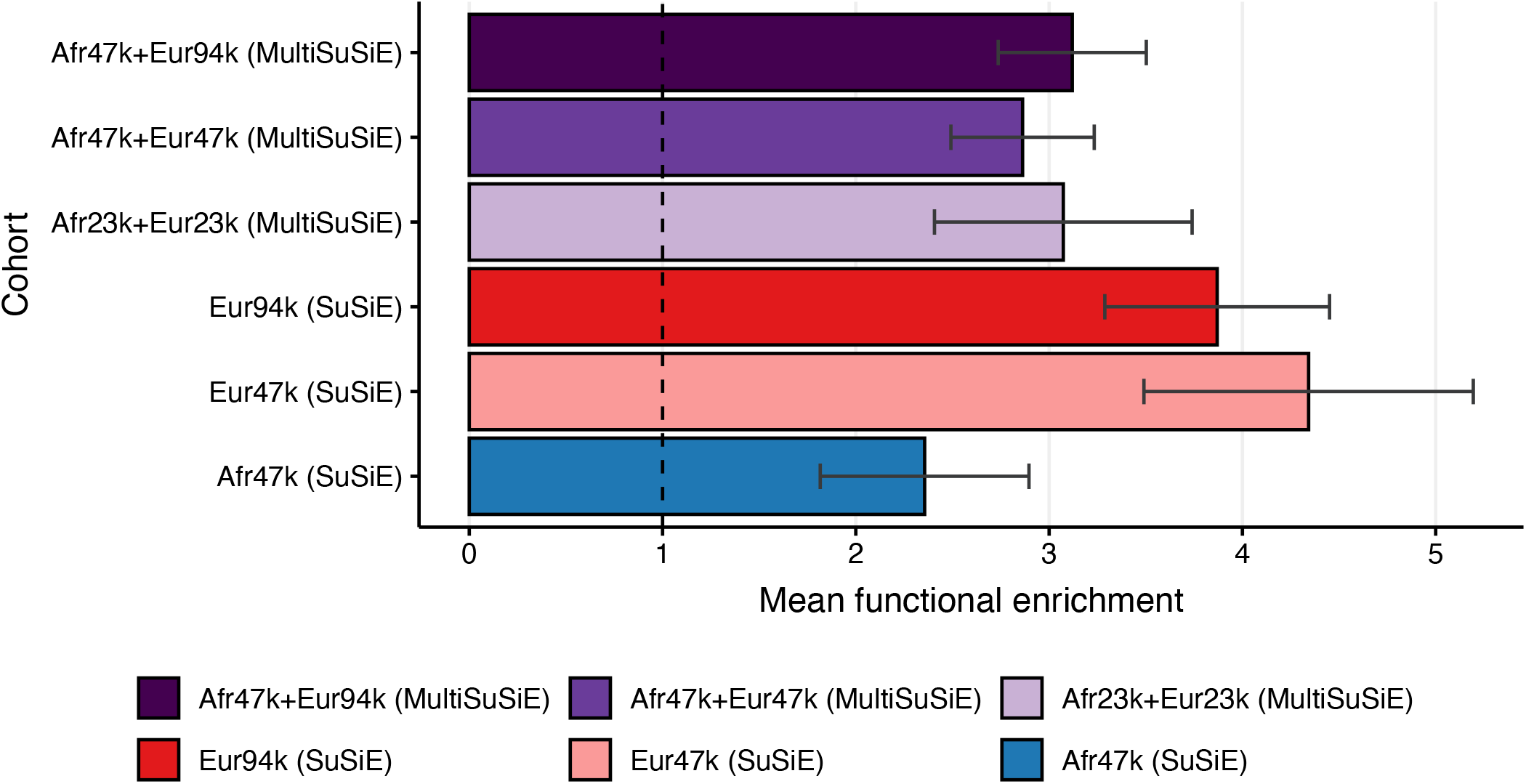
Functional enrichment of fine-mapped variants across ancestries and sample sizes. We report mean functional enrichment of fine-mapped variants (*P*(*a*_*i*_ = 1|*PIP*_*i*_ > 0.5)/ *P*(*a*_*i*_ = 1), where *a*_*i*_ equals 1 if variant *i* is in annotation *a* and 0 otherwise), averaged across 11 functional annotations. Error bars denote 95% confidence intervals based on a genomic block-jackknife with 200 blocks. The vertical dashed bar denotes no functional enrichment. Numerical results are reported in Supplementary Table 11.

### MultiSuSiE identifies well-studied or biologically plausible fine-mapped variants

We have shown that fine-mapped variants (PIP > 0.5) identified by MultiSuSiE using Afr47k+Eur47k may not be identified using other cohorts (Figure 3) or other methods (Figure 4). Below, we dissect four GWAS loci in detail, to investigate the reasons why this may be the case. Fine-mapping results of top variants for all cohorts/methods analyzed are reported in Figure 6a and Supplementary Table 12.

**Figure 6:**
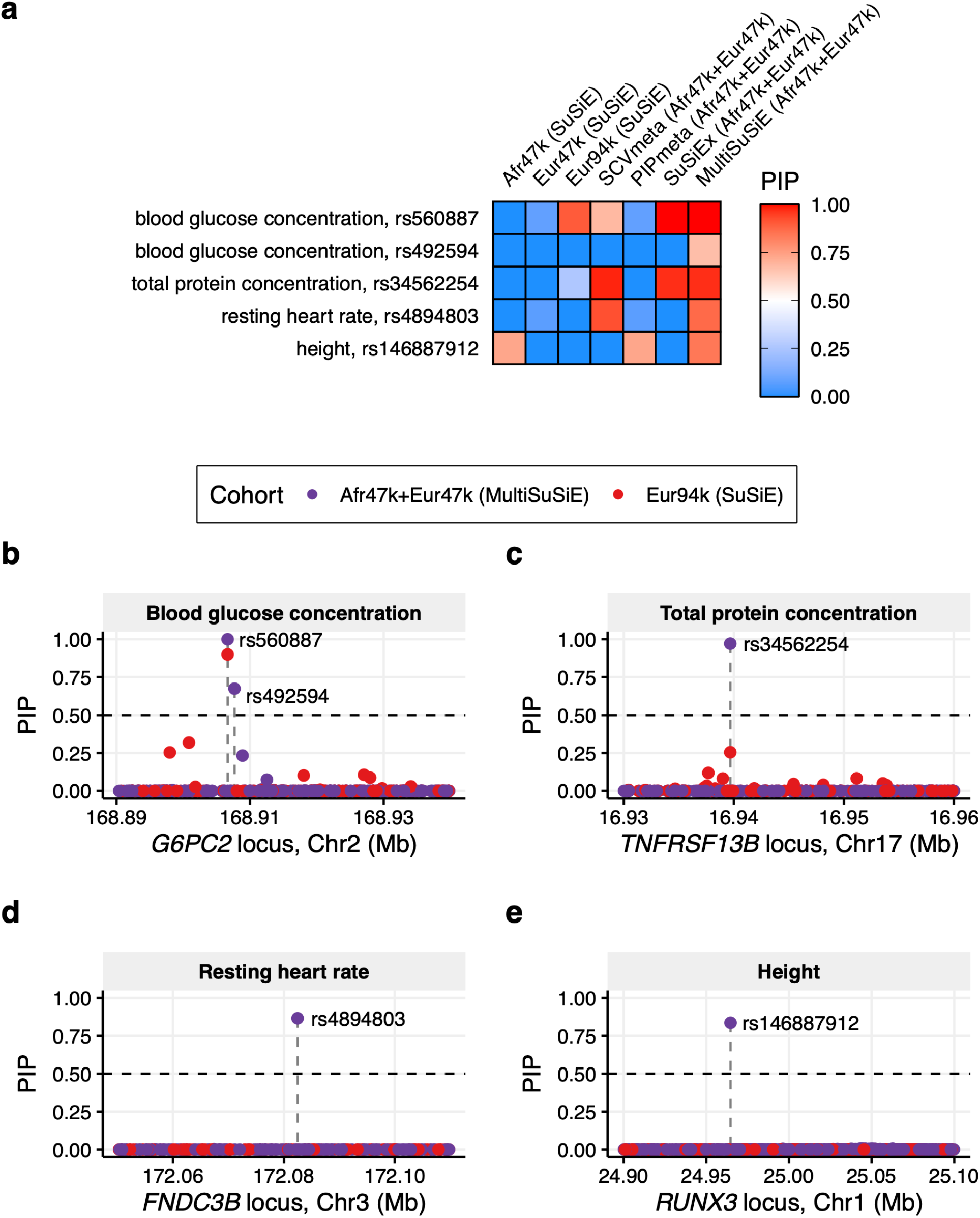
Examples of variants fine-mapped using MultiSuSiE. We report **(a)** PIPs for 5 variants spanning 4 example loci for four multi-ancestry fine-mapping methods applied to Afr47k+Eur47k and SuSiE applied to three single-ancestry cohorts and **(b-e)** PIPs for MultiSuSiE applied to Afr47k+Eur47k and SuSiE applied to Eur94k for the four example loci. Quantitative trait names are provided in grey bars above each panel. Purple and red dots denote PIPs for MultiSuSiE and SuSiE, respectively. Horizontal dashed lines denote PIP=0.5, and vertical dashed lines denote the position of variants with PIP > 0.5. Numerical results are reported in Supplementary Tables 12-13.

First, MultiSuSiE identified two fine-mapped variants (PIP > 0.5) at the *G6PC2* locus for blood glucose concentration (Figure 6b and Supplementary Table 13); both of these variants have previously been validated in experimental assays^49–51^. *G6PC2* is a gene that encodes a subunit of glucose-6-phosphatase (G6Pase), which converts glucose-6-phosphate into glucose, and *G6PC2* knockout has been shown to affect fasting glucose levels in mouse models^52^. The first variant, rs560887, was also fine-mapped using Eur94k but not fine-mapped using Afr47k or Eur47k (Supplementary Table 12). In addition, rs560887 was fine-mapped using SCVmeta and SuSiEx but not fine-mapped using PIPmeta (due to failure to fine-map in each 47k cohort). rs560887 is an intronic variant that has been experimentally shown to impact *G6PC2* splicing^50^. The second variant, rs492594, was not fine-mapped using Afr47k, Eur47k or Eur94k; failure to fine-map rs492594 in Afr47k was due to failure to fine-map rs560887 combined with weak linkage masking^53,54^ with rs560887 (*p*=2.9*10^−4^ for rs492594 in single-variant GWAS vs. *p*=2.6*10^−5^ when adjusting for rs560887, due to positive LD and opposite effect directions), and failure to fine-map rs492594 in Eur47k or Eur94k was due to low evidence of association in Europeans (*p*=0.80 in Eur47k, *p*=0.11 in Eur94k), potentially due to previously reported gene-gene interactions^51,55^. In addition, rs492594 was not fine-mapped using SCVmeta (due to failure to account for weak linkage masking with rs560887 in single causal variant fine-mapping), PIPmeta (due to failure to fine-map in each 47k cohort), or SuSiEx (due to failing its p-value filter, which is based on single-ancestry marginal effect sizes and does not account for linkage masking). rs492594 is a missense variant in a functionally relevant protein motif of G6PC2^51^, and has been shown to decrease G6PC2 protein abundance^49^ and G6Pase activity^51^ in cell line models.

Second, MultiSuSiE fine-mapped a missense variant, rs34562254, at the *TNFRSF13B* locus for total protein concentration (Figure 6c and Supplementary Table 13). *TNFRSF13B* codes for a B cell transmembrane protein involved in adaptive immune response signaling. Rare variation in *TNFRSF13B* is associated with common variable immunodeficiency^56,57^, and *TNFRSF13B* knockout affects antibody levels in mice^58^; antibodies make up approximately 20% of total plasma protein in humans^59^, linking *TNFRSF13B* to total protein concentration. rs34562254 was not fine-mapped using Afr47k, Eur47k, or Eur94k (Supplementary Table 12); failure to fine-map rs34562254 in Afr47k was due to low GWAS effect size (*p*=3.1*10^−4^ in Afr47k), and failure to fine-map rs34562254 in Eur94k was due to high levels of LD (LD 4^th^ moment = 29 in Eur94k vs. 4.0 in Afr47k). Additionally, rs34562254 was fine-mapped using SuSiEx and SCVmeta, but not by PIPmeta (due to failure to fine-map in each 47k cohort). rs34562254 is a missense variant for *TNFRSF13B* with CADD score^60^ of 17 (top 1.9%), SIFT score^61^ of 0.03 (deleterious), and PolyPhen-2^62^ score of 0.73 (possibly damaging). rs34562254 has been previously implicated for total protein concentration by GWAS^63^, but not by statistical fine-mapping, to the best of our knowledge.

Third, MultiSuSiE fine-mapped a conserved variant, rs4894803, at the *FNDC3B* locus for resting heart rate (Figure 6d and Supplementary Table 13). *FNDC3B* codes for a circular RNA transcript, circ-FNDC3B, in addition to its regular protein product; circ-FNDC3B is involved in multiple aspects of cardiovascular physiology including cardiomyocte apoptosis^64^, blood vessel formation^64^, and regulation of ADAM10, a gene whose overexpression is related to aortic aneurysm^65^. rs4894803 was not fine-mapped using Afr47k, Eur47k, or Eur94k; failure to fine-map rs4894803 using Afr47k was due to lower MAF (0.14 in Afr47k vs 0.42 in Eur47k), and failure to fine-map rs4894803 using Eur47k and Eur94k was due to high levels of LD (LD 4^th^ moment = 24 in Eur47k vs 6.1 in Afr47k). rs4894803 was also fine-mapped by SCVmeta, but not by SuSiEx (due to failing its p-value filter) or PIPmeta (due to failure to fine-map in each 47k cohort). rs4894803 has a CADD^60^ score of 22.8 (top 0.52%) and is highly conserved with a GERP^66^ score of 2,545 (*p*=5*10^−^ 233), and Zoonomia placental mammal PhyloP^67,68^ score of 6.3 (top 0.51%). rs4894803 has been previously implicated for resting heart rate by GWAS^69^, but not by statistical fine-mapping, to the best of our knowledge.

Fourth, MultiSuSiE fine-mapped a single-nucleotide insertion, rs146887912, at the *RUNX3* locus for height (Figure 6e, Supplementary Table 13). *RUNX3* is a tumor suppressor gene^70^ that encodes a transcription factor; *RUNX3* knockout affects limb length, bone development, and osteopenia risk in mice^71,72^. rs146887912 was fine-mapped using Afr47k, but not Eur47k or Eur94k due to its lower MAF in Europeans (0.034 in Afr47k vs 0.0043 in Eur47k). Additionally, rs146887912 was fine-mapped by PIPmeta, but not SuSiEx (due to its purity filter) or SCVmeta (due to the presence of multiple causal variants at the locus). rs146887912 is a single-nucleotide insertion in the 5’-untranslated region of *RUNX3*, is in a polypyrimidine tract of an alternative transcript for *RUNX3*, and lies in an Ensembl predicted promoter region^73^. rs146887912 has not been previously implicated for any trait by GWAS or statistical fine-mapping^74^, to the best of our knowledge, although the surrounding locus has been previously implicated for height^75,76^. This may be because most GWAS use imputed genotypes; rs146887912 is not present in Haplotype Reference Consortium-imputed UK Biobank data, but is present in UKBB WGS data (at ancestry-specific allele frequencies similar to AoU), highlighting the utility of fine-mapping with WGS data.

## Discussion

We have developed MultiSuSiE, a fast and powerful method for multi-ancestry fine-mapping, and applied MultiSuSiE to 14 quantitative traits leveraging WGS data in 47k African-ancestry and 94k European-ancestry individuals from All of Us; our analysis of WGS data avoids ancestry-based differences in imputation quality in genotyping chip data^26^, which may produce false-positive discoveries^27,28^. MultiSuSiE extends the approach introduced by SuSiE^13,18^ to efficiently search the high-dimensional space of causal variant configurations, while allowing effect sizes to vary across ancestries. MultiSuSiE attained higher power and/or much lower computational cost than two other very recently proposed multi-ancestry extensions of SuSiE^21,22^, making analysis of large-scale All of Us data feasible. MultiSuSiE also outperformed conventional meta-analysis-based multi-ancestry fine-mapping strategies^15,34–36^. Fine-mapping using African-ancestry and multi-ancestry cohorts was more powerful than fine-mapping using European-ancestry cohorts at matched sample sizes, with differences between African-ancestry vs. European-ancestry cohorts explained by differences in LD 4th moments. We highlighted several examples where MultiSuSiE implicates well-studied or biologically plausible fine-mapped variants that were not implicated by other methods.

MultiSuSiE, SuSiEx^21^, and MESuSiE^22^ all extend SuSiE to accommodate multiple ancestries, but differ in their model assumptions and algorithmic details. First, MultiSuSiE and SuSiEx assume that all causal variants are shared across ancestries, but MESuSiE does not. Thus, MESuSiE estimates many more parameters, greatly increasing running time (Table 2); in principle this choice enables MESuSiE to avoid a potential source of model misspecification, but in practice we showed that MultiSuSiE is not sensitive to model misspecification arising from ancestry-specific causal variants (Supplementary Figures 15-16). Second, SuSiEx assumes that the effect sizes of shared causal variants are uncorrelated across ancestries, while MultiSuSiE and MESuSiE do not. However, this assumption appears to have little impact in practice (Supplementary Figures 9 and 20). Instead, the difference in performance between MultiSuSiE and SuSiEx is driven by two filters applied by SuSiEx on the estimated credible sets (Supplementary Figure 9). These methodological distinctions are summarized in Supplementary Table 14. We note that MCMC-based approaches such as PAINTOR^6,23,37^ and MGfm^11^ had far larger running times (Table 2) and were not included in our large-scale simulations or analyses of real traits, but SuSiEx^21^ and MESuSiE^22^ have previously been reported to outperform PAINTOR in smaller-scale simulations.

We recommend the use of MultiSuSiE in preference to other methods for multi-ancestry fine-mapping based on its higher power and/or much lower computational cost (a particularly important consideration in multi-ancestry fine-mapping due to the larger number of polymorphic variants). We recommend against excluding variants that are not polymorphic in all ancestries, as we observed that this increases FDR and decreases power in simulations (Supplementary Figures 9 and 25). We recommend analyzing WGS data when available, and using in-sample LD^18,24^ (see below).

Our work has several limitations. First, we excluded variants with MAF < 0.01 in both Eur47k and Afr47k. Exclusion of causal variants could in principle induce false-positive results at tagging variants that survive this filter; however, the functional enrichment of fine-mapped (PIP > 0.5) variants that are rare or low-frequency (MAF < 0.05) in at least one population is similar to that of variants that are common (MAF > 0.05) in both populations (2.85 vs 2.88, *p* = 0.91 for difference), suggesting that our exclusion criteria did not produce substantial false positives in practice. Second, as with all fine-mapping methods, exclusion of variants that are difficult to genotype, even in WGS data, (including tandem repeats^77–79^ and other structural variants^80,81^) could induce false-positive results at tagging variants. Indeed, at the *HBA* locus for mean corpuscular volume, SuSiE using Afr47k fine-mapped (at PIP > 0.5) 17 single nucleotide variants, 1 insertion, and 1 deletion (Supplementary Table 15); all fine-mapped variants were jointly significant in a multiple linear regression model. We hypothesize that these are false positive discoveries, driven by LD with a previously reported allelic series of copy number variants at the locus with large causal effects on blood cell traits^80^; thus, accurate genotyping of non-SNP variants is an important future research direction. Third, as with all multiple causal variant fine-mapping methods, we recommend applying MultiSuSiE using in-sample LD^18,24^; when in-sample LD is not available, an alternative is to use LD reference panels from each target population that span at least 10% of the target GWAS sample size, as previously recommended^12^. Fourth, consistent with previous reports of imperfect calibration of SuSiE PIPs^12,31^, MultiSuSiE PIPs are not perfectly calibrated; however, MultiSuSiE is well-calibrated with respect to a conservative FDR upper bound (1 – PIP threshold) proposed in previous work^12^ (Figure 2, Supplementary Figure 2). Fifth, we focus here on fine-mapping using data from a single WGS study; caution is warranted when fine-mapping across studies or fine-mapping using imputed genotypes, which may increase false discovery rates^27^. Sixth, causal variants fine-mapped in cohorts with African ancestry have lower functional enrichment. This phenomenon is consistent across fine-mapping methods and is also observed when directly quantifying functional enrichment of *χ*^2^ association statistics, hence is unlikely to be an artifact of fine-mapping (Supplementary Note); further investigation of this phenomenon is a direction for future research. Seventh, we have not applied MultiSuSiE to binary traits, which require careful consideration of sample size, minor allele frequency, and disease prevalence to ensure correct calibration^82,83^. Eighth, we have not applied MultiSuSiE to leverage functional priors^12^ or analyze three or more ancestry groups, but these use cases are supported by our software. Despite all these limitations, MultiSuSiE is a fast and powerful method for multi-ancestry fine-mapping.

## Data Availability

All of Us summary statistics for the 14 traits analyzed in Eur94k, Afr47k, and Eur47k are available at https://zenodo.org/records/11111186 (DOI: 10.5281/zenodo.11111186). In accordance with the All of Us Data and Statistics Dissemination Policy, summary statistics for variant-trait-cohort combinations with a minor allele count less than 40 have been censored. All of Us v7 short read individual-level whole-genome sequencing data is available to authorized users on the AoU Researcher Workbench. MultiSuSiE fine-mapping results generated in this study are available at https://zenodo.org/records/11111186 (DOI: 10.5281/zenodo.11111186). UK Biobank data is available at http://www.ukbiobank.ac.uk.

## Supplementary Note

### Lower functional enrichment of fine-mapped variants with Afr47k

As discussed in the main text, cohorts with African ancestry have lower functional enrichment of fine-mapped variants, which persists when correcting for differences in fine-mapping power and considering different fine-mapping methods (Figure 5, Supplementary Figures 30-31). (We believe that the functional enrichment difference is not driven by ancestry-biased functional annotations, because (i) we excluded annotations derived from large genotyping or whole-genome sequencing datasets and (ii) the difference is consistent across the 11 annotations (Supplementary Figure 32)).

We performed two additional analyses to investigate this finding. First, we stratified Afr47k into two cohorts based on each individual’s proportion of European ancestry (Afr23k-low and Afr23k-high, with 11.6% and 24.6% mean European ancestry, respectively), and performed fine-mapping in each cohort using SuSiE; we analyzed 1,431 3Mb windows that contained a variant in the central 1Mb with GWAS *p*<5*10^−6^ in at least one of the two stratified cohorts, restricting to variants with MAF > 0.01 in Afr47k. We determined that Afr23k-high and Afr23k-low identified similar numbers of fine-mapped variants at each PIP threshold (Supplementary Figure 33) and that the functional enrichment of variants fine-mapped at PIP > 0.5 was non-significantly larger in Afr23k-low compared to Afr23k-high (1.93 vs 1.53, *p*=0.12 for difference; Supplementary Figure 34). These findings do not provide statistically significant evidence that admixture-LD in cohorts with African ancestry^38^ greatly impacts fine-mapping results.

Second, we directly quantified the functional enrichment of *χ*^2^ association statistics in Afr47k, Eur47k, Afr23k-low, and Afr23k-high (we did not apply S-LDSC^84^, which has significant complexities in analyses of admixed populations^85^). We computed 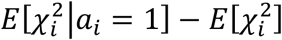, where 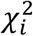 is the marginal *χ*^2^ association statistic for variant *i* and *a*_*i*_ is the annotation value for variant *i*. We determined that functional enrichment of *χ*^2^ association statistics for variants with MAF > 0.01 was significantly larger in Eur47k compared to Afr47k (0.0267 vs. 0.0117, *p*=2*10^−11^ for difference, Supplementary Figure 35); results for MAF > 0.001 were similar (0.0244 vs. 0.0104, p=2*10^−12^ for difference). For completeness, we also computed a normalized functional enrichment statistic,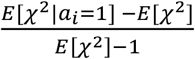, which we expect to be attenuated towards 0 due to LD (which increases the denominator), and more attenuated towards 0 in European-ancestry cohorts (due to higher LD). The normalized quantity remained (non-significantly) larger in Eur47k than Afr47k (0.344 vs. 0.303 for MAF > 0.01, *p*=0.28 for difference, 0.345 vs. 0. 296 for MAF > 0.001, *p*=0.19 for difference), despite the expectation of greater attenuation towards 0 in Eur47k. However, we determined that the (non-normalized) functional enrichment of *χ*^2^ association statistics for variants with MAF > 0.01 was larger (with nominal significance) in Afr23k-low compared to Afr23k-high (0.00712 vs. 0.00527, *p*=0.015 for difference, Supplementary Figure 36).

The lower functional enrichment of fine-mapped variants in cohorts with African ancestry is a topic for future investigation, as these results do not establish the cause of this phenomenon.

This finding is not specific to MultiSuSiE, but also observed then fine-mapping using other methods (Supplementary Figure 31) or directly quantifying functional enrichment of *χ*^2^ association statistics (Supplementary Figure 35).

## Figures

**Supplementary Figure 1:**
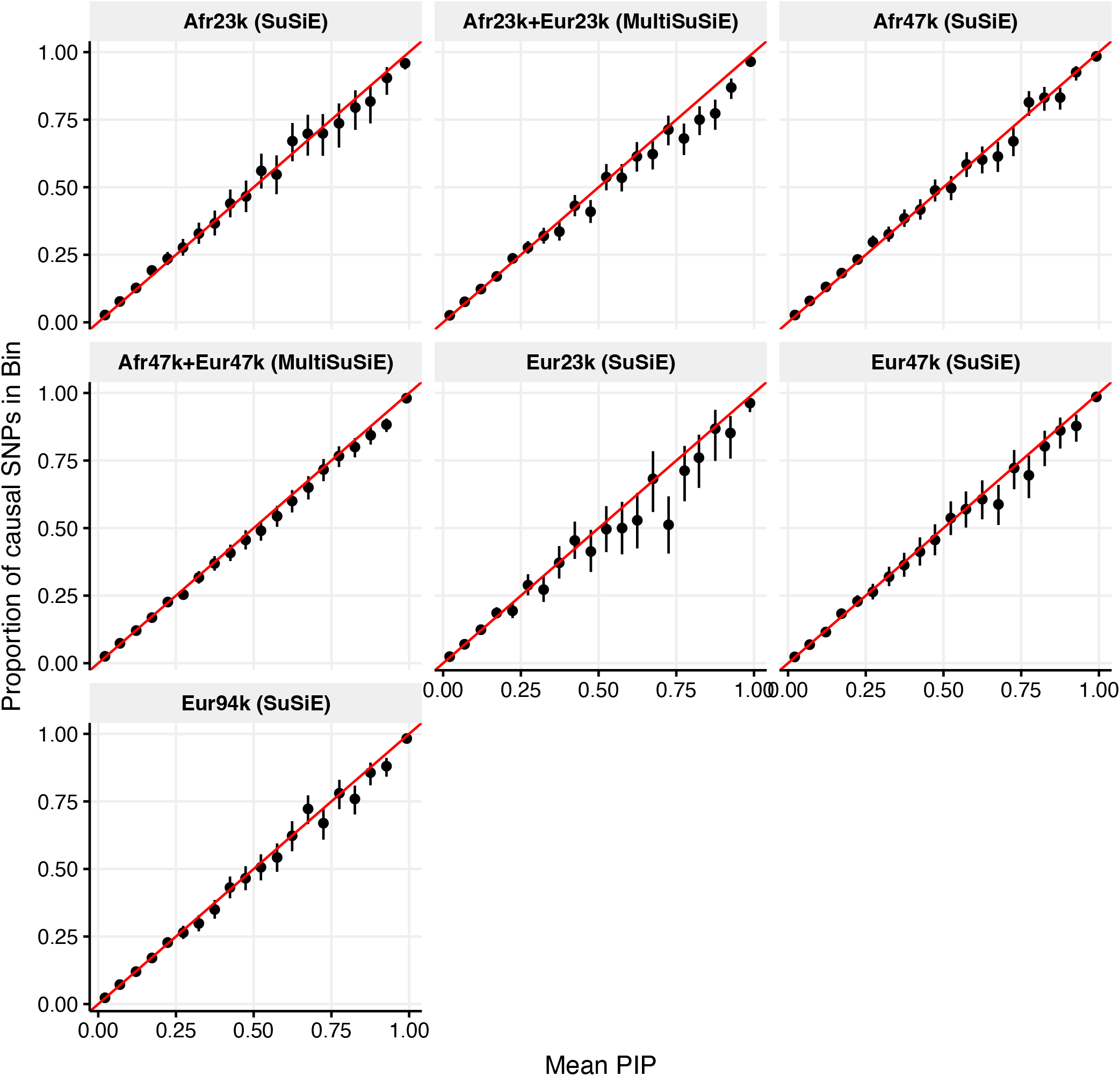
PIP calibration across ancestries and cohorts analyzed. We report the mean PIP and proportion of causal variants of 20 equally spaced PIP-bins of variants with PIP > .01. Error bars denote 95% Agresti-Coull binomial proportion confidence intervals. The red line has an intercept of 0 and slope of 1 and corresponds to perfect PIP calibration.

**Supplementary Figure 2:**
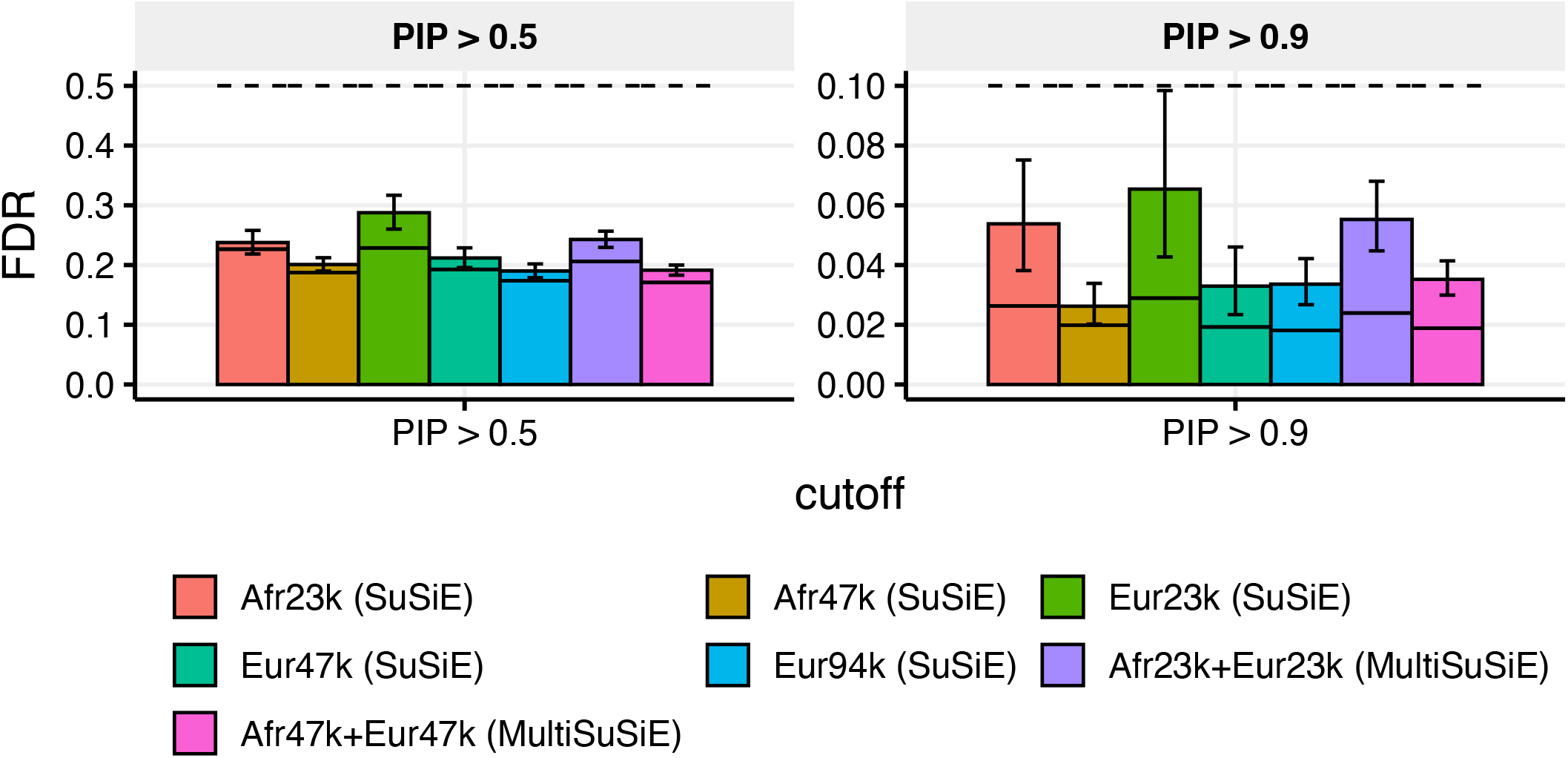
FDR across ancestries and sample sizes analyzed, including two additional cohorts with low sample size, Afr23k and Eur23k. We report the FDR (bars), conservative FDR upper bound (dashed line), and expected FDR (solid bars) at PIP > 0.5 and PIP > 0.9. Error bars denote 95% Agresti-Coull binomial proportion confidence intervals.

**Supplementary Figure 3:**
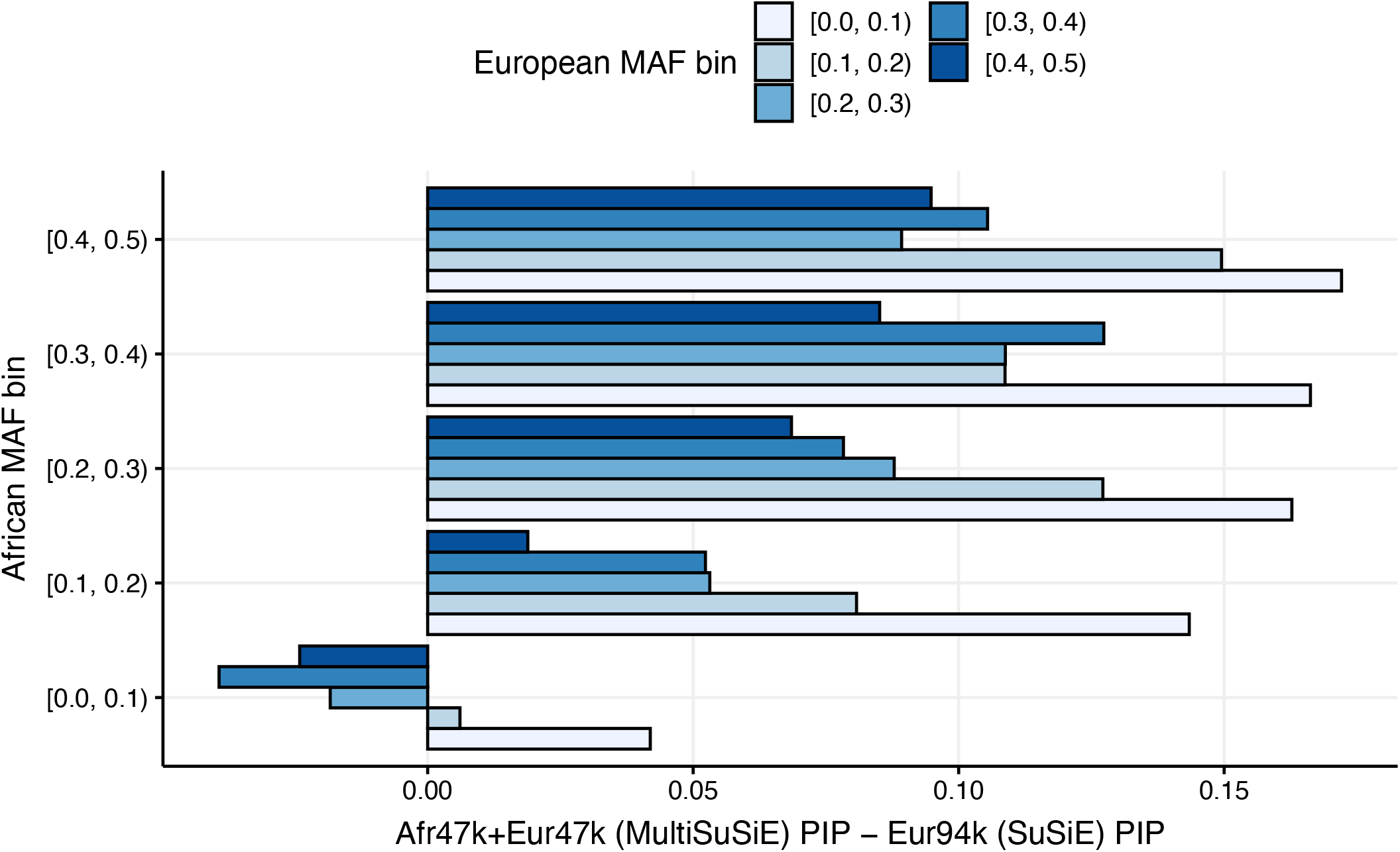
Mean difference in PIP between Afr47k+Eur47k and Eur94k, stratified by African and European MAF of variant. We report the difference in PIP between MultiSuSiE applied to Afr47k+Eur47k and SuSiE applied to Eur94k, averaged across variants.

**Supplementary Figure 4:**
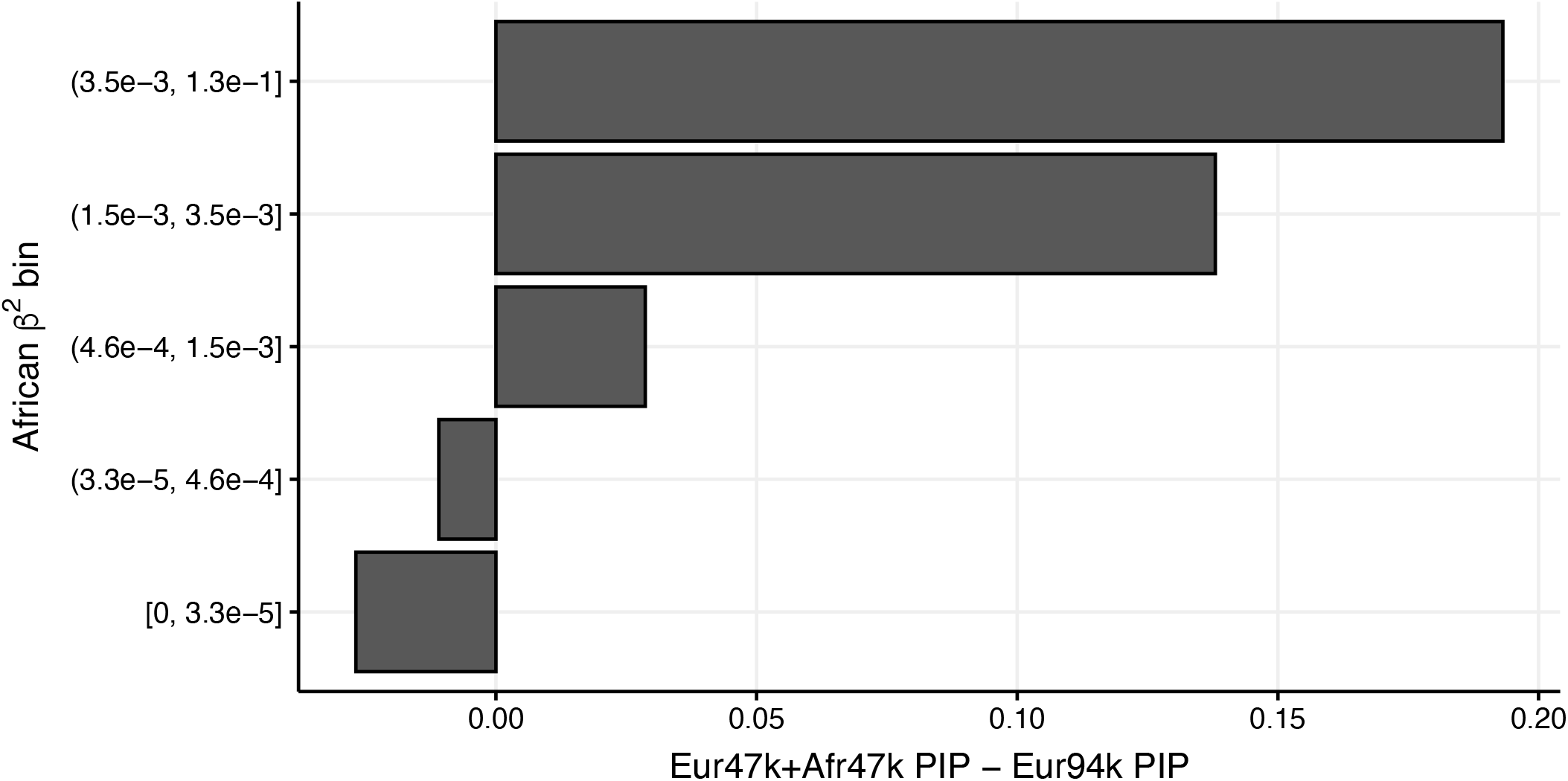
Mean difference in PIP between Afr47k+Eur47k and Eur94k, stratified by African per-allele effect size. We report the difference in PIP between MultiSuSiE applied to Afr47k+Eur47k and SuSiE applied to Eur94k, averaged across variants.

**Supplementary Figure 5:**
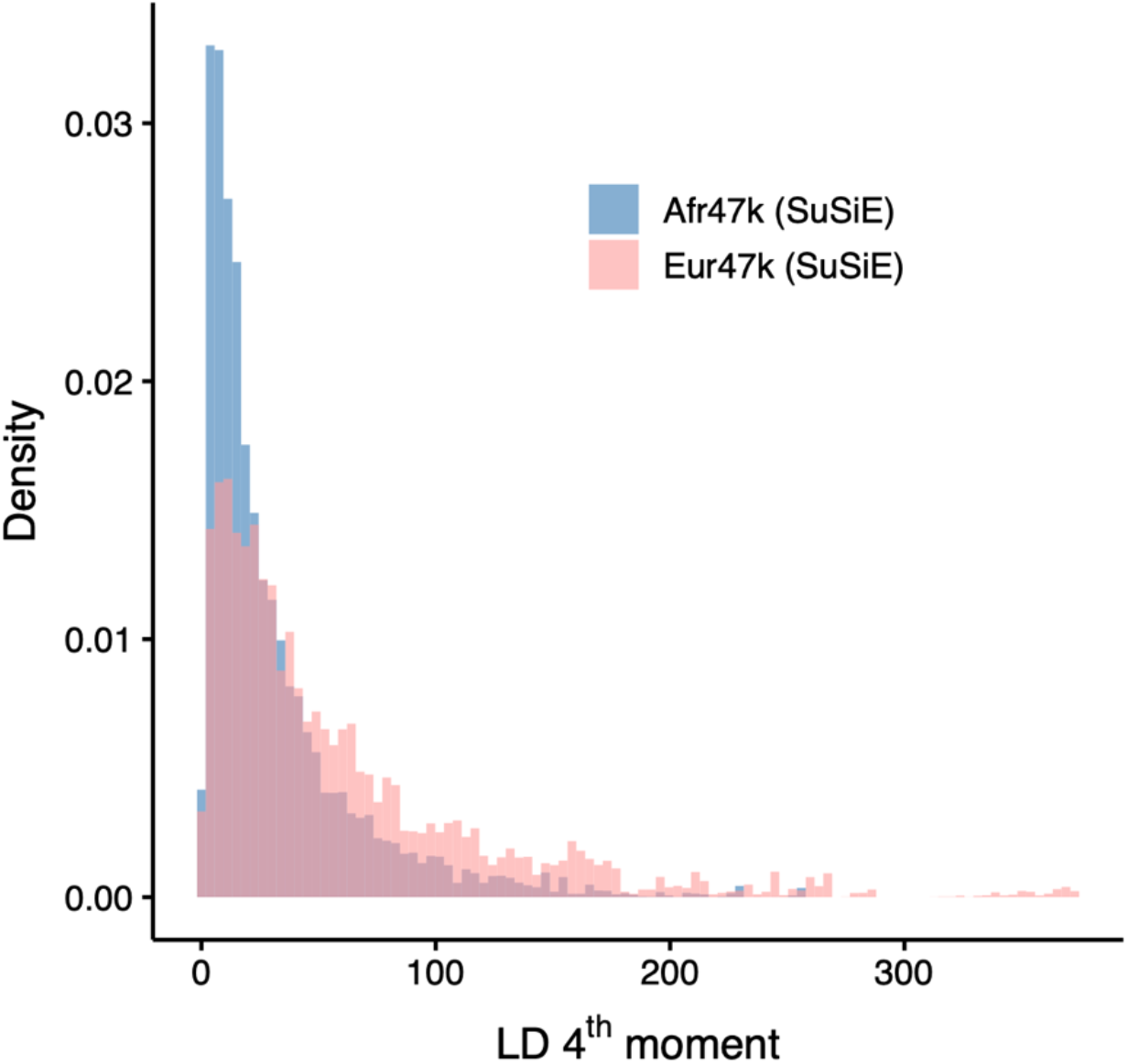
Distribution of LD 4^th^ Moments for variants with MAF > 0.05 in Afr47k and Eur47k. For each ancestry, we report the proportion of variants with MAF > 0.05 in that ancestry with LD 4^th^ moment in each of 100 equally spaced bins, pooled across all 10 3Mb simulated loci.

**Supplementary Figure 6:**
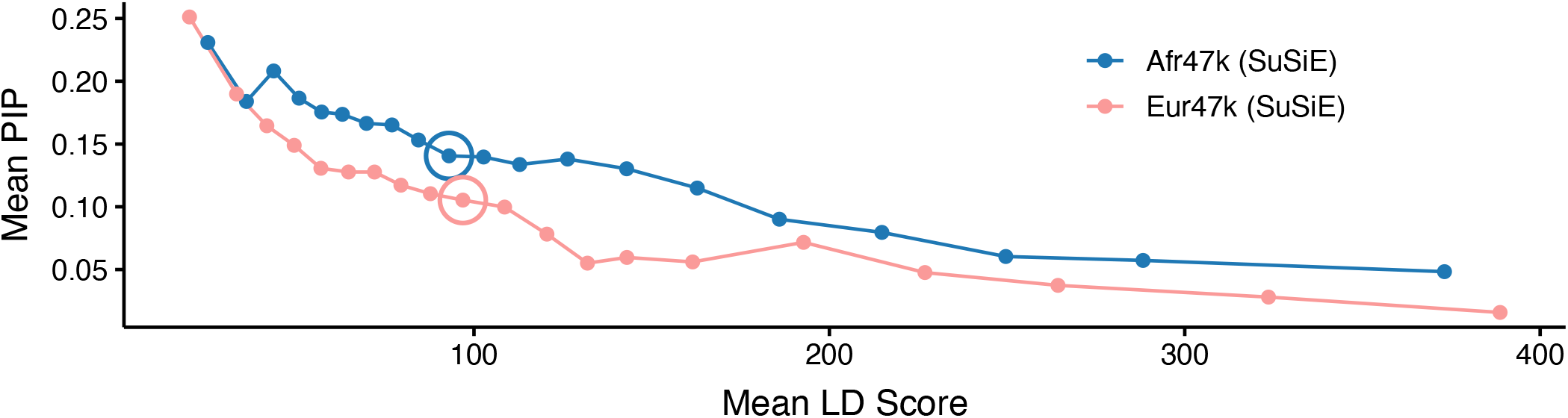
Relationship between LD score and PIP for common causal variants in fine-mapping simulations of Afr47k and Eur47k. For each ancestry, we report the mean PIP of causal variants with MAF > .05 in that ancestry in 20 equally sized LD score bins in Afr47k and Eur47k fine-mapping simulations. Circled dots denote the 10^th^ LD score bin.

**Supplementary Figure 7:**
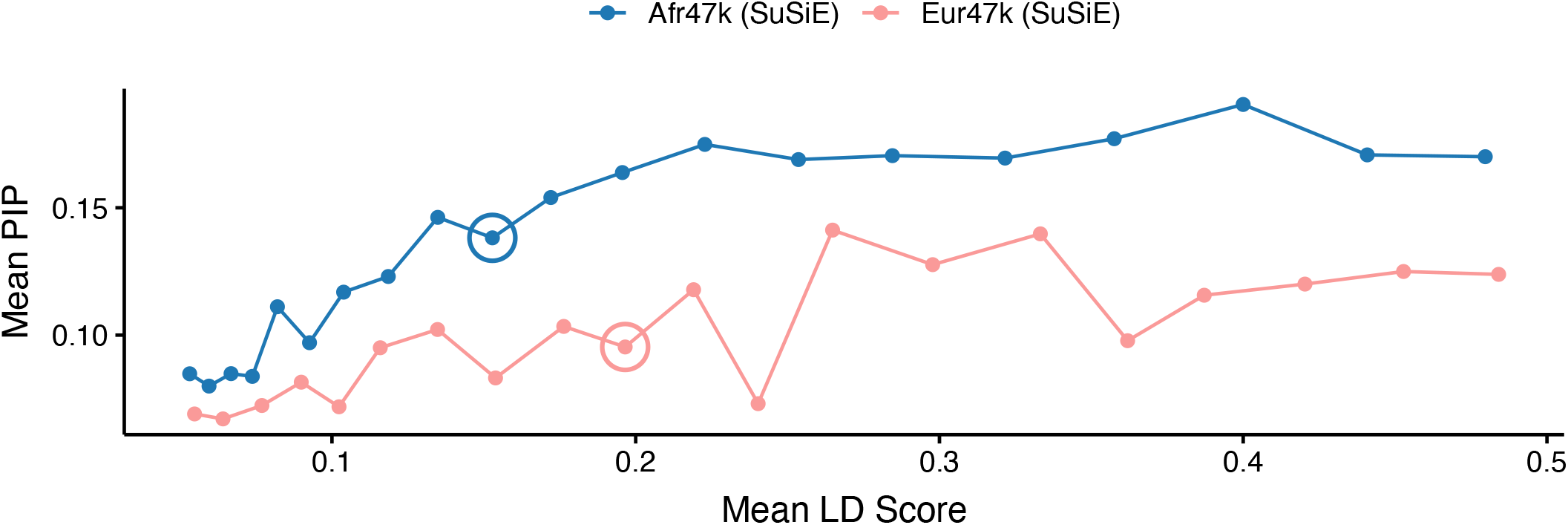
Relationship between MAF and PIP for common causal variants in fine-mapping simulations of Afr47k and Eur47k. For each ancestry, we report the mean PIP of causal variants with MAF > .05 in that ancestry in 20 equally sized MAF bins in Afr47k and Eur47k fine-mapping simulations. Circled dots denote the 10^th^ LD score bin.

**Supplementary Figure 8:**
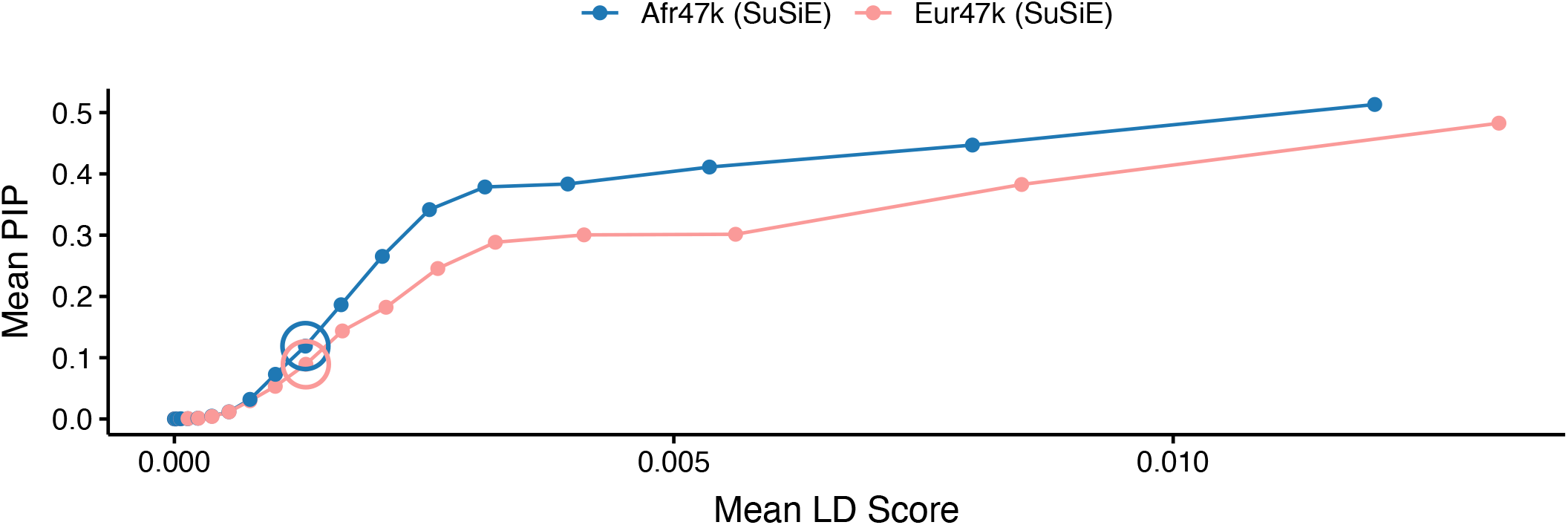
Relationship between squared per-allele effect size and PIP for common causal variants in fine-mapping simulations of Afr47k and Eur47k. For each ancestry, we report the mean PIP of causal variants with MAF > .05 in that ancestry in 20 equally sized squared per-allele effect size bins in Afr47k and Eur47k fine-mapping simulations. Circled dots denote the 10^th^ LD score bin.

**Supplementary Figure 9:**
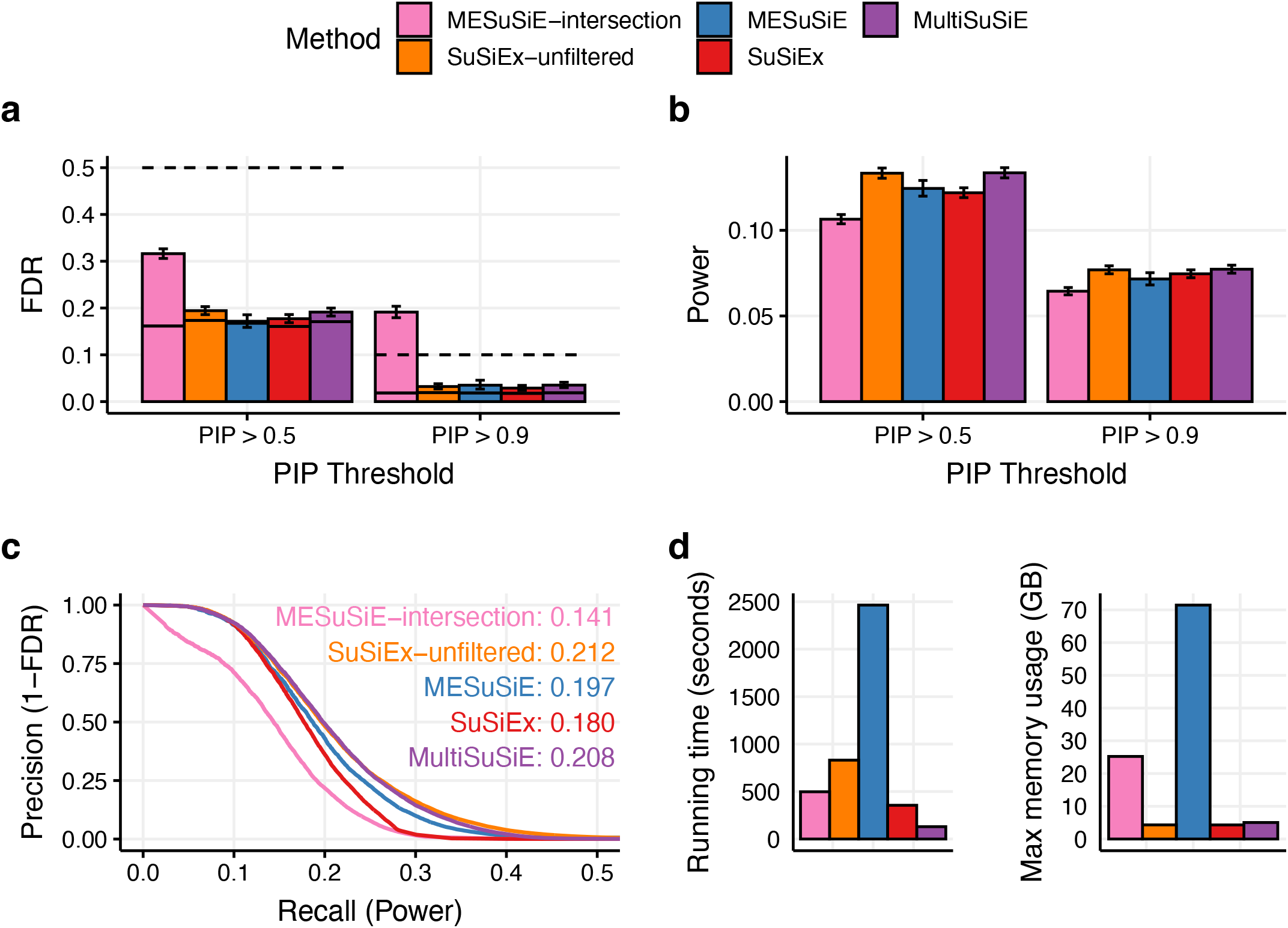
Simulation results for Afr47k+Eur47k across fine-mapping methods, including MESuSiE-intersection and SuSiEx-unfiltered. We report **(a)** the FDR (bars), conservative FDR upper bound (dashed line), and expected FDR (solid bars) at PIP > 0.5 and PIP > 0.9, **(b)** power at PIP > 0.5 and PIP > 0.9, **(c)** Precision-recall curves varying PIP threshold. Upper right hand text indicates the AUPRC of each method. **(d)** Running times and memory requirements for all methods. Error bars denote 95% Agresti-Coull binomial proportion confidence intervals.

**Supplementary Figure 10:**
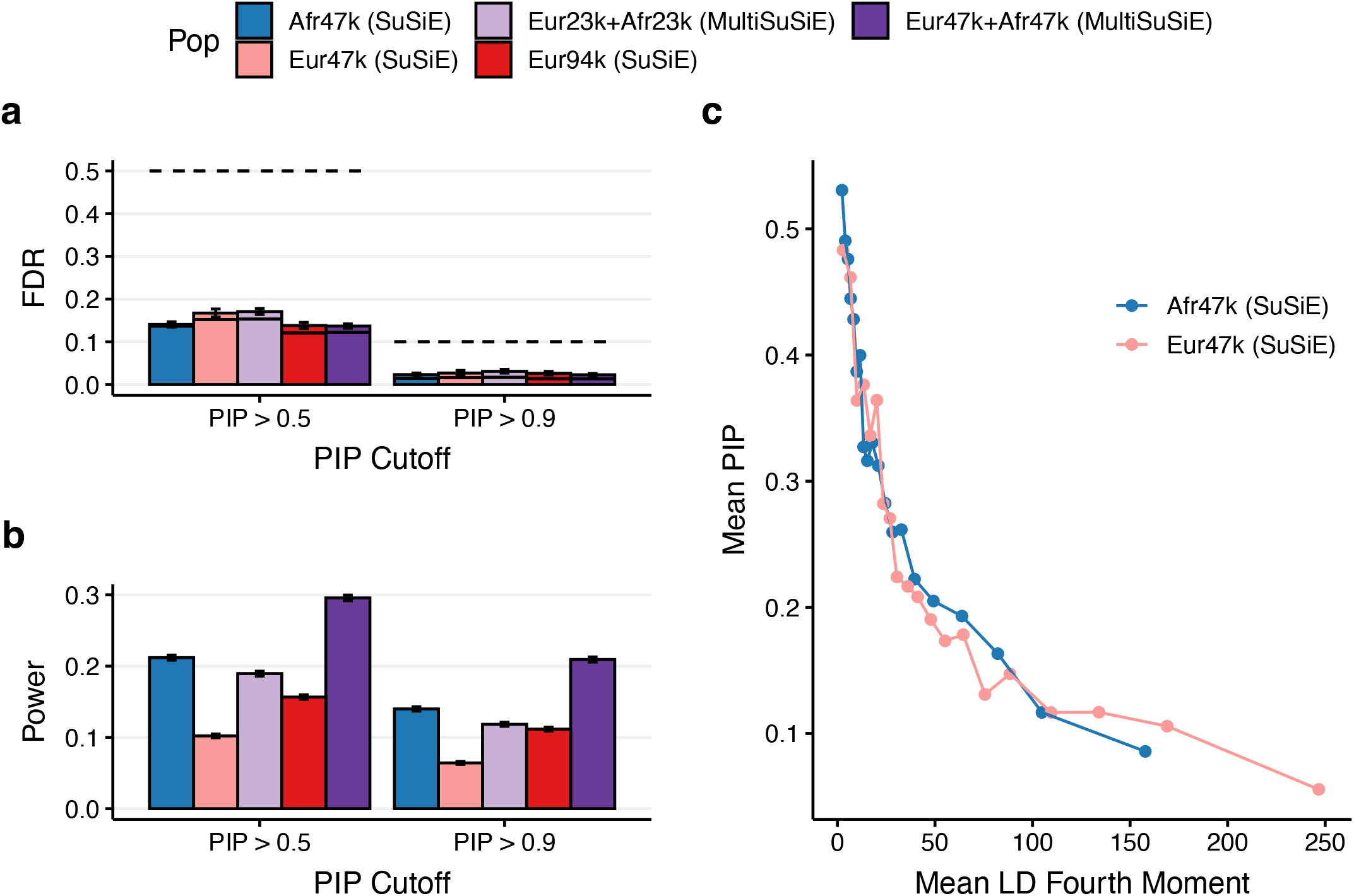
Simulation results across ancestries and sample sizes in simulations with increased h^2^. We report **(a)** the FDR (bars), conservative FDR upper bound (dashed line), and expected FDR (solid bars) at PIP > 0.5 and PIP > 0.9, **(b)** power at PIP > 0.5 and PIP > 0.9, **(c)** Mean PIP of causal variants with MAF > .05 within 20 equally sized LD 4^th^ moment bins in Afr47k and Eur47k fine-mapping. Error bars denote 95% Agresti-Coull binomial proportion confidence intervals.

**Supplementary Figure 11:**
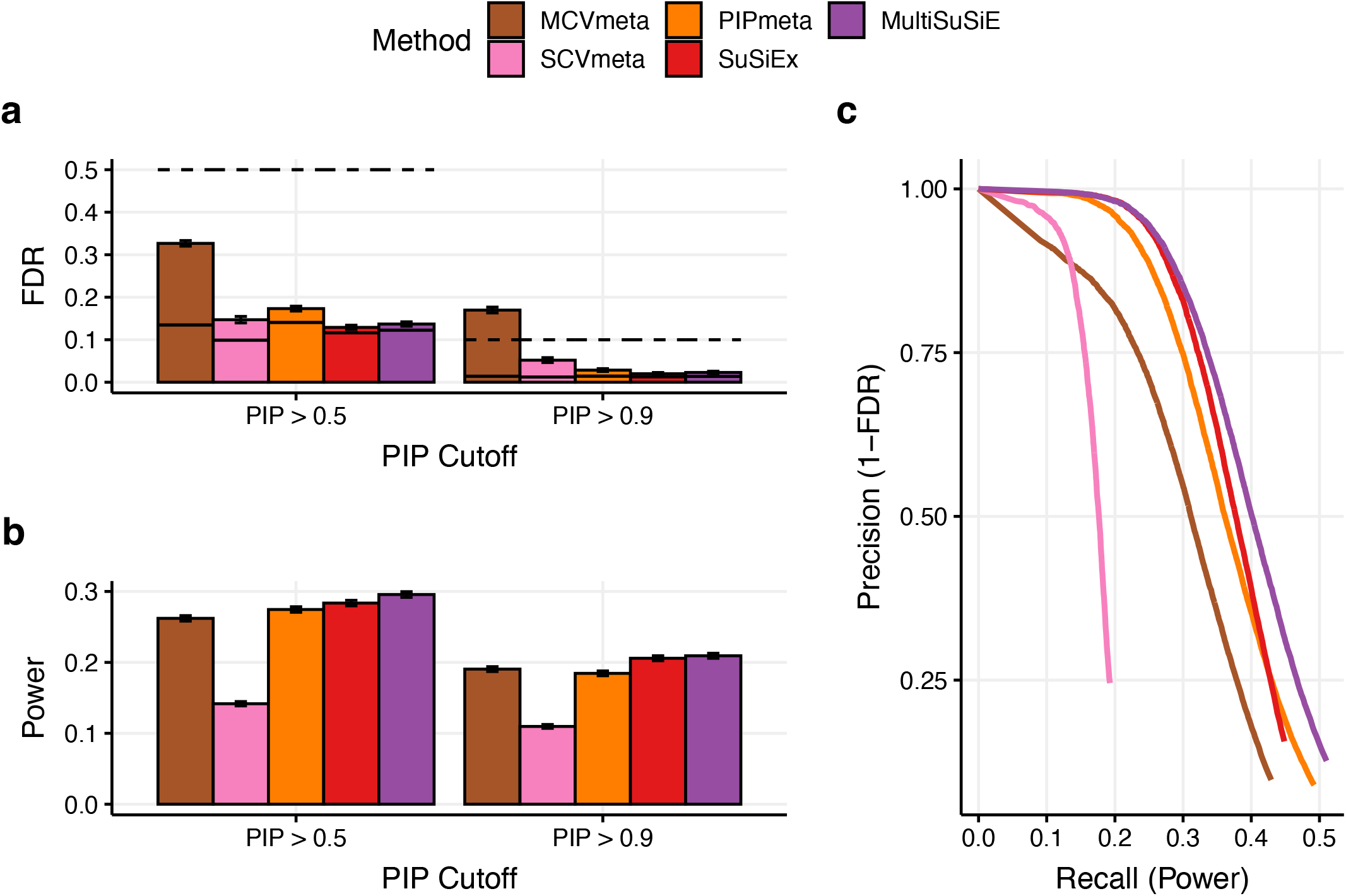
Simulation results for Afr47k+Eur47k across fine-mapping methods in simulations with increased h^2^. We report **(a)** the FDR (bars), conservative FDR upper bound (dashed line), and expected FDR (solid bars) at PIP > 0.5 and PIP > 0.9, **(b)** power at PIP > 0.5 and PIP > 0.9, **(c)** precision-recall curves varying PIP threshold from 1 to 0.01. Error bars denote 95% Agresti-Coull binomial proportion confidence intervals.

**Supplementary Figure 12:**
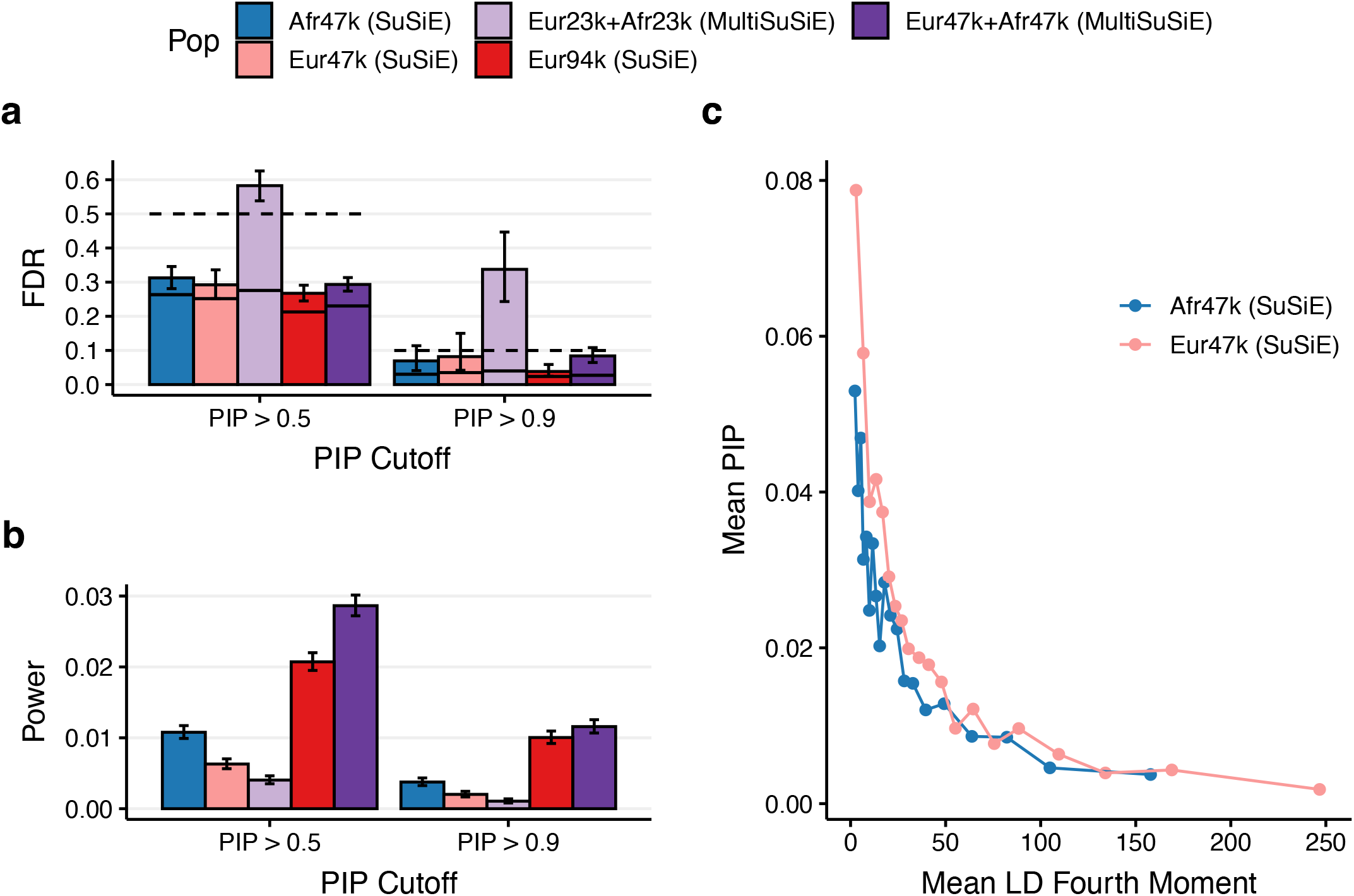
Simulation results across ancestries and sample sizes in simulations with decreased h^2^. We report **(a)** the FDR (bars), conservative FDR upper bound (dashed line), and expected FDR (solid bars) at PIP > 0.5 and PIP > 0.9, **(b)** power at PIP > 0.5 and PIP > 0.9, **(c)** Mean PIP of causal variants with MAF > .05 within 20 equally sized LD 4^th^ moment bins in Afr47k and Eur47k fine-mapping. Error bars denote 95% Agresti-Coull binomial proportion confidence intervals.

**Supplementary Figure 13:**
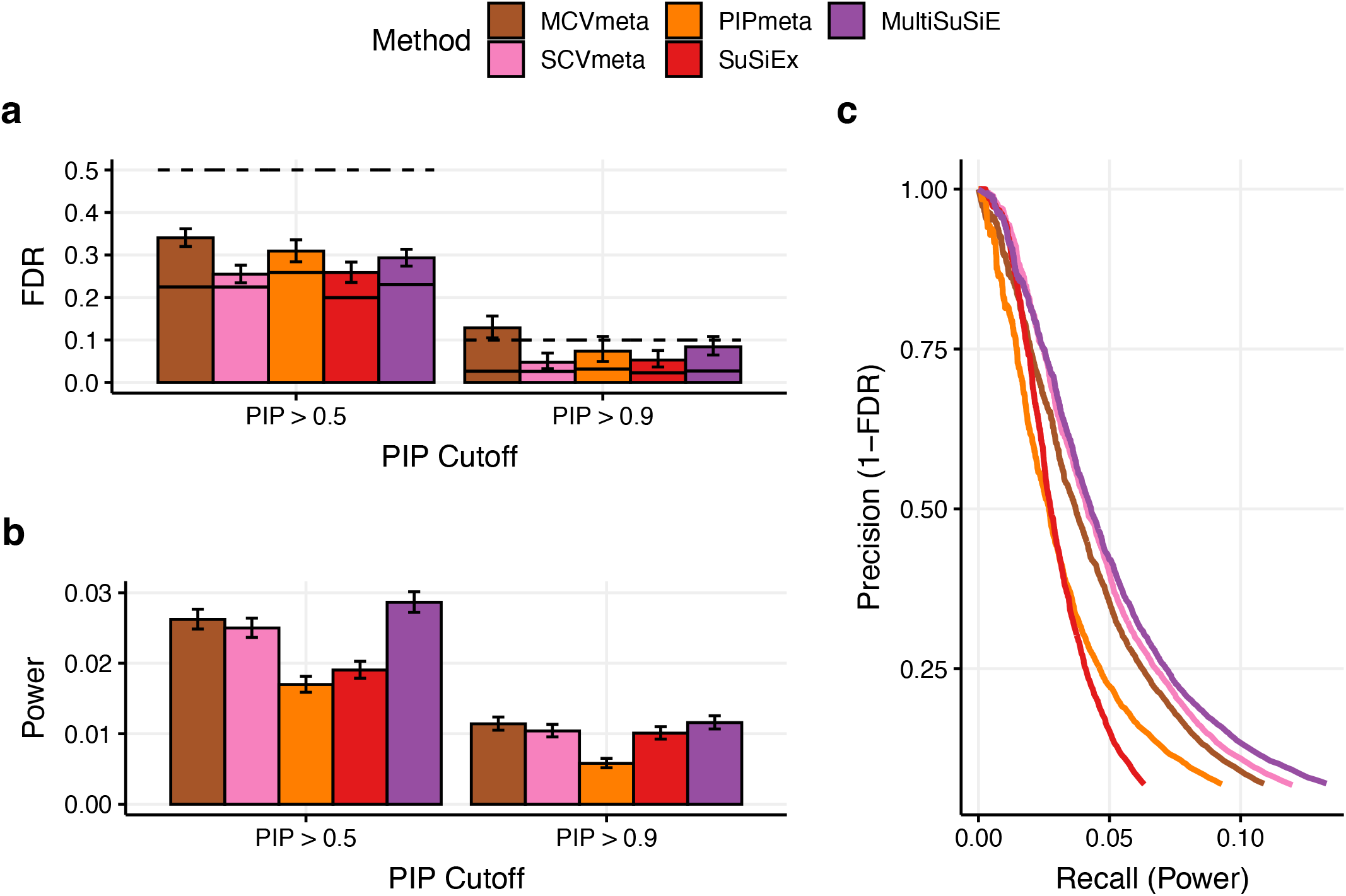
Simulation results for Afr47k+Eur47k across fine-mapping methods in simulations with decreased h^2^. We report **(a)** the FDR (bars), conservative FDR upper bound (dashed line), and expected FDR (solid bars) at PIP > 0.5 and PIP > 0.9, **(b)** power at PIP > 0.5 and PIP > 0.9, **(c)** precision-recall curves varying PIP threshold from 1 to 0.01. Error bars denote 95% Agresti-Coull binomial proportion confidence intervals.

**Supplementary Figure 14:**
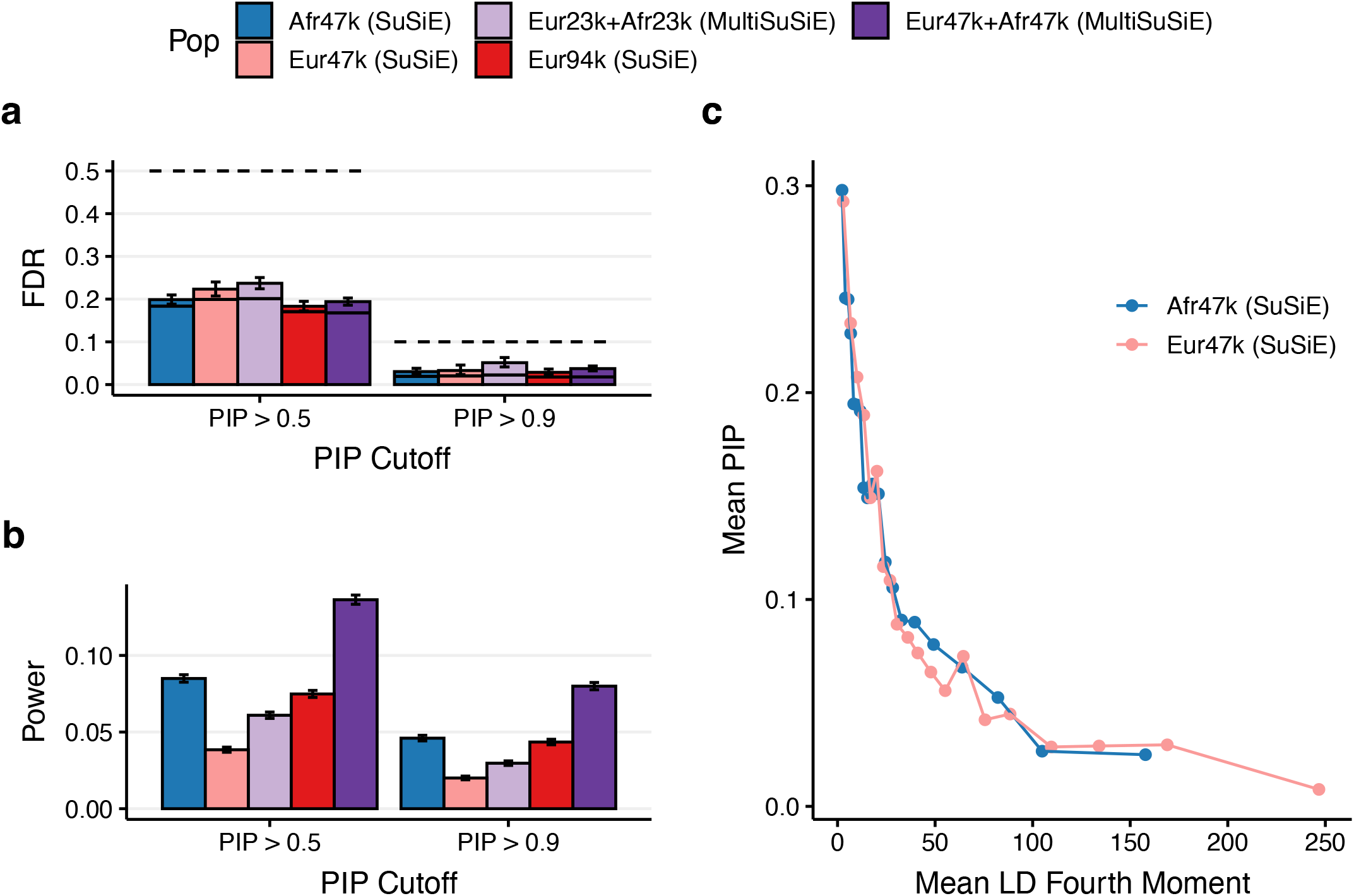
Simulation results across ancestries and sample sizes in simulations with differences in causal variant identity across ancestries but equal per-allele effect sizes at shared causals. We report **(a)** the FDR (bars), conservative FDR upper bound (dashed line), and expected FDR (solid bars) at PIP > 0.5 and PIP > 0.9, **(b)** power at PIP > 0.5 and PIP > 0.9, **(c)** Mean PIP of causal variants with MAF > .05 within 20 equally sized LD 4^th^ moment bins in Afr47k and Eur47k fine-mapping. Error bars denote 95% Agresti-Coull binomial proportion confidence intervals.

**Supplementary Figure 15:**
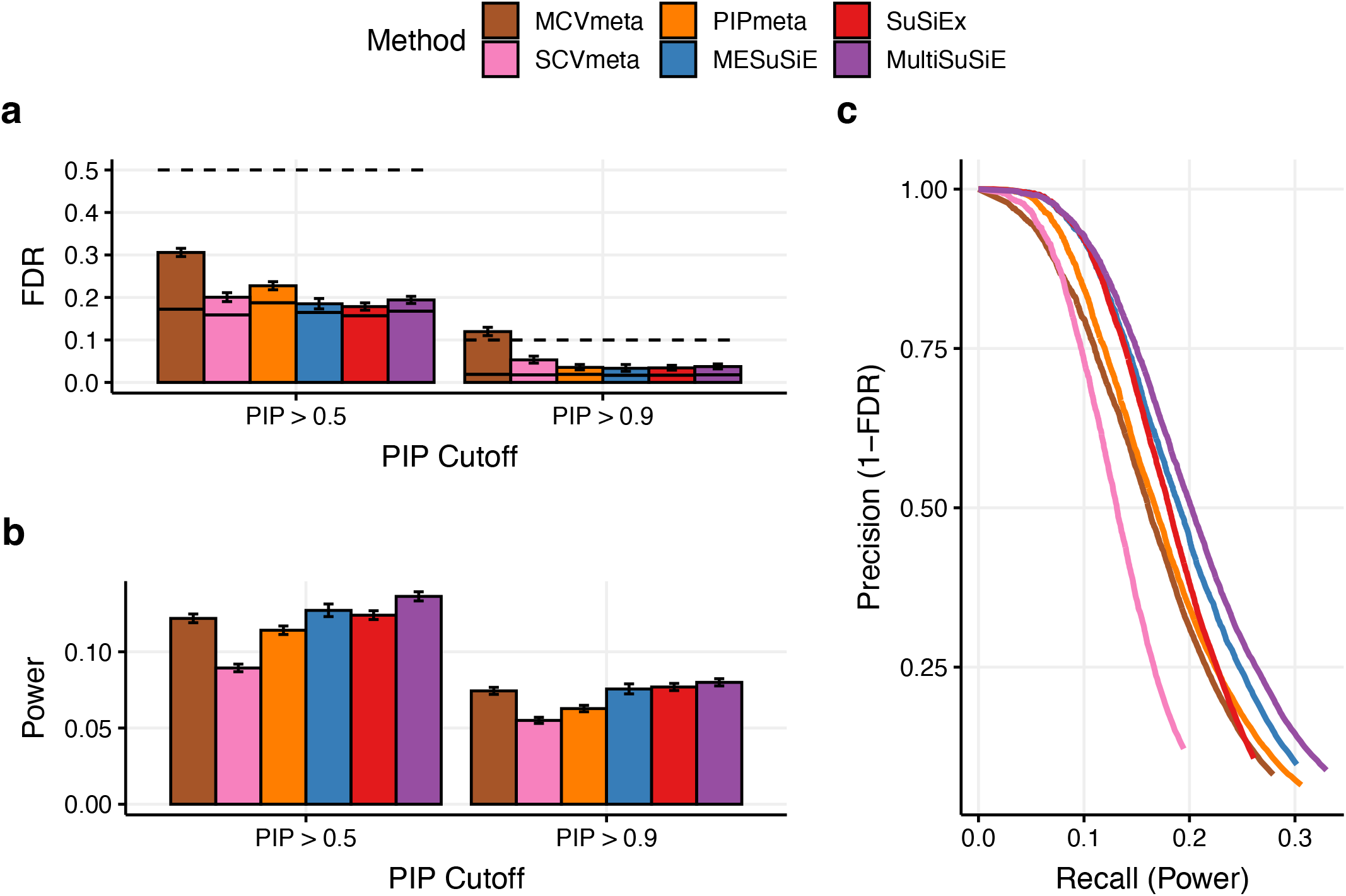
Simulation results for Afr47k+Eur47k across fine-mapping methods in simulations with differences in causal variant identity across ancestries but equal per-allele effect sizes at shared causals. We report **(a)** the FDR (bars), conservative FDR upper bound (dashed line), and expected FDR (solid bars) at PIP > 0.5 and PIP > 0.9, **(b)** power at PIP > 0.5 and PIP > 0.9, **(c)** precision-recall curves varying PIP threshold from 1 to 0.01. Error bars denote 95% Agresti-Coull binomial proportion confidence intervals.

**Supplementary Figure 16:**
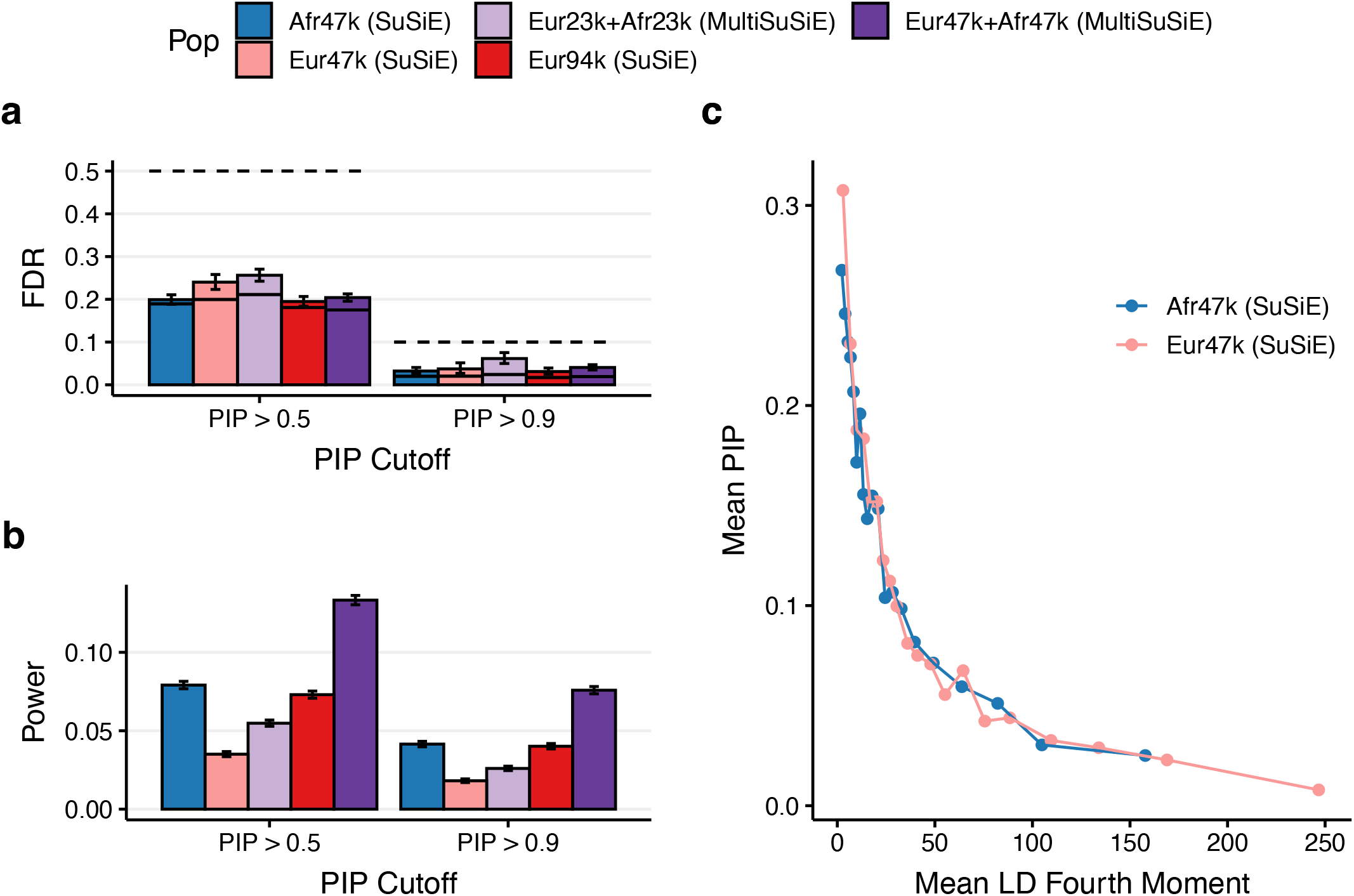
Simulation results across ancestries and sample sizes in simulations with identical causal variant identity across ancestries and differences in per-allele effect sizes. We report **(a)** the FDR (bars), conservative FDR upper bound (dashed line), and expected FDR (solid bars) at PIP > 0.5 and PIP > 0.9, **(b)** power at PIP > 0.5 and PIP > 0.9, **(c)** Mean PIP of causal variants with MAF > .05 within 20 equally sized LD 4^th^ moment bins in Afr47k and Eur47k fine-mapping. Error bars denote 95% Agresti-Coull binomial proportion confidence intervals.

**Supplementary Figure 17:**
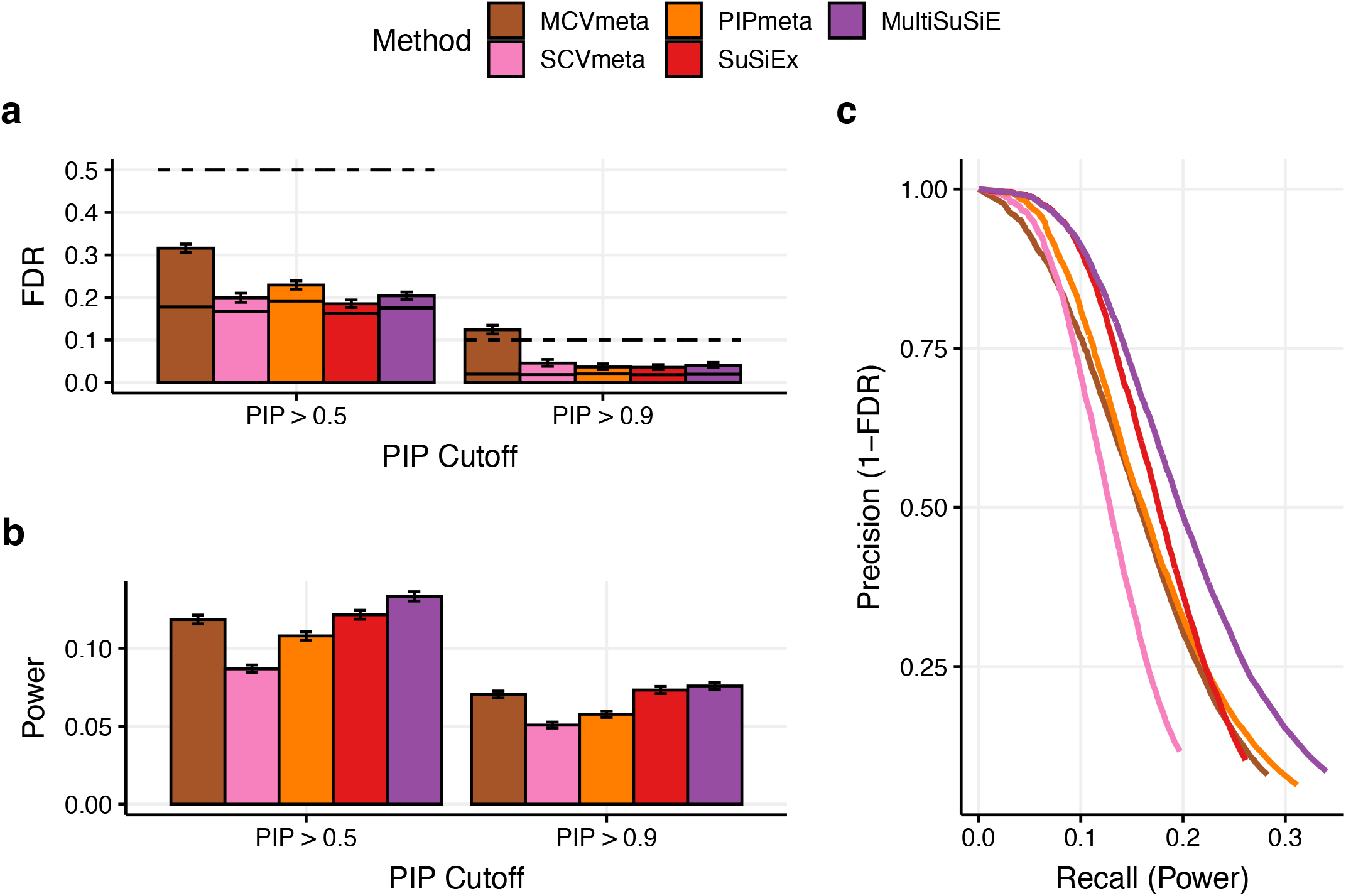
Simulation results for Afr47k+Eur47k across fine-mapping methods in simulations with identical causal variant identity across ancestries and differences in per-allele effect sizes. We report **(a)** the FDR (bars), conservative FDR upper bound (dashed line), and expected FDR (solid bars) at PIP > 0.5 and PIP > 0.9, **(b)** power at PIP > 0.5 and PIP > 0.9, **(c)** precision-recall curves varying PIP threshold from 1 to 0.01. Error bars denote 95% Agresti-Coull binomial proportion confidence intervals.

**Supplementary Figure 18:**
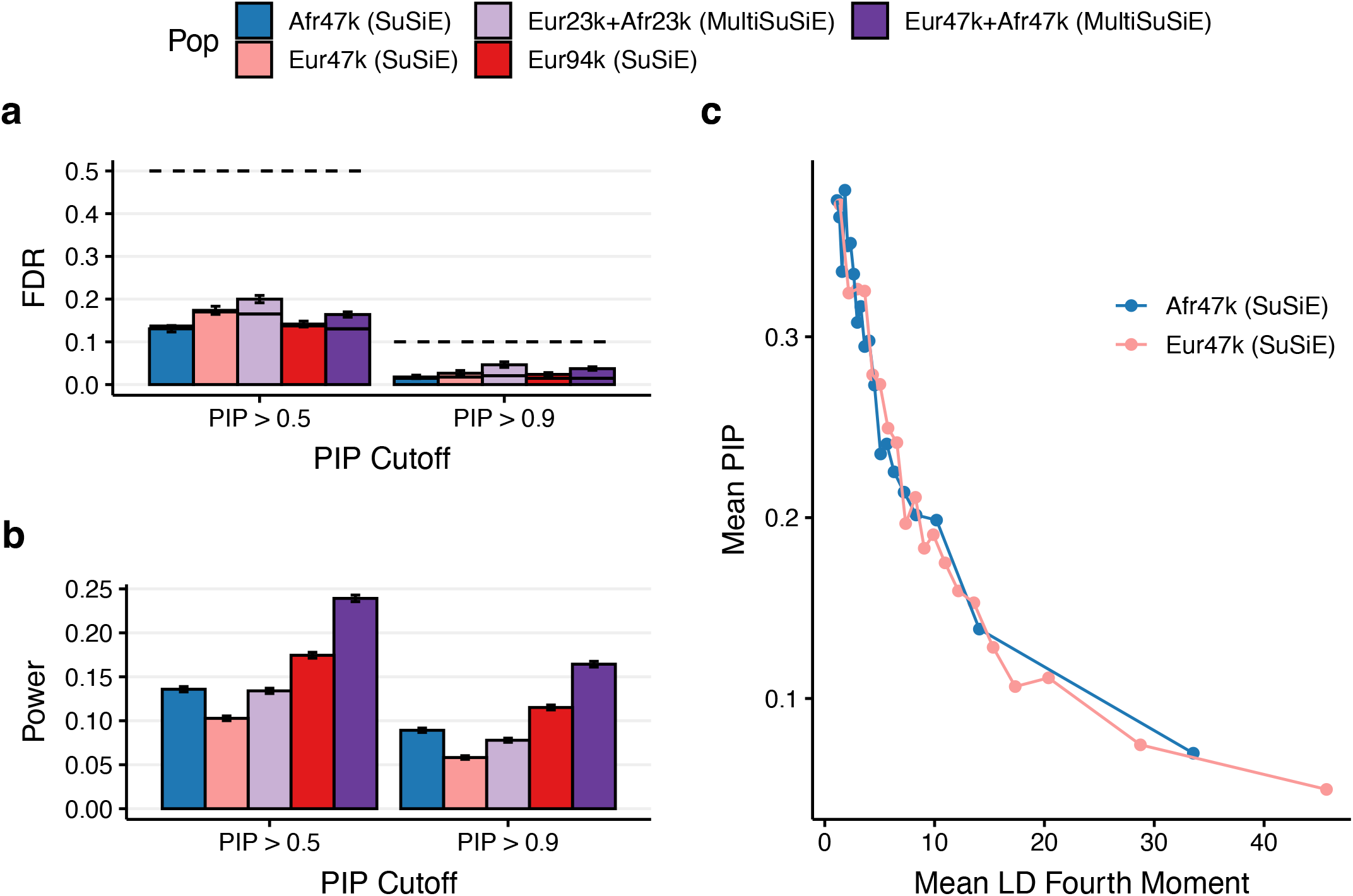
Simulation results across ancestries and sample sizes in simulations with variants selected to match MAF distributions between Afr47k and Eur47k. We report **(a)** the FDR (bars), conservative FDR upper bound (dashed line), and expected FDR (solid bars) at PIP > 0.5 and PIP > 0.9, **(b)** power at PIP > 0.5 and PIP > 0.9, **(c)** Mean PIP of causal variants with MAF > .05 within 20 equally sized LD 4^th^ moment bins in Afr47k and Eur47k fine-mapping. Error bars denote 95% Agresti-Coull binomial proportion confidence intervals.

**Supplementary Figure 19:**
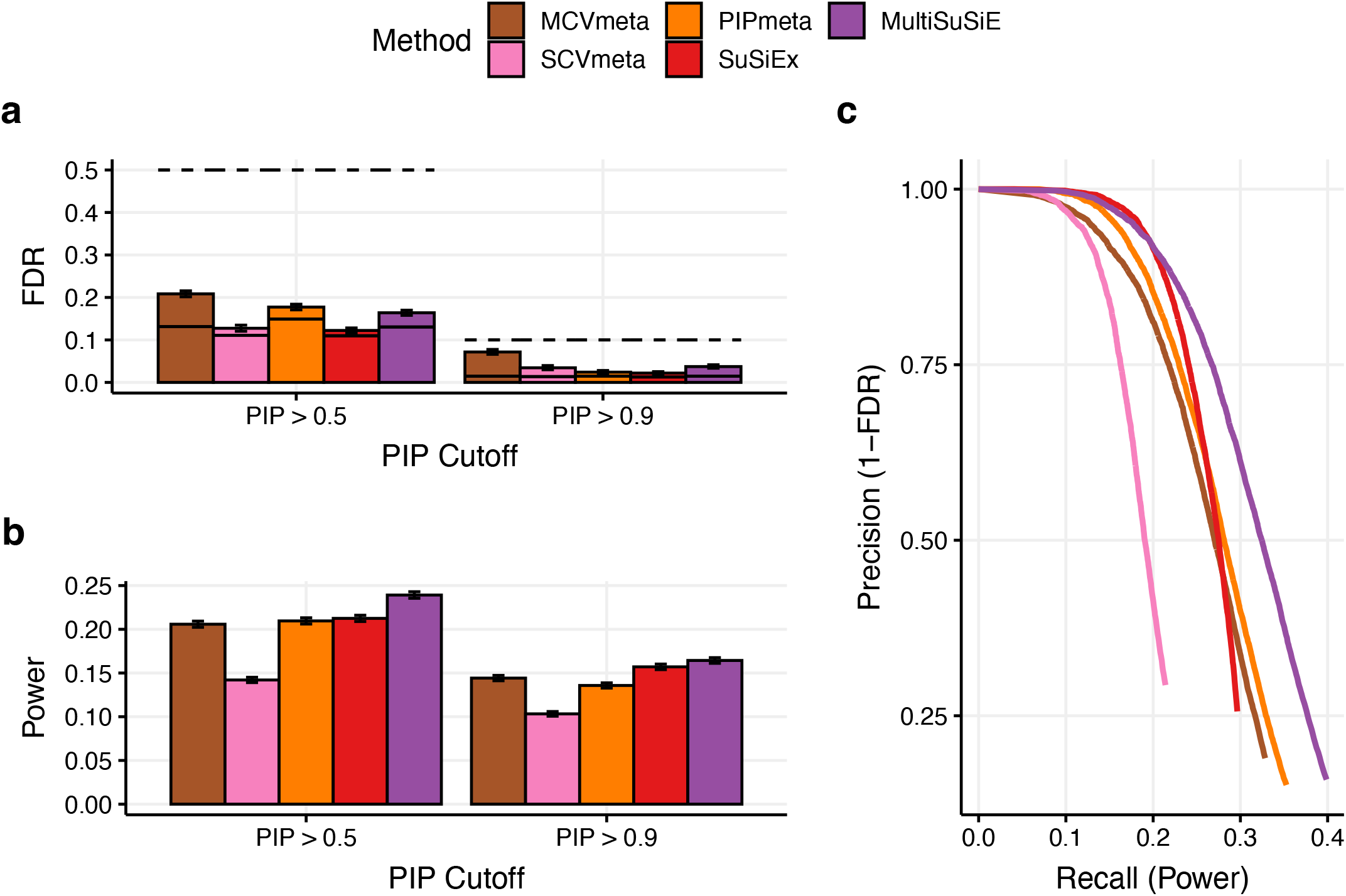
Simulation results for Afr47k+Eur47k across fine-mapping methods in simulations with variants selected to match MAF distributions between Afr47k and Eur47k. We report **(a)** the FDR (bars), conservative FDR upper bound (dashed line), and expected FDR (solid bars) at PIP > 0.5 and PIP > 0.9, **(b)** power at PIP > 0.5 and PIP > 0.9, **(c)** precision-recall curves varying PIP threshold from 1 to 0.01. Error bars denote 95% Agresti-Coull binomial proportion confidence intervals.

**Supplementary Figure 20:**
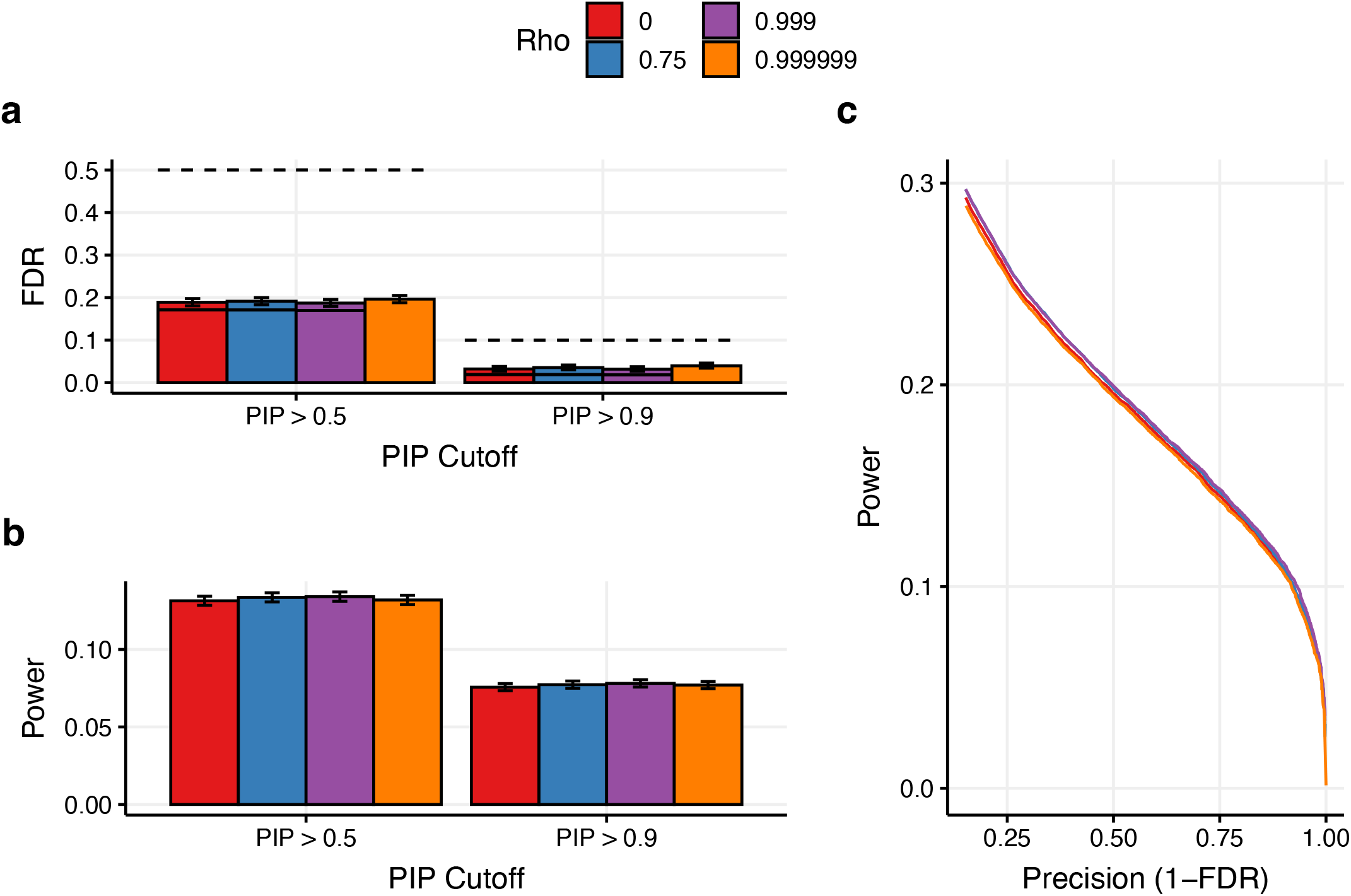
Simulation results for Afr47k+Eur47k using MultiSuSiE across values of the cross-ancestry correlation hyperparameter, *ρ. W*e report **(a)** the FDR (bars), conservative FDR upper bound (dashed line), and expected FDR (solid bars) at PIP > 0.5 and PIP > 0.9, **(b)** power at PIP > 0.5 and PIP > 0.9, **(c)** precision-recall curves varying PIP threshold. Error bars denote 95% Agresti-Coull binomial proportion confidence intervals.

**Supplementary Figure 21:**
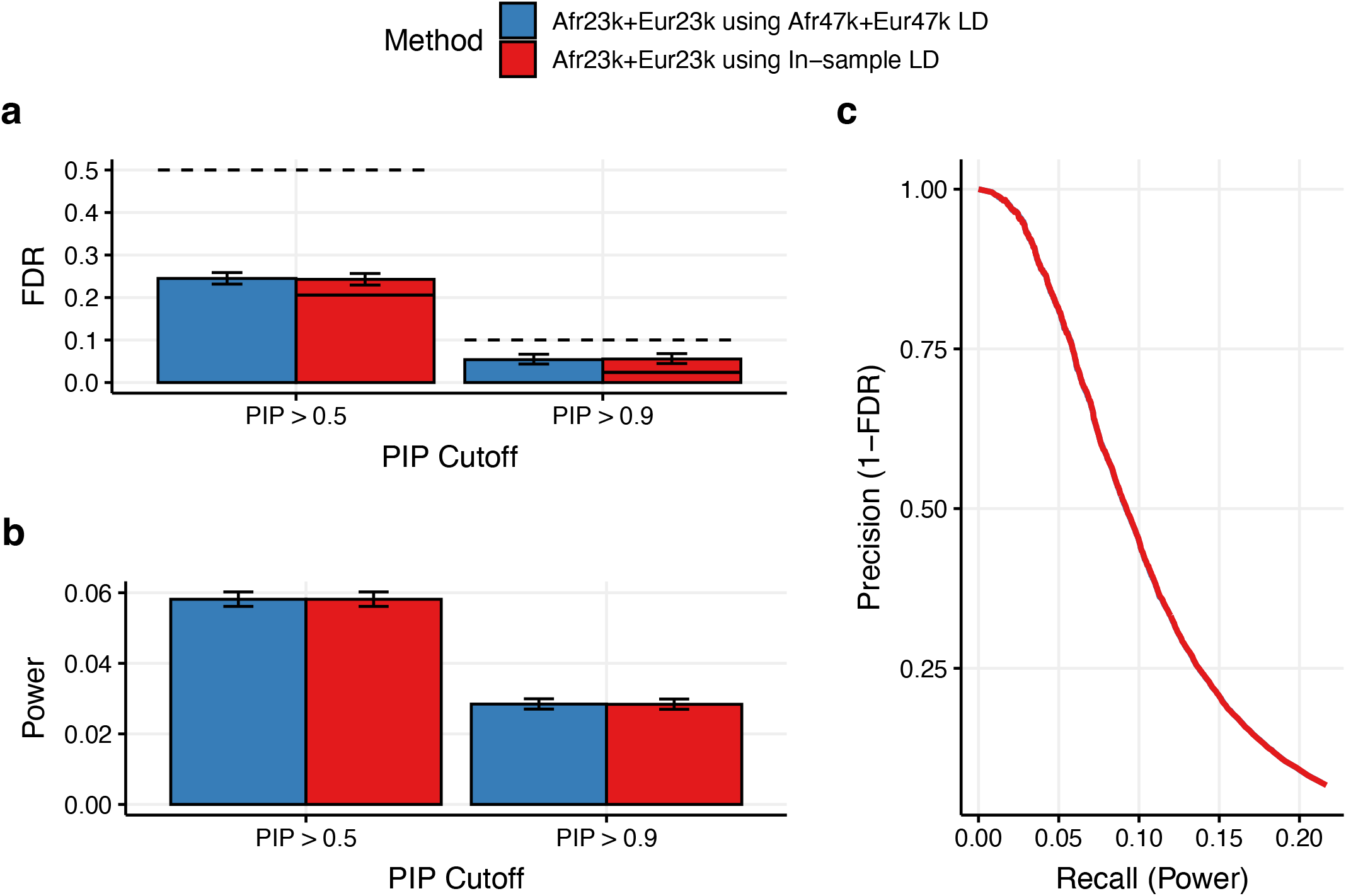
Simulation results for fine-mapping with Afr23k+Eur23k summary statistics and Afr47k+Eur47k LD using MultiSuSiE. We report **(a)** the FDR (bars), conservative FDR upper bound (dashed line), and expected FDR (solid bars) at PIP > 0.5 and PIP > 0.9, **(b)** power at PIP > 0.5 and PIP > 0.9, **(c)** precision-recall curves varying PIP threshold. Error bars denote 95% Agresti-Coull binomial proportion confidence intervals.

**Supplementary Figure 22:**
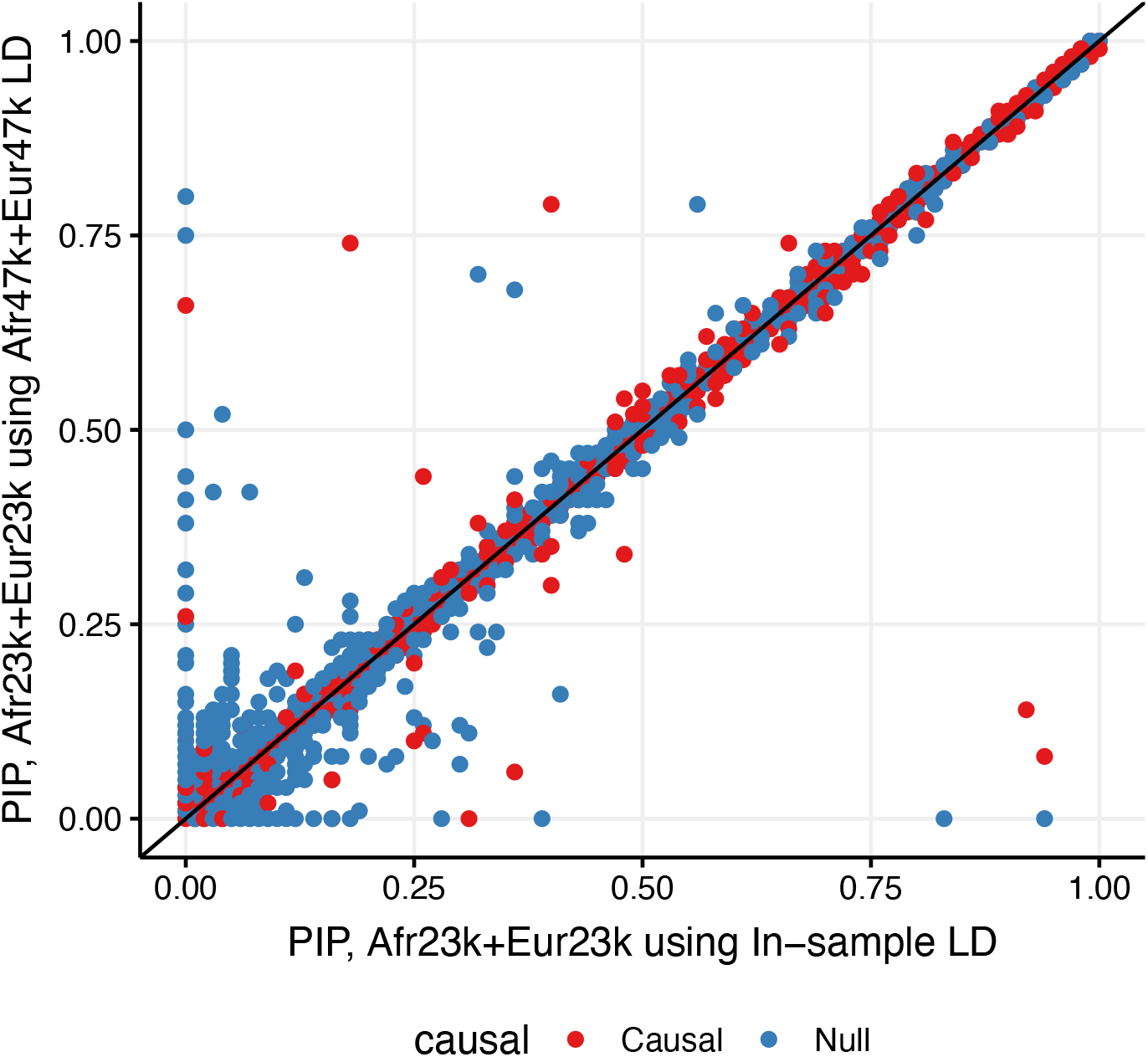
Variant-level simulation results for fine-mapping of Afr23k+Eur23k summary statistics with Afr47k+Eur47k LD using MultiSuSiE. Each point corresponds to a variant-simulation pair.

**Supplementary Figure 23:**
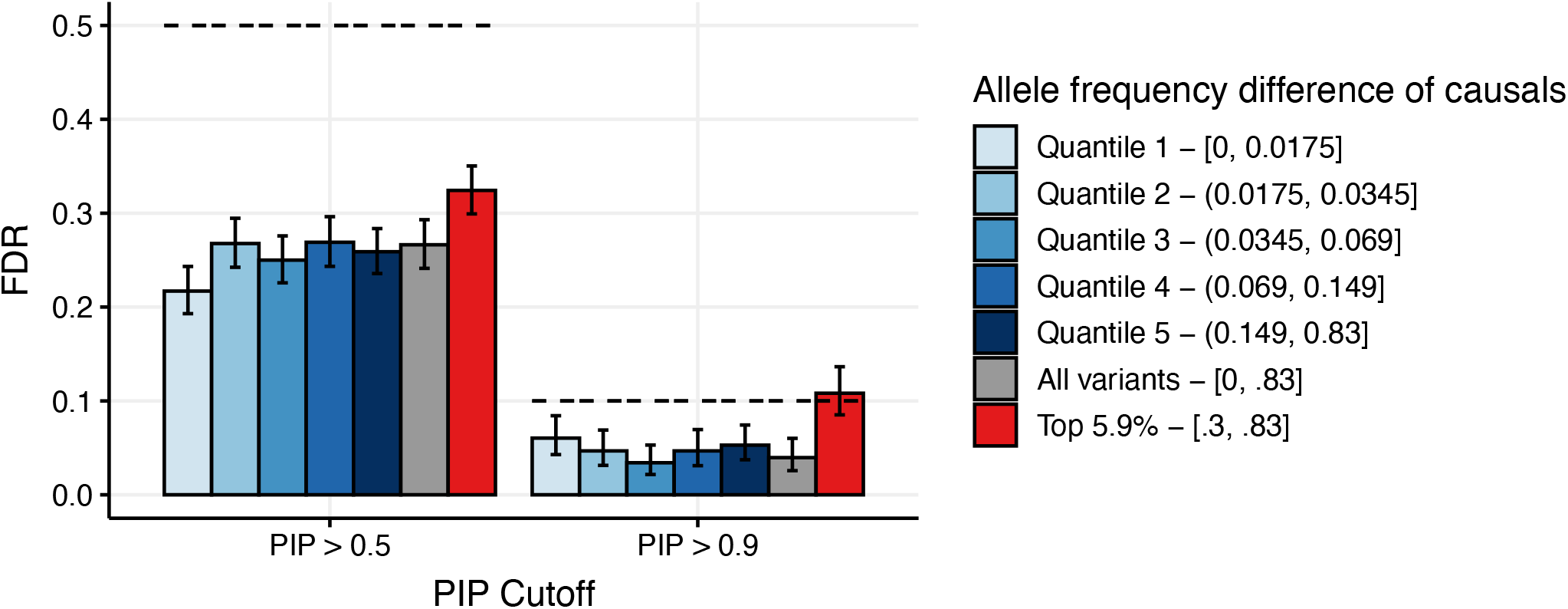
FDR of individual-level UKBioBank simulations varying the allele-frequency difference of causal variants. Simulations used individual-level imputed UKBioBank (UKBB) release 3 African (N=7k) and European (N=7k) ancestry genotypes. We directly simulated phenotypes using individual-level genotypes (rather than directly simulating summary statistics from in-sample LD) to ensure accurate modeling of long-range admixture LD. We restricted to variants with INFO score greater than 0.6 and UKBB MAF > 0.01 in either ancestry group. Individuals were assigned ancestry groups using UKB data-field 21000. Chromosome 11 was separated into 43 1Mb “causal windows” separated by 2Mb, each with a single, randomly selected causal variant. The central 1Mb of each 3Mb locus from the main simulations corresponded to a causal window. Causal variants were selected from variant sets stratified by absolute allele frequency difference between UKBioBank African and European ancestry individuals. Relative to main simulations, the per-variant heritability was increased by 47,401/7,000, so that the mean 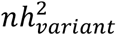 matched across the UKBB simulations and main simulations. As in the main simulations, cross-ancestry differences in effect sizes were due to differences in causal variant identify and per-allele effect sizes at shared causal variants. Phenotypes were simulated using a standard additive genetic model, *Y*_*k*_ *= X*_*k*_*β*_*k*_ *+ ϵ*_*k*_, where *β*_*k*_ *i*s a vector of true effect sizes for population *k*. The same loci used in the main simulations were fine-mapped across 1,000 simulated replicates. MultiSuSiE was run using GWAS summary statistics (adjusted for PC1), and in-sample LD (not adjusted for LD). We report the FDR (bars) and conservative FDR upper bound (dashed lines). Error bars denote 95% Agresti-Coull binomial proportion confidence intervals.

**Supplementary Figure 24:**
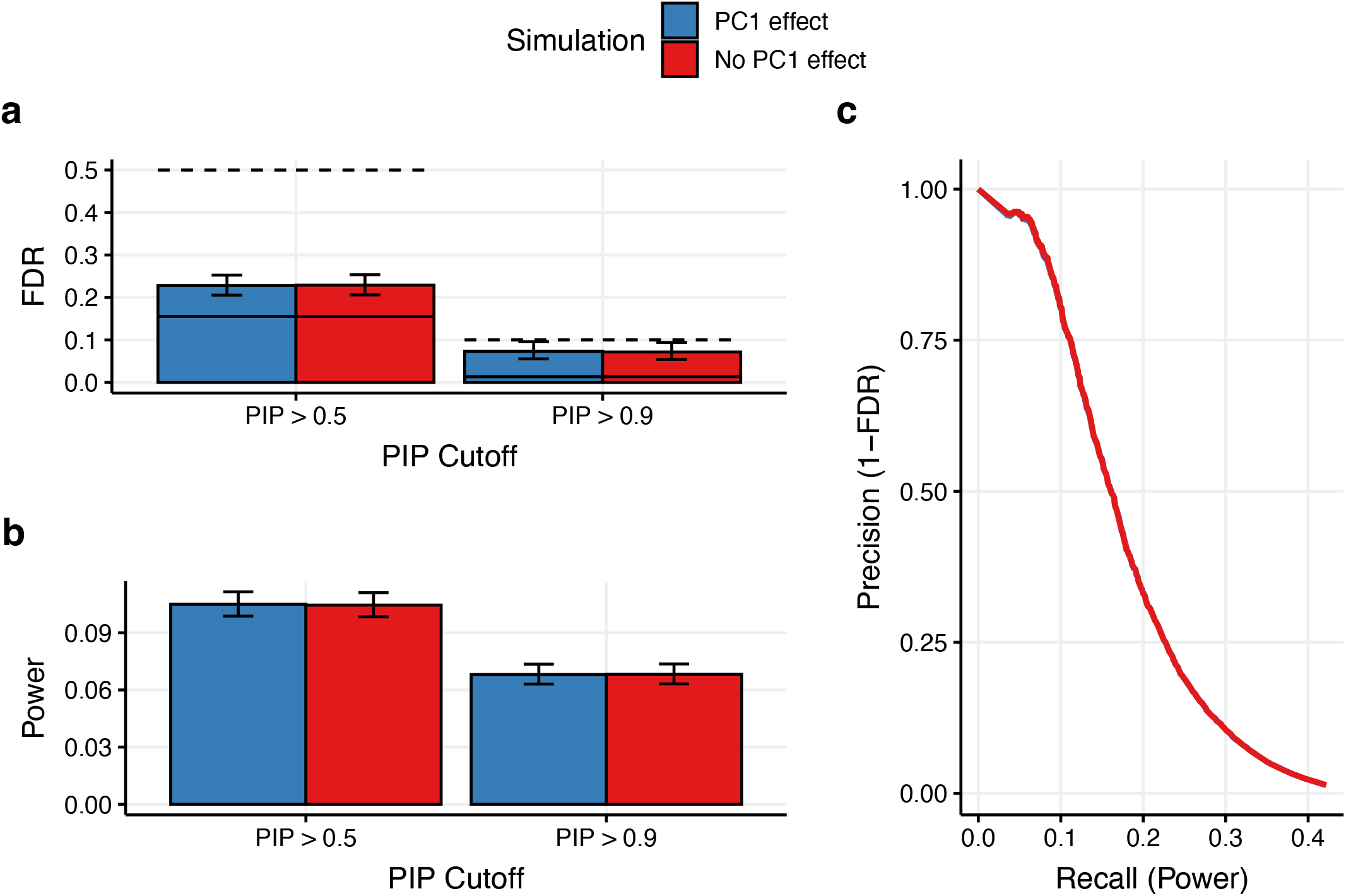
Summary of individual-level UKBioBank simulations with population stratification. We report **(a)** the FDR (bars), conservative FDR upper bound (dashed line), and expected FDR (solid bars) at PIP > 0.5 and PIP > 0.9, **(b)** power at PIP > 0.5 and PIP > 0.9, **(c)** precision-recall curves varying PIP threshold from 1 to 0.01. Error bars denote 95% Agresti-Coull binomial proportion confidence intervals. Simulations used individual-level imputed UKBioBank (UKBB) release 3 African (N=7k) and European (N=7k) ancestry genotypes. Simulations were identical to those described in Supplementary Figure 23 with the following exceptions: (1) we simulated causal variants in the central 1Mb of each 3Mb window used in the main simulations, not in the 43 causal windows used for Supplementary Figure 23, (2) we simulated 5 causal variants per window on average, (3) we did not select causal variants based on absolute allele frequency difference, (4) we simulated phenotypes as *Y*_*k*_ *= X*_*k*_*β*_*k*_ *+ ϵ*_*k*_ *+ PC*1_*k*_ *∗ β*_*PC*1_, where *β*_*k*_ *i*s a vector of true effect sizes for population *k, PC*1 *i*s a vector containing the values of the first principle component for population *k*, and *β*_*PC*1_ *i*s set so that *Var*(*PC*1 *∗ β*_*PC*1_) *= Var*(*ϵ*_*k*_) in simulations with PC1 effects (blue) and set to 0 in simulations without PC1 effects (red).

**Supplementary Figure 25:**
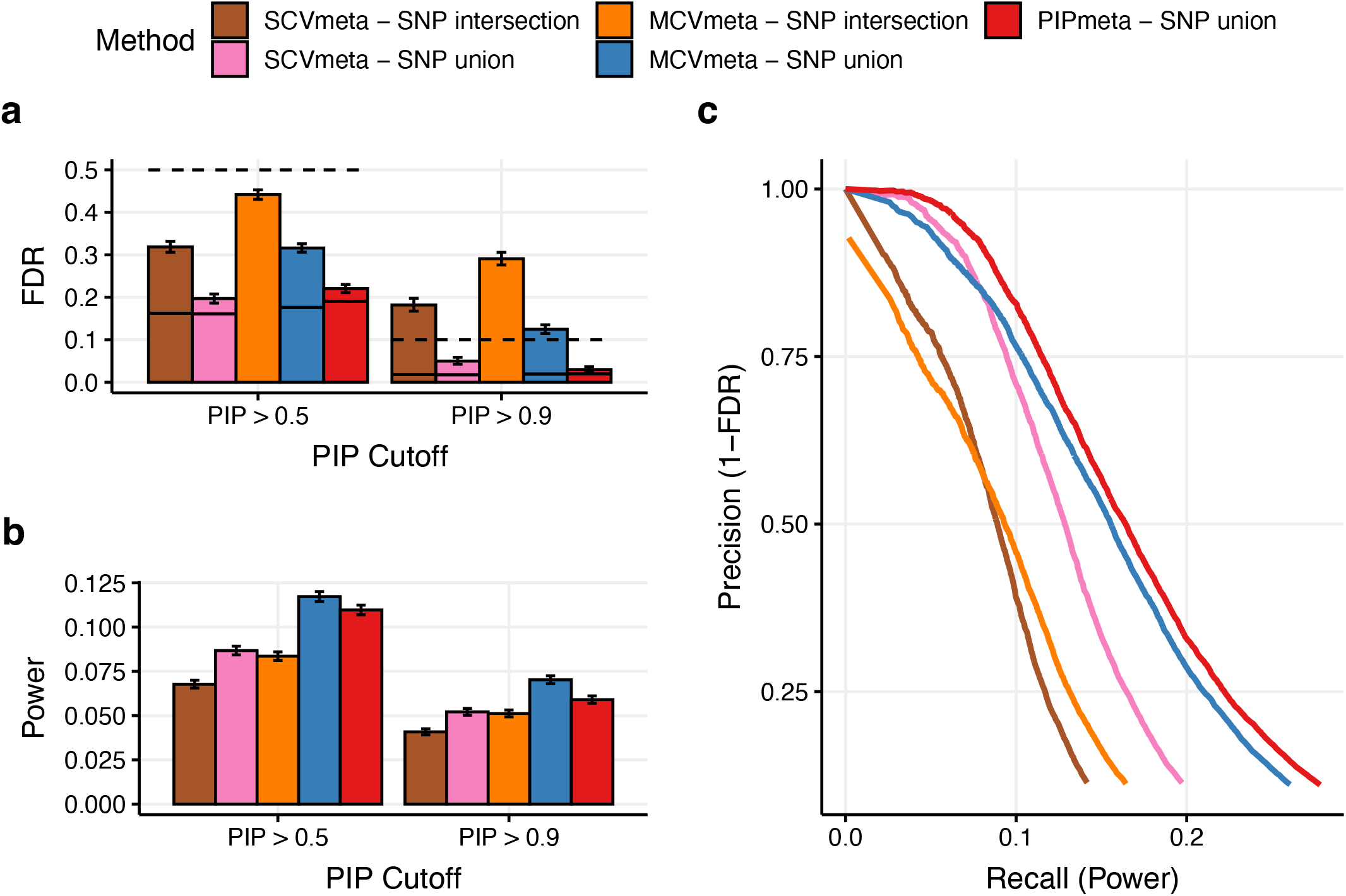
Simulation results for Afr47k+Eur47k using meta-analysis methods, restricting to variants with MAF > 0.01 in both populations. *W*e report **(a)** the FDR (bars), conservative FDR upper bound (dashed line), and expected FDR (solid bars) at PIP > 0.5 and PIP > 0.9, **(b)** power at PIP > 0.5 and PIP > 0.9, **(c)** precision-recall curves varying PIP threshold from 1 to .01. Methods labeled with SNP intersection restrict to variants with MAF > 0.01 in both ancestries. Methods labeled with SNP union include variants with MAF > 0.01 in at least one ancestry. Error bars denote 95% Agresti-Coull binomial proportion confidence intervals.

**Supplementary Figure 26:**
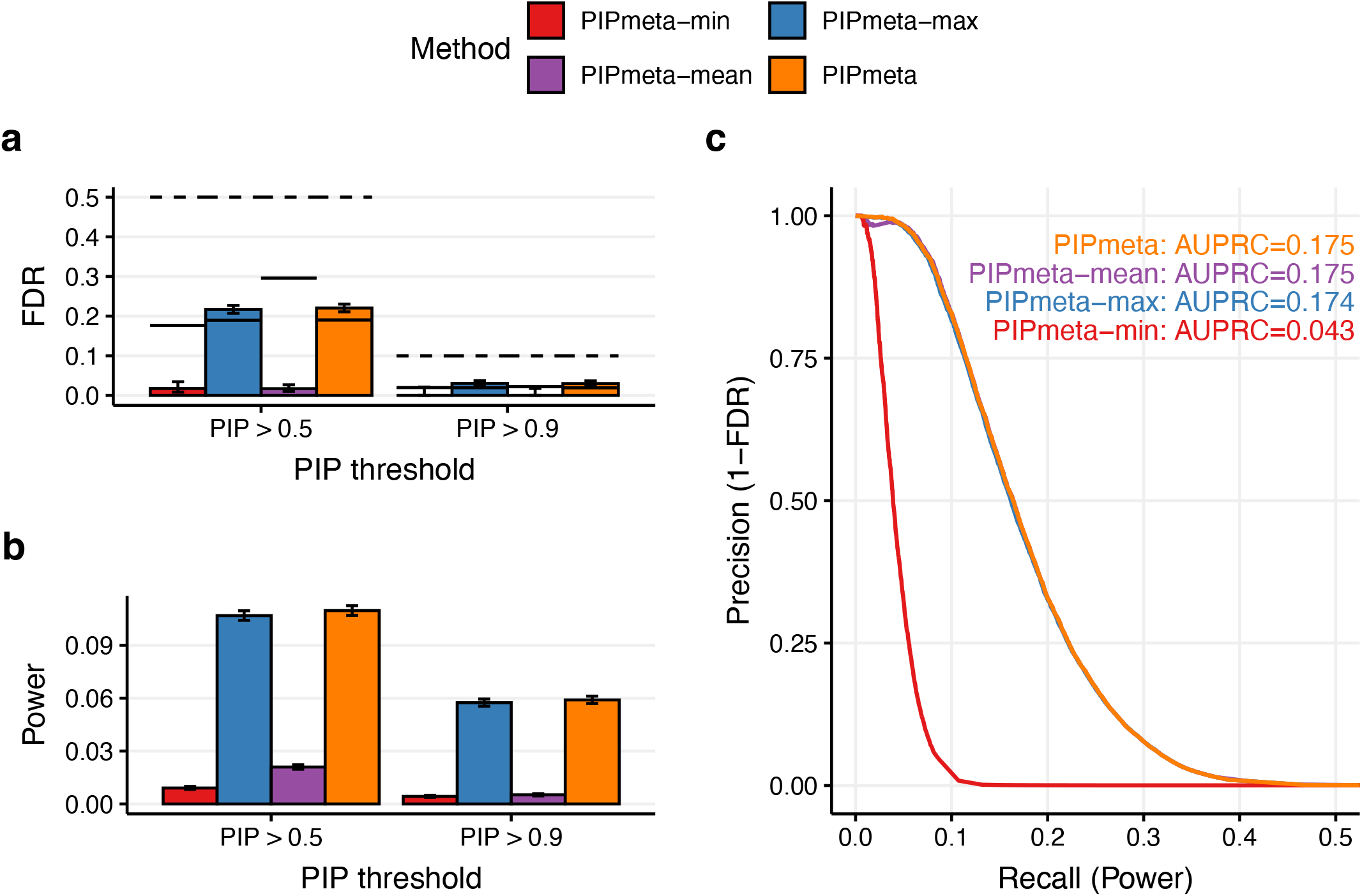
Simulation results for Afr47k+Eur47k using post-hoc meta-analysis methods, varying method for combining PIPs across ancestries. *W*e report **(a)** the FDR (bars), conservative FDR upper bound (dashed line), and expected FDR (solid bars) at PIP >0.5 and PIP > 0.9, **(b)** power at PIP > 0.5 and PIP > 0.9, **(c)** precision-recall curves varying PIP threshold. Error bars denote 95% Agresti-Coull binomial proportion confidence intervals. PIPmeta-mean uses the mean PIP across ancestries. PIPmeta-max uses the maximum PIP across ancestries. PIPmeta-min uses the minimum PIP across ancestries.

**Supplementary Figure 27:**
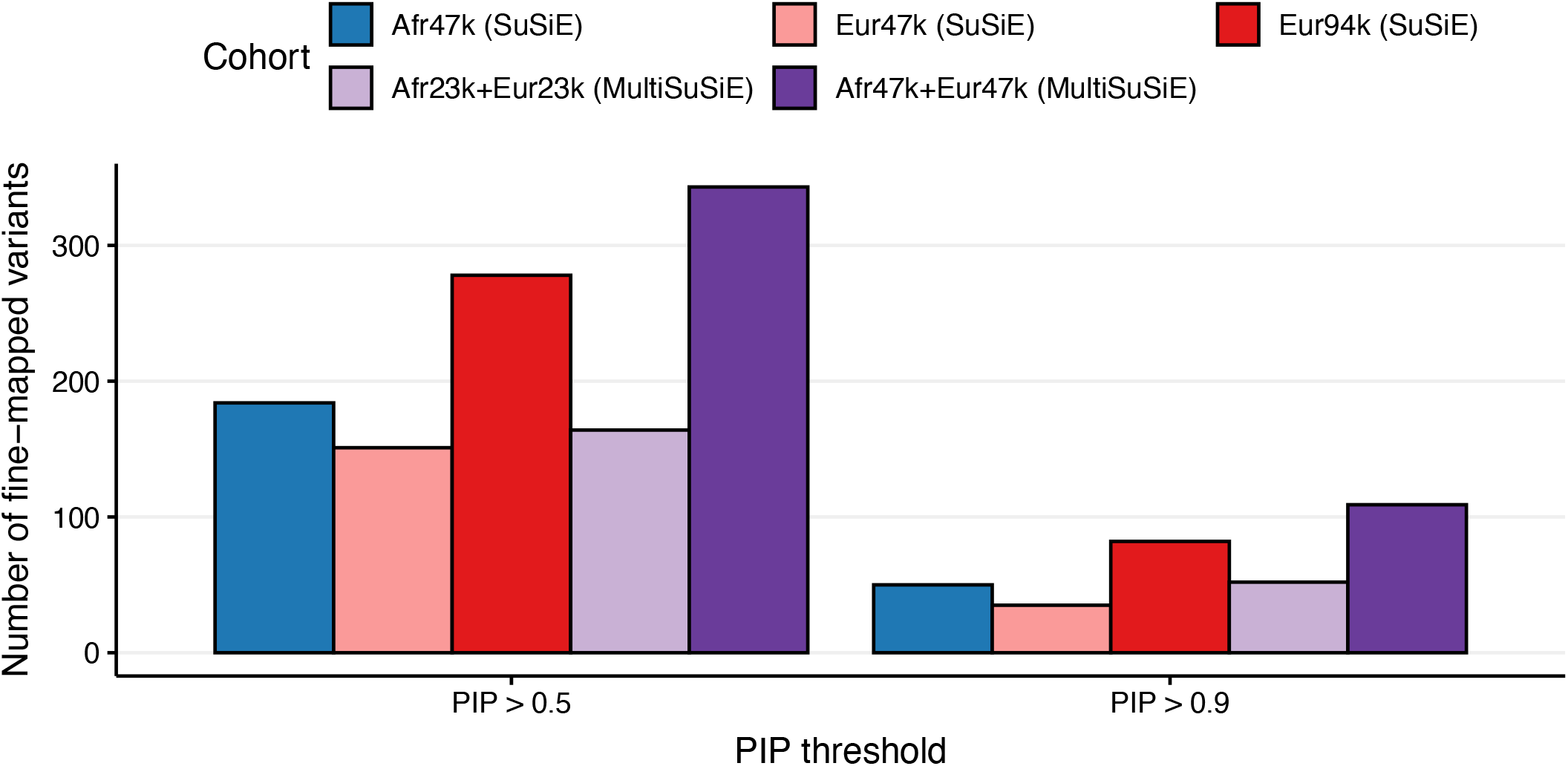
Real trait results across ancestries and sample sizes analyzed, restricting each cohort to loci that contain a variant with *p*<5*10^−6^. We report the number of fine-mapped variants found with a cohort.

**Supplementary Figure 28:**
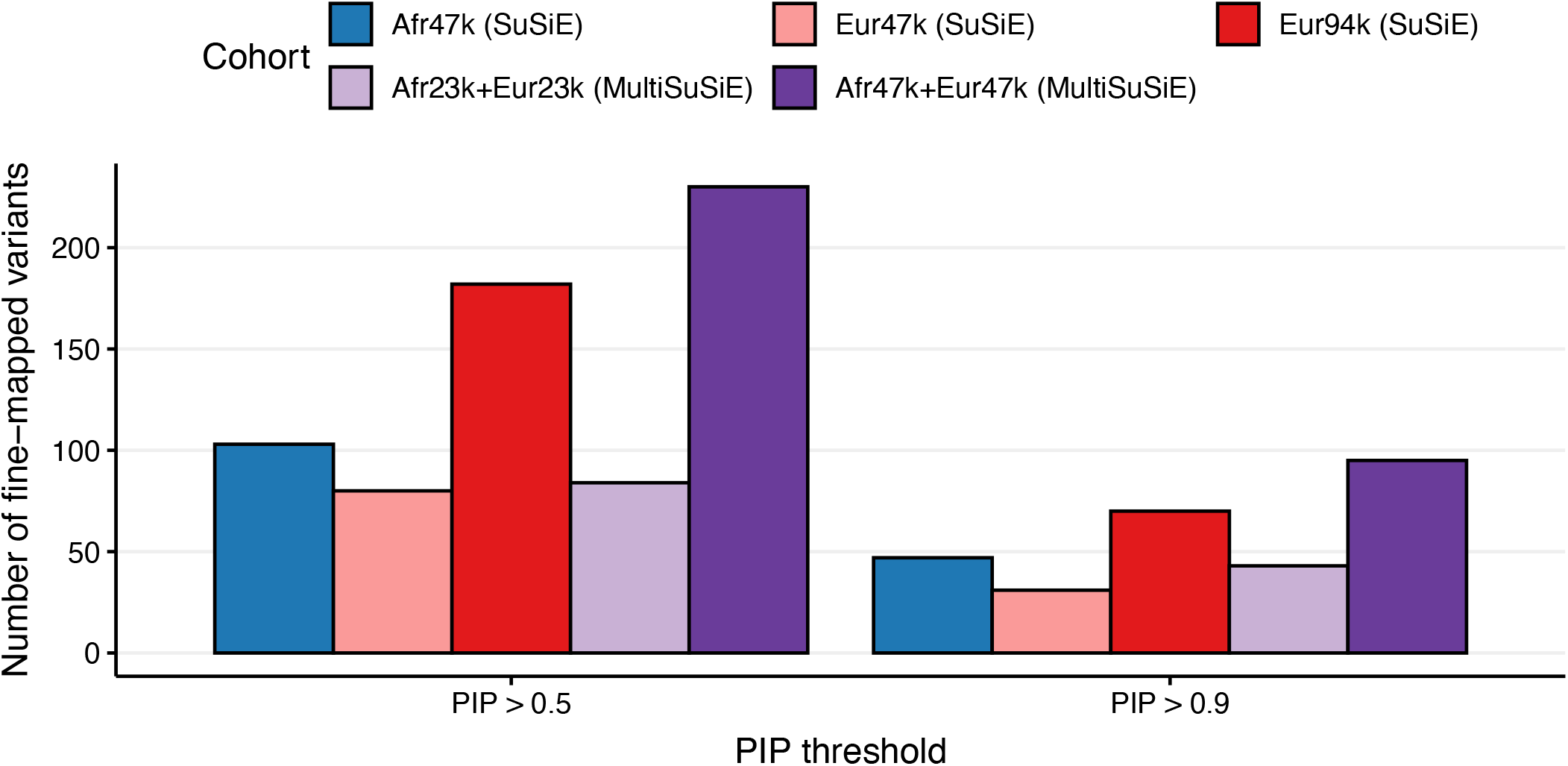
Real trait results across ancestries and sample sizes analyzed, restricting each cohort to loci that contain a genome-wide significant variant (*p*<5*10^−8^). We report the number of fine-mapped variants found with a cohort.

**Supplementary Figure 29:**
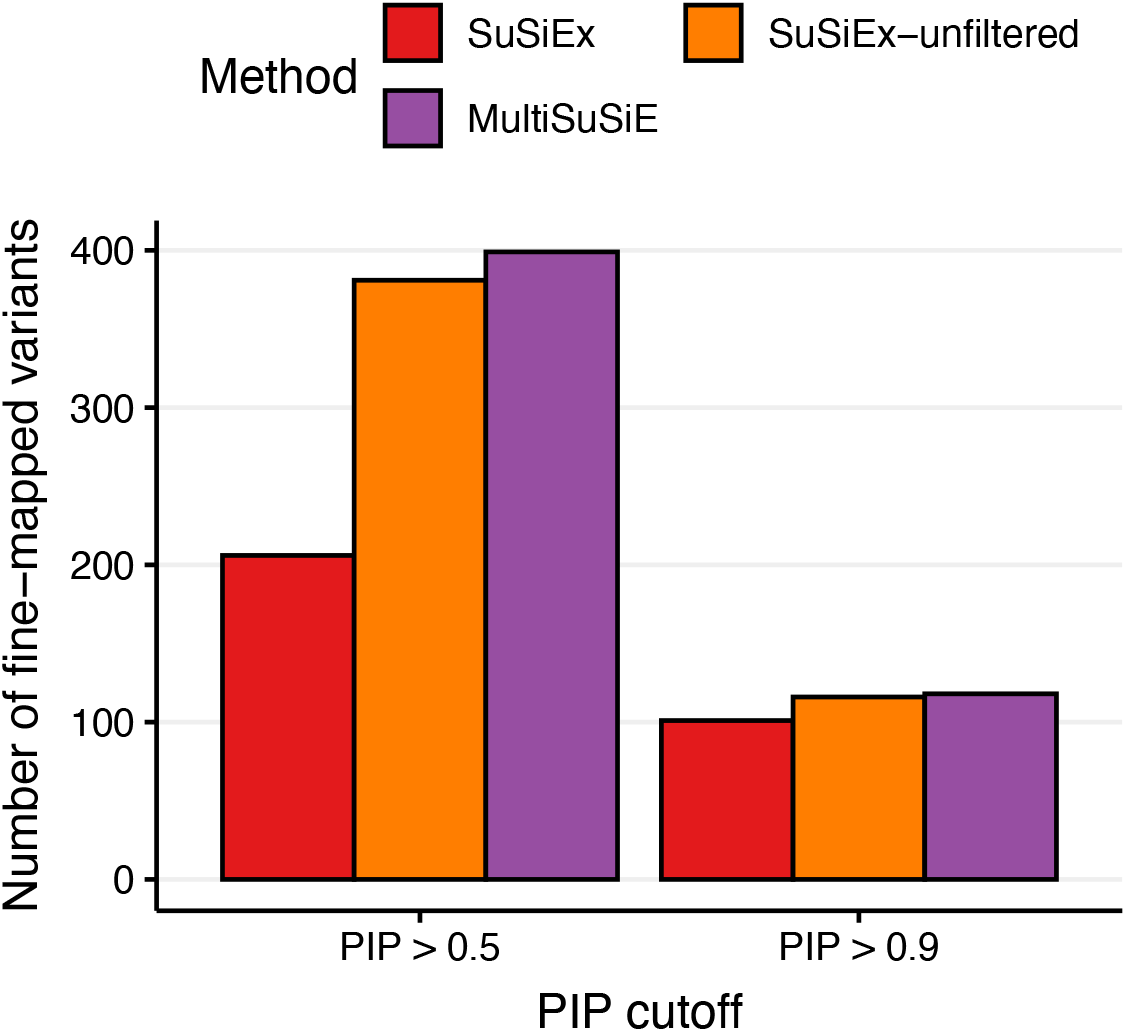
Real trait results for Afr47k+Eur47k across fine-mapping methods including SuSiEx-unfiltered. We report the number of variant-trait pairs with PIP > 0.5 or 0.9 across 14 uncorrelated traits using Afr47k+Eur47k.

**Supplementary Figure 30:**
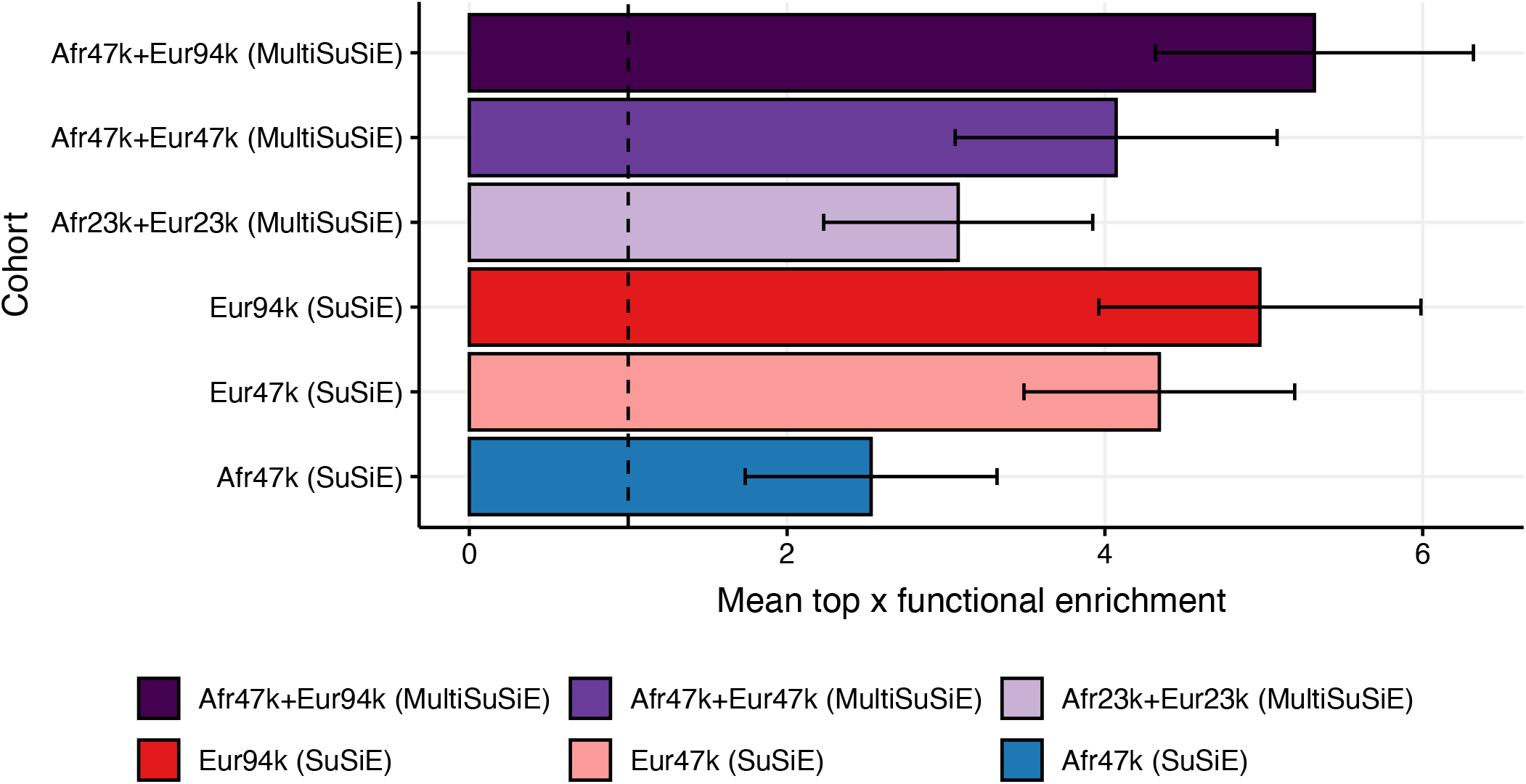
Functional enrichment of fine-mapped variants across ancestries and sample sizes, controlling for power by restricting to the top 118 variants for each cohort. We report mean top x functional enrichment (*P*(*a*_*i*_ *= 1*|*PIP*_*i*_ *≥ PIP*_(*P–x–*1)_)*/P*(*a*_*i*_ *=* 1), where *PIP*_(*p*)_ is the *p*-th smallest PIP for a cohort or method and *P* is the number of variants, *x*=118 because all methods identified at least 118 variants with PIP > 0.5) averaged over 11 functional annotations. Error bars denote 95% confidence intervals based on a genomic block-jackknife with 200 blocks. The vertical dashed bar denotes no functional enrichment.

**Supplementary Figure 31:**
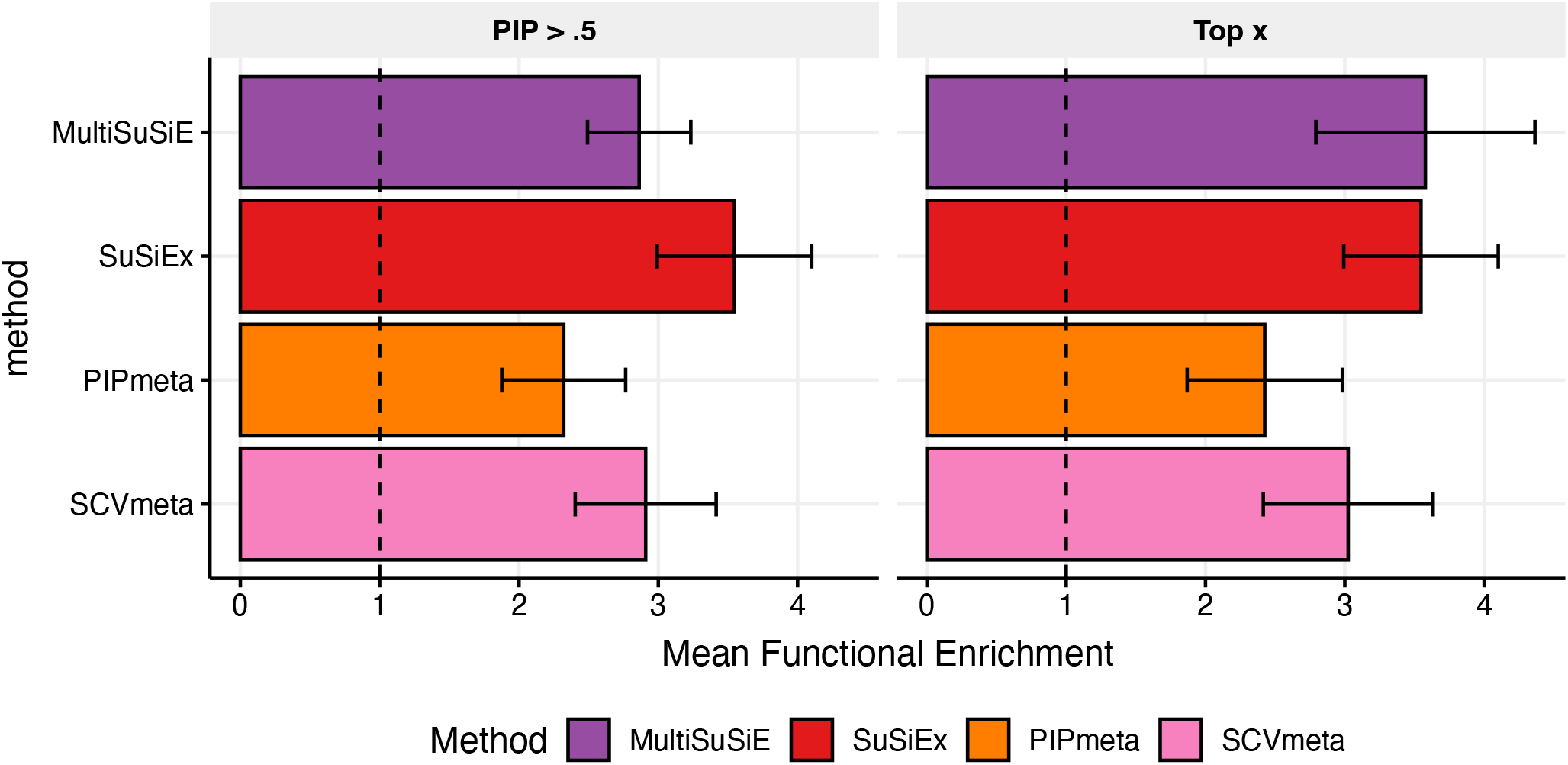
Functional enrichment of fine-mapped variants for Afr47k+Eur47k across fine-mapping method (rows) and enrichment metric (columns). We report mean functional enrichment (*P*(*a*_*i*_ *= 1*|*PIP*_*i*_ *>* .5)*/P*(*a*_*i*_ *= 1*), where *ai* equals 1 if variant *i* is in annotation *a* and 0 otherwise) and top-206 functional enrichment (*P*(*a*_*i*_ *= 1*|*PIP*_*i*_ *≥ PIP*_(*M–x–*1)_)*/P*(*a*_*i*_ *= 1*), where *PIP*_(*p*)_ *i*s the *p*-th smallest PIP for a cohort or method, and *M* is the number of variants, *x*=206 because all methods identified at least 206 variants with PIP > 0.5), averaged over 11 functional annotations. Error bars denote 95% confidence intervals based on a genomic block-jackknife with 200 blocks. The vertical dashed bar denotes no functional enrichment.

**Supplementary Figure 32:**
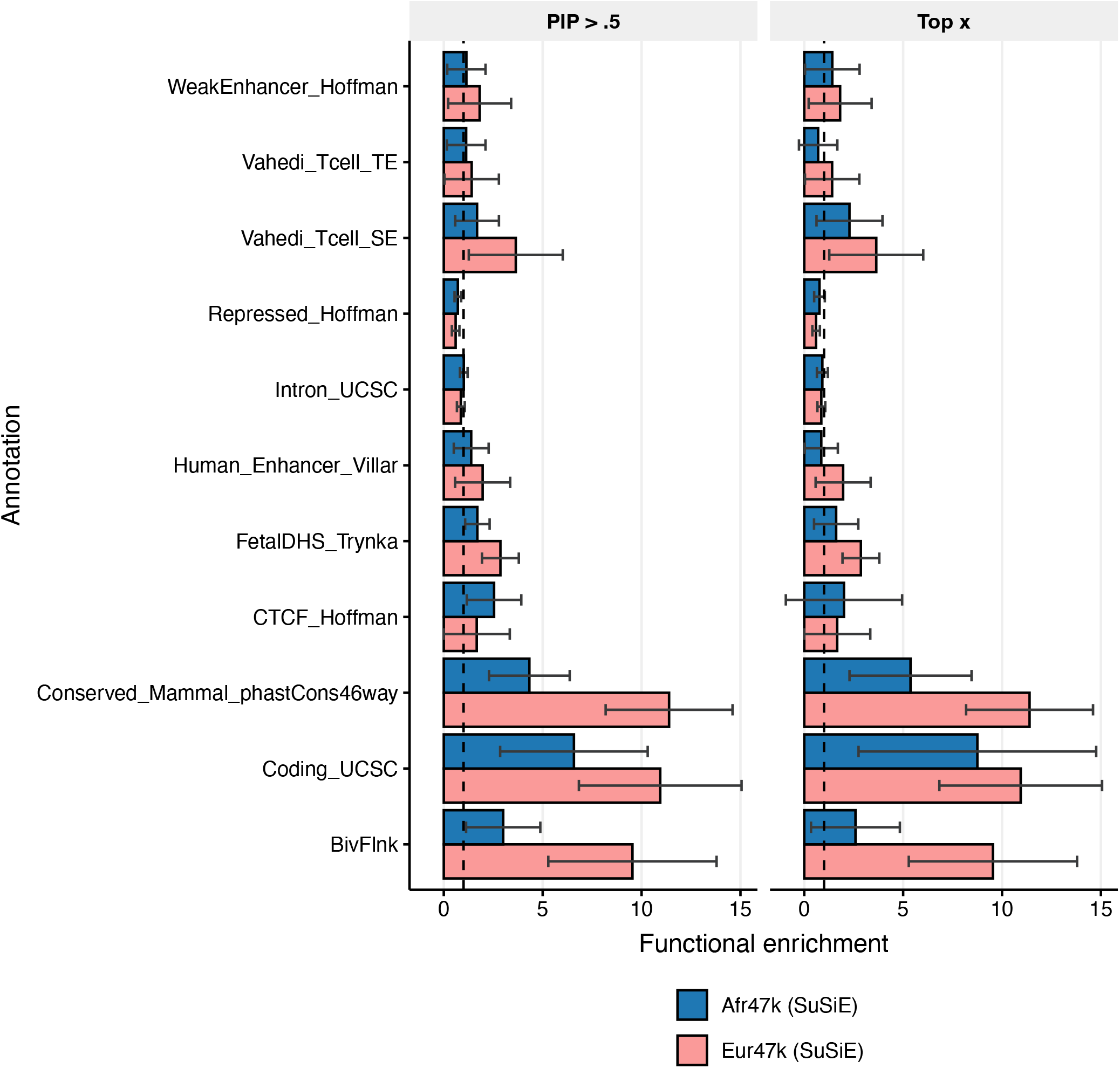
Functional enrichment of fine-mapped variants of Afr47k and Eur47k, separated by annotation. We report functional enrichment (*P*(*a*_*i*_ *= 1*|*PIP*_*i*_ *>* .5)*/P*(*a*_*i*_ *= 1*), where *ai* equals 1 if variant *i* is in annotation *a* and 0 otherwise) and top-206 functional enrichment (*P*(*a*_*i*_ *= 1*|*PIP*_*i*_ *≥ PIP*_(*M–x–*1)_)*/P*(*a*_*i*_ *= 1*), where *PIP*_(*p*)_ *i*s the *p*-th smallest PIP for a cohort or method, and *M* is the number of variants, *x*=118 because all cohorts identified at least 118 variants with PIP > 0.5). Error bars denote 95% confidence intervals based on a genomic block-jackknife with 200 blocks. The vertical dashed bar denotes no functional enrichment.

**Supplementary Figure 33:**
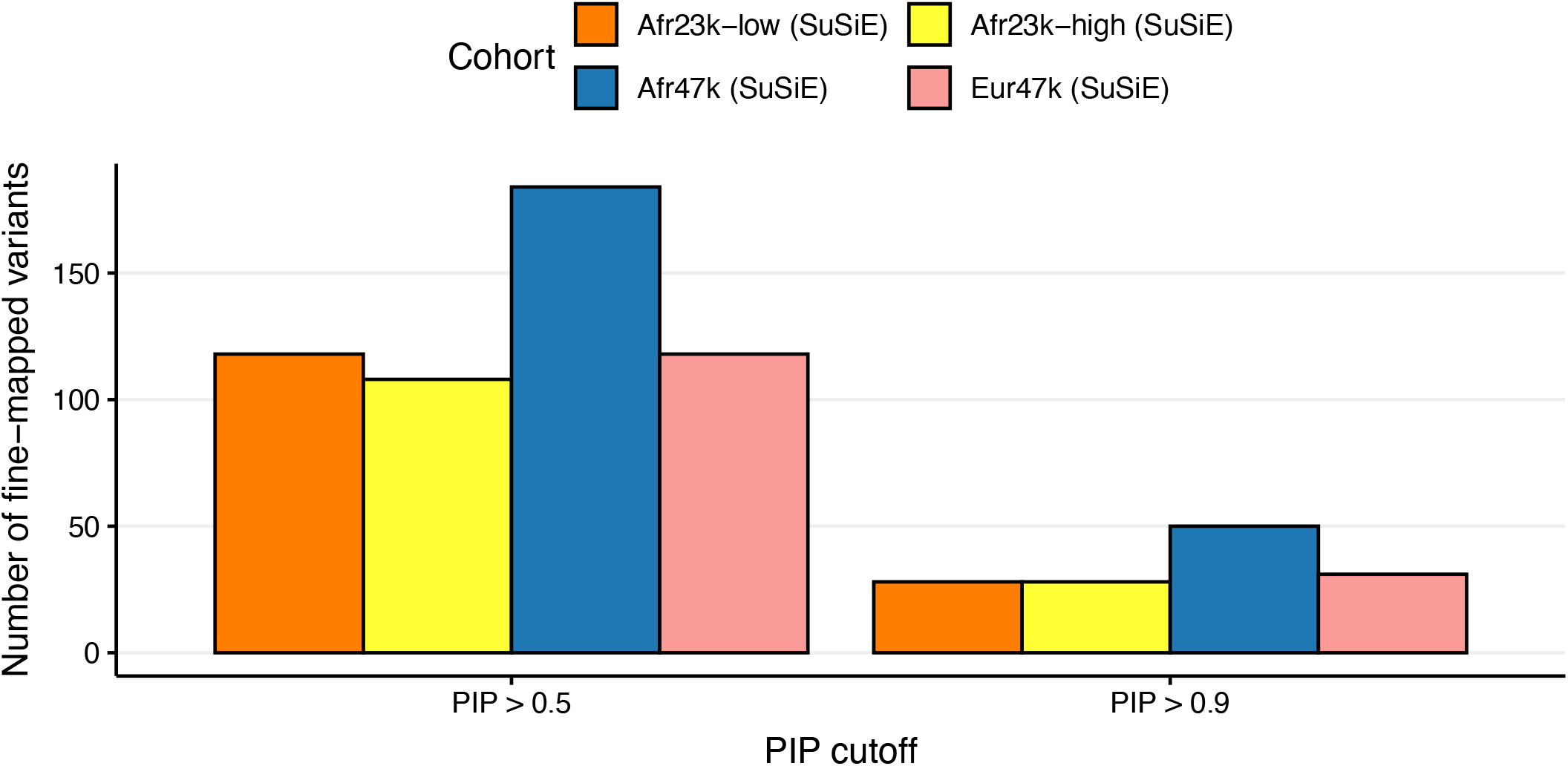
Real trait results across ancestries and sample sizes. We report the number of variant-trait pairs fine-mapped by MultiSuSiE (for multi-ancestry cohorts) or SuSiE (for single-ancestry cohorts) at PIP > 0.5 and PIP > 0.9 across 14 quantitative traits.

**Supplementary Figure 34:**
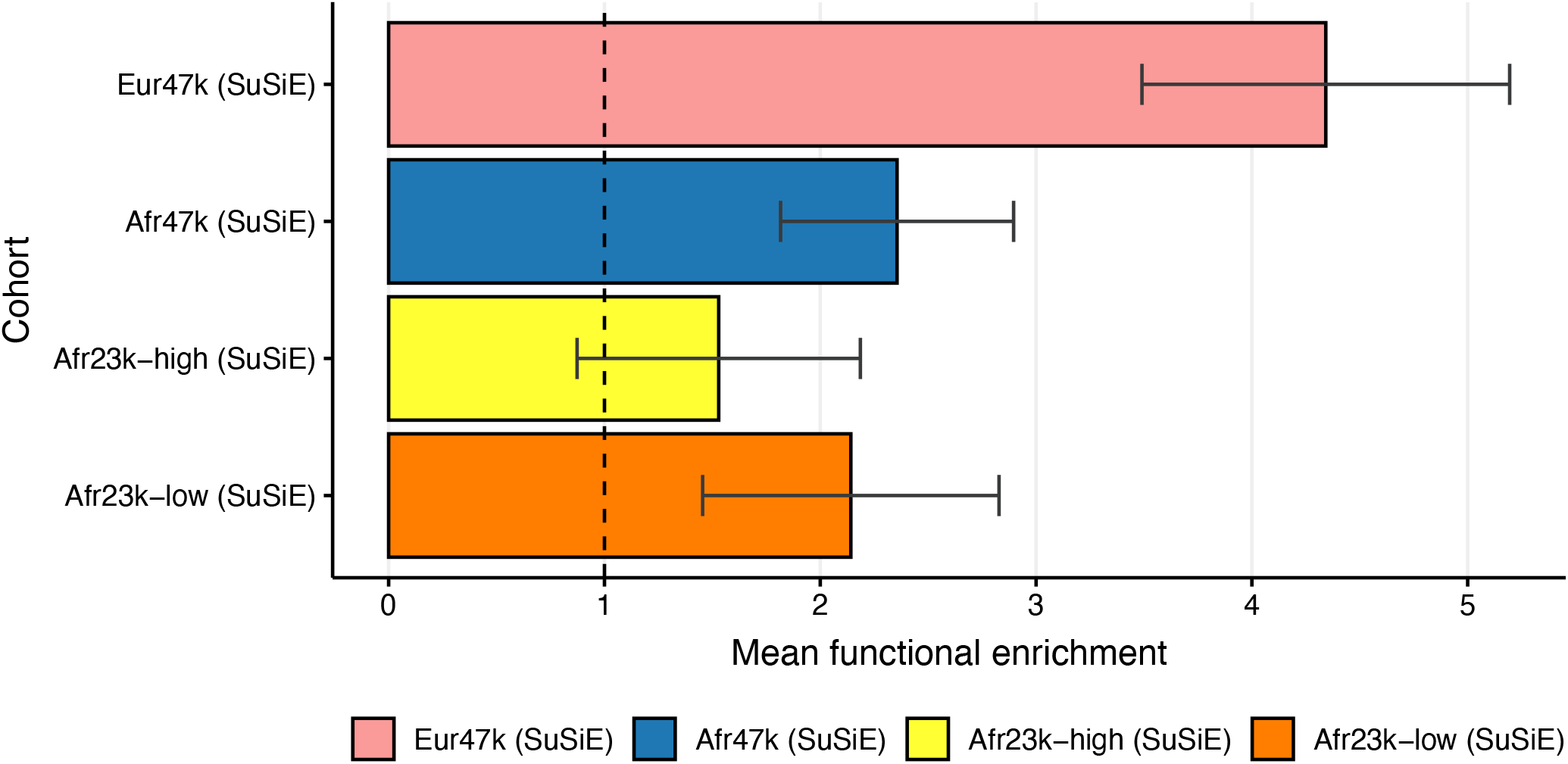
Functional enrichment of fine-mapped variants across ancestries and admixture proportions. We report mean functional enrichment of fine-mapped variants (*P*(*a*_*i*_ *= 1*|*PIP*_*i*_ *> 0*.5)*/P*(*a*_*i*_ *= 1*), where *ai* equals 1 if variant *i* is in annotation *a* and 0 otherwise), averaged across 11 functional annotations. Error bars denote 95% confidence intervals based on a genomic block-jackknife with 200 blocks. The vertical dashed bar denotes no functional enrichment. The vertical dashed bar corresponds to no functional enrichment.

**Supplementary Figure 35:**
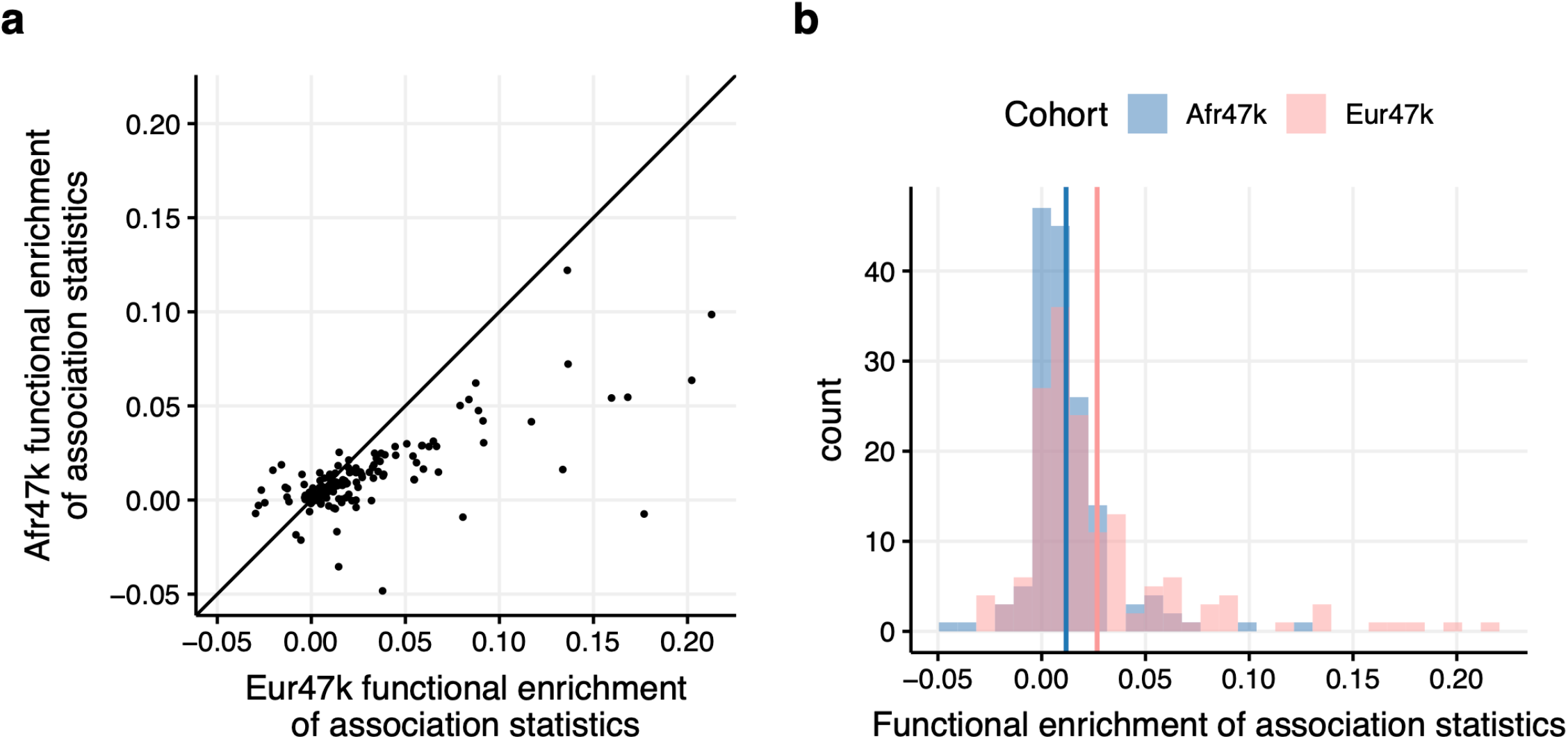
Functional enrichment of association statistics in Afr47k and Eur47k. Each point in (a) and count in (b) corresponds to an approximately independent trait-annotation pair. In (a), the black line has slope of 1 and intercept of 0. In (b), the blue and pink lines correspond to the mean functional enrichment of association statistics in Afr47k and Eur47k, respectively. functional enrichment of association statistics is defined as 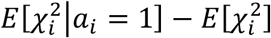 where 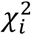 is the marginal GWAS chi-squared statistic for variant *i* and *a*_*i*_ equals 1 if variant *i* is in annotation *a* and 0 otherwise.

**Supplementary Figure 36:**
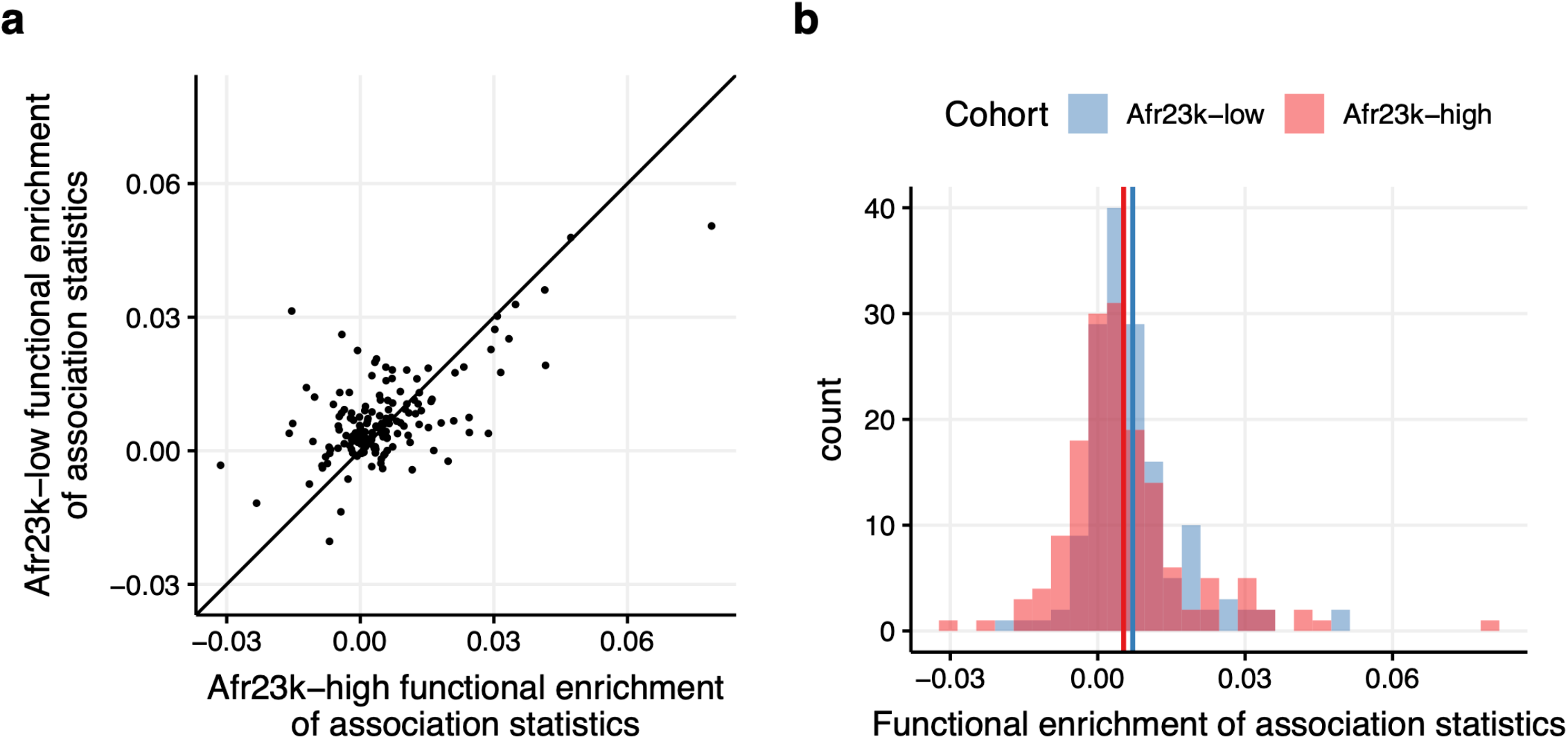
Functional enrichment of association statistics in Afr23k-low and Afr23k-high. Each point in (a) and count in (b) corresponds to an approximately independent trait-annotation pair. In (a), the black line has slope of 1 and intercept of 0. In (b), the blue and red lines correspond to the mean functional enrichment of association statistics in Afr23k-low and Afr23k-high, respectively. Functional enrichment of association statistics is defined as 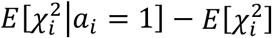 where 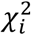 is the marginal GWAS chi-squared statistic for variant *i* and *a*_*i*_ equals 1 if variant *i* is in annotation *a* and 0 otherwise.

## Supplementary Tables

**Supplementary Table 1:** Summary of loci used in simulations. We report the start and end coordinates of each locus as well as the start and end of each causal window.

| chromosome | locus_start | locus_end | causal_window_start | causal_window_end | n_variants |
| --- | --- | --- | --- | --- | --- |
| 11 | 1 | 3000000 | 1000000 | 2000000 | 21054 |
| 11 | 6000001 | 9000000 | 7000000 | 8000000 | 23634 |
| 11 | 12000001 | 15000000 | 13000000 | 14000000 | 19729 |
| 11 | 18000001 | 21000000 | 19000000 | 20000000 | 23215 |
| 11 | 30000001 | 33000000 | 31000000 | 32000000 | 17262 |
| 11 | 36000001 | 39000000 | 37000000 | 38000000 | 22029 |
| 11 | 60000001 | 63000000 | 61000000 | 62000000 | 18398 |
| 11 | 66000001 | 69000000 | 67000000 | 68000000 | 17561 |
| 11 | 72000001 | 75000000 | 73000000 | 74000000 | 17789 |
| 11 | 78000001 | 81000000 | 79000000 | 80000000 | 22479 |

**Supplementary Table 2: FDR and power across ancestries and sample sizes analyzed**. We report the FDR and power at PIP > 0.5 and PIP > 0.9.(see Excel file)

**Supplementary Table 3: Mean PIP and mean LD 4**^**th**^ **moment of common causal variant bins for Eur47k and Afr47k**. We report statistics for LD 4^th^ moment bins. For each ancestry, we stratified causal variants with MAF > 0.05 into 20 equally sized bins based on LD 4^th^ moments. (see Excel file)

**Supplementary Table 4: LD fourth moments explain the effect of ancestry on causal PIP**. We report fit statistics for linear regressions on causal variant PIP varying explanatory variables used. We pooled variants from SuSiE fine-mapping simulation results for Afr47k and Eur47k, restricting to causal variants with ancestry-specific MAF > .05. We regressed causal variant PIPs on population labels (univariate regression) or both population labels and log-transformed LD fourth moments (bivariate regression). Population labels had a highly significant effect on causal variant PIPs in the univariate regression (*p*_*population*_<2*10^−16^) but an insignificant effect on causal variant PIPs in the bivariate regression (*p*_*population*_=0.16, *p*_*log Id fourth moment*_ <2*10^−16^)). We report the estimates of each parameter from 10 linear models. P-values less than 2*10^−16^ are reported as 2*10^−16^.(see Excel file)

**Supplementary Table 5: FDR and power for Afr47k+Eur47k across fine-mapping methods**. We report the FDR and power at PIP > 0.5 and PIP > 0.9. (see Excel file)

**Supplementary Table 6: Precision and recall values for Afr47k+Eur47k across fine-mapping methods**. We report the precision and recall values for each method, varying the PIP threshold from 0 to 1. (see Excel file)

**Supplementary Table 7: Fine-mapped 3Mb loci**. We report the lead variant which resulted in the inclusion of each trait-locus pair. (see Excel file)

**Supplementary Table 8: Number of fine-mapped variant-trait pairs in real trait analysis varying ancestry and sample size**. We report the number of variant-trait pairs for each cohort at PIP > 0.5 and PIP > 0.9. (see Excel file)

**Supplementary Table 9: Number of fine-mapped variant-trait pairs in real trait analysis varying fine-mapping method** (see Excel file)

**Supplementary Table 10: Annotations used in functional enrichment analysis**. We report the name of each annotation. (see Excel file)

**Supplementary Table 11: Functional enrichment of fine-mapped variants varying ancestry and sample size**. We report the functional enrichment averaged across functional annotations and the associated jackknife standard error for each cohort. (see Excel file)

**Supplementary Table 12:**
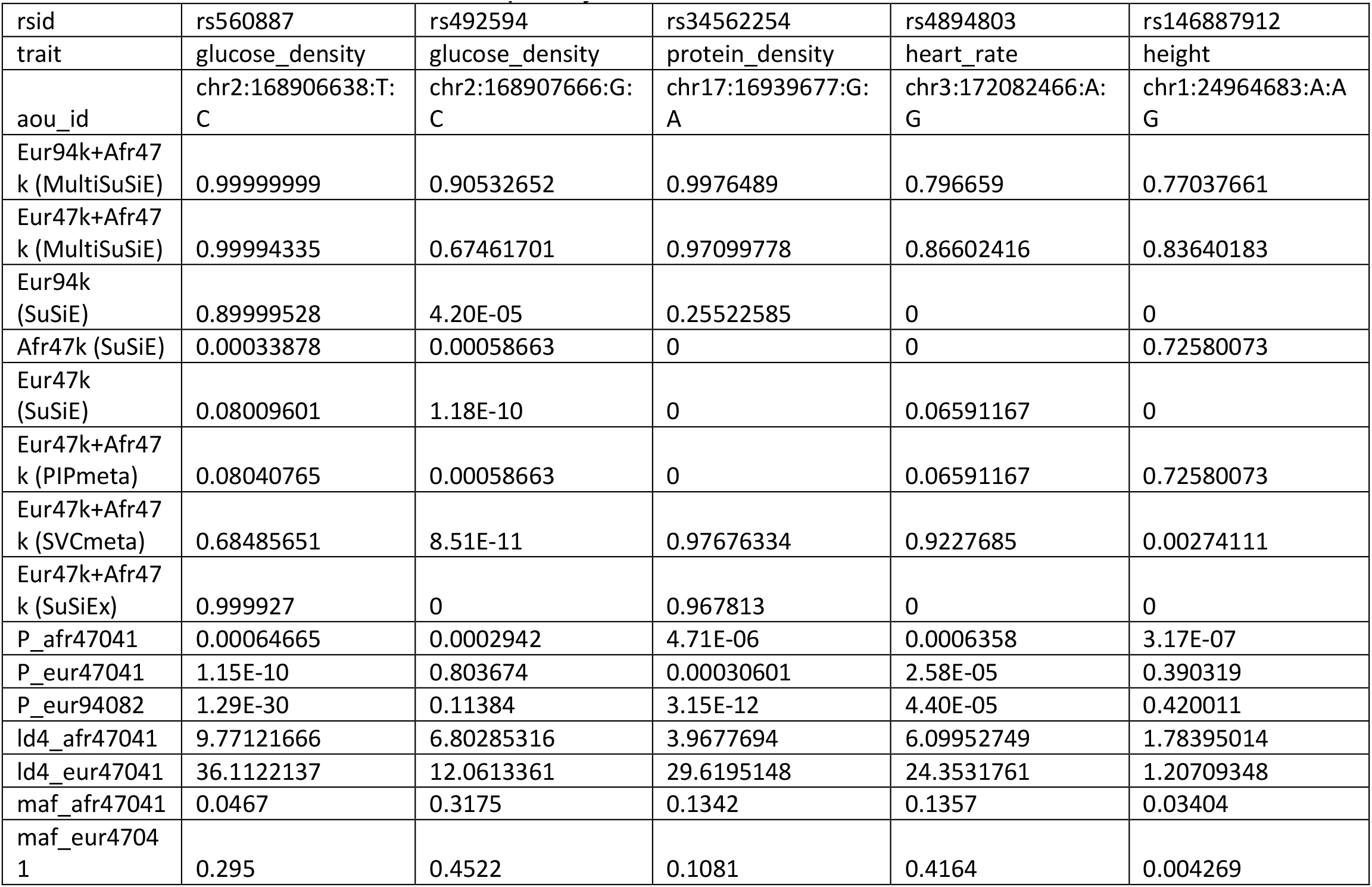
Variant-level statistics of fine-mapped example variants. We report the PIP for five methods and five cohorts, GWAS p-values for 3 cohorts, LD 4^th^ moments for Afr47k and Eur47k, and the minor allele frequency for Afr47k and Eur47k.

**Supplementary Table 13: Eur94k (SuSiE) and Afr47k+Eur47k (MultiSuSiE) PIPs at example loci**. We report the PIPs fine-mapped with MultiSuSiE in Afr47k+Eur47k and SuSiE in Eur94k for all variants included in figure 6. (see Excel file)

**Supplementary Table 14:**
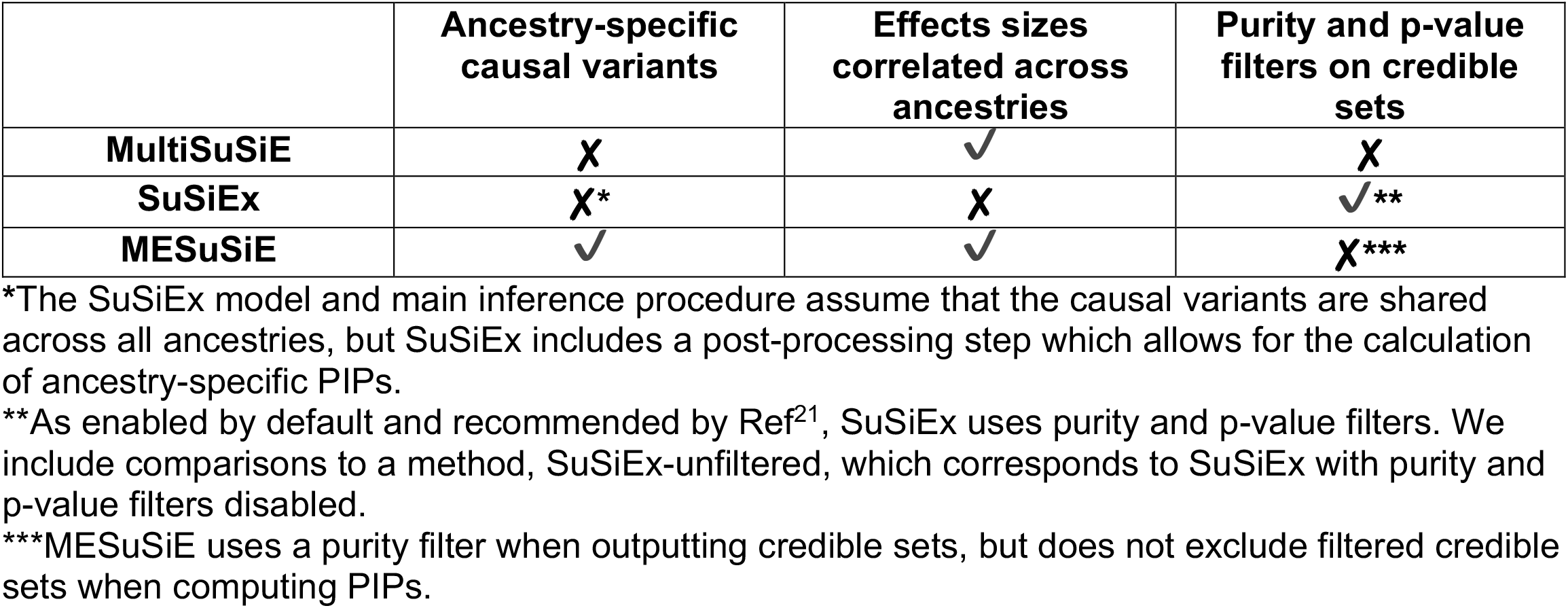
Summary of methodological distinctions between MultiSuSiE, SuSiEx, and MESuSiE. We report the key distinguishing modeling assumptions and algorithmic features (columns) of each method (rows). ✔ indicates the presence of a feature and **✘** indicates the absence of a feature.

**Supplementary Table 15:** Summary of Afr47k fine-mapping results and ordinary least squares at the *HBA* locus. We report variant-level statistics for variants fine-mapped (PIP > 0.5) using SuSiE on Afr47k with *L*=20. Next, a multiple linear regression model was fit for MCV in Afr47k, including all 19 variants with PIP > 0.5, PC1-10, age, age squared, sex, sex*age, sex*age^2^, and genotyping site as explanatory variables. Missing genotypes were mean imputed before running multiple linear regression. “t” and “p” columns refer to t and p-values for the multiple linear regression model.

| rsid | bp | PIP | chrom | maf_afr47041 | maf_eur47041 | t | p |
| --- | --- | --- | --- | --- | --- | --- | --- |
| chr16:19700:A:C | 19700 | 1 | 16 | 0.0561 | 0.026 | 8.156541 | 3.64E-16 |
| chr16:87228:C:CT | 87228 | 0.541782 | 16 | 0.1472 | 0.000908 | 6.380025 | 1.81E-10 |
| chr16:134821:C:T | 134821 | 0.827023 | 16 | 0.03134 | 0.000096 | 6.434434 | 1.27E-10 |
| chr16:144760:C:T | 144760 | 1 | 16 | 0.0298 | 0.000064 | 12.968526 | 2.58E-38 |
| chr16:160978:T:C | 160978 | 0.935577 | 16 | 0.2358 | 0.03937 | 9.088682 | 1.09E-19 |
| chr16:170831:T:A | 170831 | 1 | 16 | 0.01037 | 0.000117 | -7.562462 | 4.12E-14 |
| chr16:174620:T:C | 174620 | 0.999991 | 16 | 0.2718 | 0.4633 | 7.081571 | 1.47E-12 |
| chr16:174791:C:T | 174791 | 0.824321 | 16 | 0.01383 | 0 | -3.714184 | 2.04E-04 |
| chr16:180725:G:C | 180725 | 0.88254 | 16 | 0.0159 | 0.000096 | 18.657321 | 4.58E-77 |
| chr16:187433:C:T | 187433 | 1 | 16 | 0.04694 | 0.000043 | 17.955718 | 1.47E-71 |
| chr16:196434:AAC:A | 196434 | 1 | 16 | 0.02464 | 0.000043 | 9.101419 | 9.69E-20 |
| chr16:197471:G:A | 197471 | 1 | 16 | 0.4577 | 0.07286 | 11.871907 | 2.10E-32 |
| chr16:227192:T:C | 227192 | 0.896196 | 16 | 0.06392 | 0.000202 | -6.731648 | 1.72E-11 |
| chr16:235382:A:C | 235382 | 0.785209 | 16 | 0.01585 | 0.000149 | -5.522709 | 3.38E-08 |
| chr16:242770:G:A | 242770 | 1 | 16 | 0.02839 | 0.000117 | 8.477964 | 2.44E-17 |
| chr16:244190:G:A | 244190 | 0.932163 | 16 | 0.0469 | 0.000542 | -5.749797 | 9.06E-09 |
| chr16:254316:C:T | 254316 | 1 | 16 | 0.06713 | 0.00016 | -13.153898 | 2.31E-39 |
| chr16:264781:G:T | 264781 | 0.79982 | 16 | 0.1006 | 0.000223 | 24.342605 | 4.14E-129 |
| chr16:390165:A:C | 390165 | 0.904074 | 16 | 0.06224 | 0.000191 | 6.384878 | 1.75E-10 |

## Methods

### MultiSuSiE model

Suppose we wish to fine-map a GWAS locus with *P* variants, up to *L* of which have causal effects, using *K* ancestry groups. MultiSuSiE estimates ancestry-specific posterior mean causal effect sizes and an overall posterior inclusion probability (PIP) for each variant by fitting the following model:

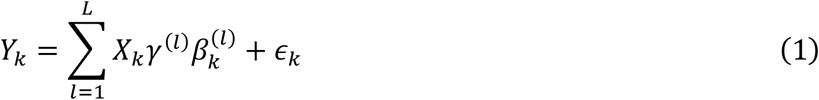

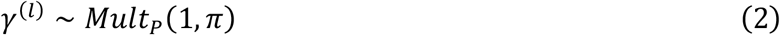

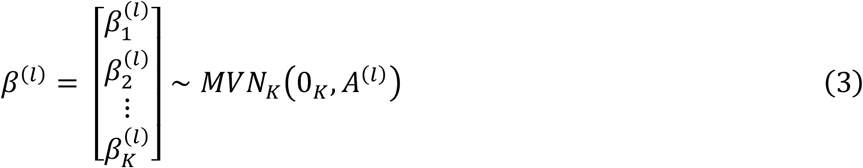

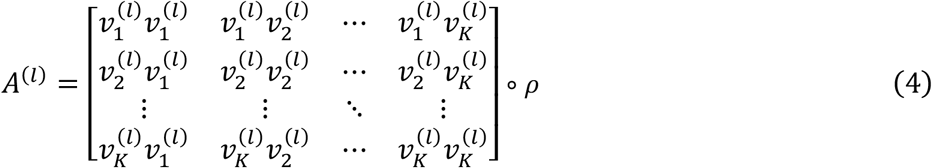

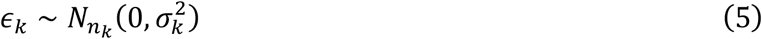

where *Y*_*k*_ is a phenotype vector for ancestry *k, X*_*k*_ is a genotype matrix for ancestry *k, γ*^(*l*)^ is a causal status indicator vector of length *P* for component *l*, 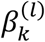 is the scalar effect size in ancestry *k* of the causal variant captured by component *l, ϵ*_*k*_ is a ancestry-specific noise vector, π is a vector of length *P* that gives the prior probability that each variant is causal, 0_*M*_ is a zero matrix of shape *M, A*^(*l*)^ is the effect size covariance matrix for component *l*, 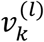 is the effect size prior variance in ancestry *k* for component *l*, ∘ denotes the element-wise product, *ρ* is a *K* × *K* cross-ancestry per-allele effect size correlation matrix, and 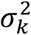 is the residual variance for ancestry *k*. If we set *K* = 1 this is the SuSiE model^13^.

### MultiSuSiE inference

We use iterative Bayesian stepwise selection (IBSS) to estimate posterior distributions for *γ*^(*l*)^ and 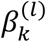. At a high level, our inference procedure proceeds in two steps: (1) for each of the *L* effects in turn, (1a) residualize phenotypes for all ancestries, using all estimated effects except for effect *l*, (1b) update the effect size prior distribution for effect *l*, (1c) estimate the posterior distribution for effect *l* using the residualized phenotypes, and (2) update residual variance prior distributions. Rather than immediately describing the details of our inference procedure, we will first discuss a much simpler model in the <u>Multi-ancestry single variable Bayesian linear regression</u> section. We will then build up additional complexity in the <u>Multi-ancestry single effect Bayesian linear regression</u> section. Next, we give the full IBSS algorithm in <u>Iterative Bayesian stepwise selection for MultiSuSiE</u>. Finally, we discuss additional implementation details in <u>Prior updates</u>, <u>ELBO calculation</u>, <u>Implementation with summary statistics</u>, and <u>Additional implementation details</u>.

#### Multi-ancestry single variable Bayesian linear regression

Multi-ancestry single variable Bayesian linear regressions allow us to estimate ancestry specific effect sizes for a single variable. We’ll derive expressions for the effect size posterior distribution and the Bayes factor for the model.

Suppose we wish to perform a linear regression for a single variant with index *j*. We’ll show how to fit the following model:

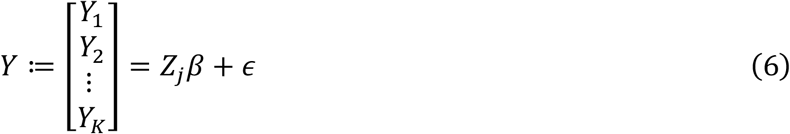

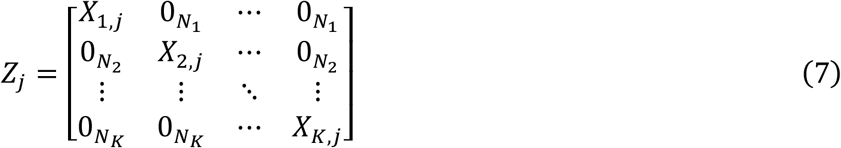

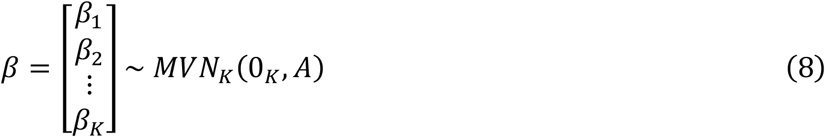

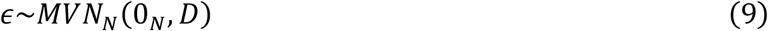

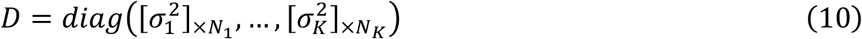

where *X*_*k,j*_ is an *N*_*k*_-vector of genotypes in ancestry *k* for variant *j, N*_*k*_ is the sample size in ancestry *k, β*_*k*_ is the effect size in ancestry *k, A* is the effect size covariance matrix, *ϵ* is a length *N* noise vector with diagonal covariance matrix given by *D*, 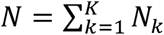, *diag*(*C*) is a diagonal matrix with the vector *C* on its diagonal, [*c*]_×*M*_ is a vector of length *M* in which every element is *c*, and all other terms have been defined in the *MultiSuSiE model* section. Note that we’ve changed the parameterization of the residual noise vector relative to the *MultiSuSiE model* section. We want to be able to calculate the posterior effect size distribution for *β* and the Bayes factor for the model.

To calculate the posterior effect size distribution for *β* we first find the covariance between *β* and *Y* (throughout this section we will treat *Z*_*j*_, *A*, and *D* as fixed, and drop the conditioning from the notation):

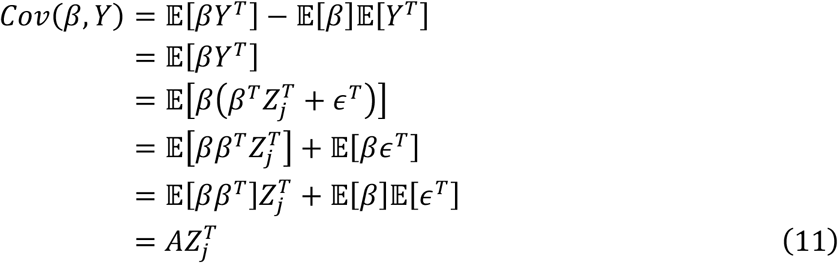

and note the distribution of *Y* is:

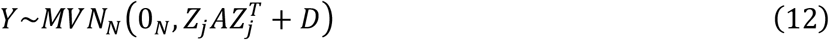

which allows us to write the joint distribution of *β* and *Y* as

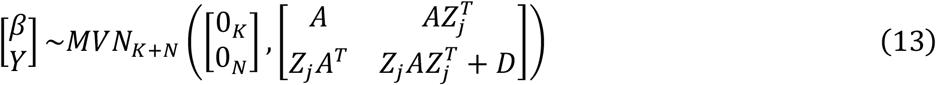

so the the posterior distribution of *β* is:

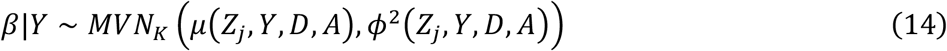

with *μ*(*Z*_*j*_, *Y, A, D*) and *ϕ*^2^(*Z*_*j*_, *Y, A, D*) defined as:

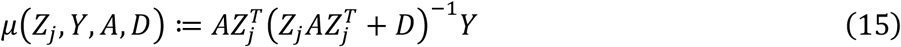

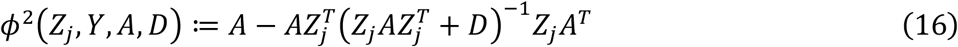

However, computing *μ*(*Z*_*j*_, *Y, A, D*) and *ϕ*^2^(*Z*_*j*_, *Y, A, D*) as written requires us to invert 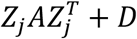, which is of dimension *N* × *N*, and will be expensive for large *N*. We can use the Woodbury matrix identity to get:

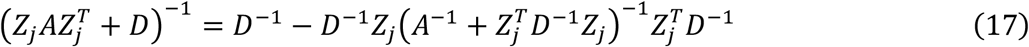

which only requires us to invert matrices of dimension *K* × *K*. Plugging this into *μ*(*Z*_*j*_, *Y, A, D*) and *ϕ*^2^(*Z*_*j*_, *Y, A, D*), we get

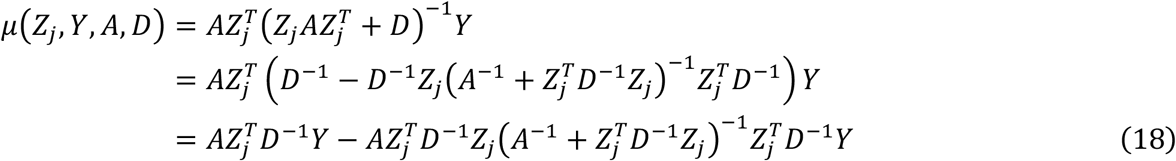

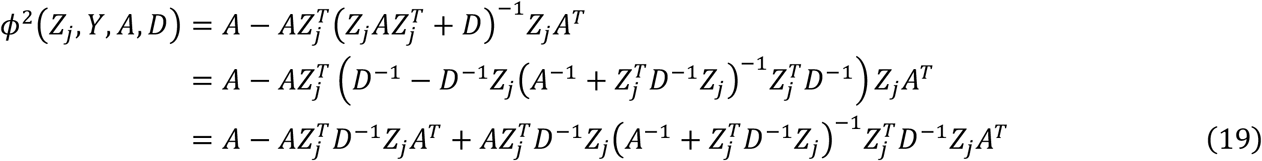

Next, let’s move on to calculating the likelihood and Bayes factor for the model. Using equation 12, we know the probability density function is (denoting the determinant of *C* with |*C*|):

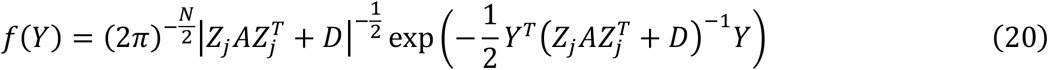

Here, computing 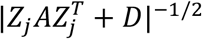 will be expensive. Instead, we’ll use to matrix determinant lemma to get:

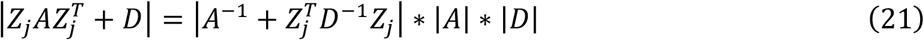

Combining this with the Woodbury matrix identity as used above, we can write the log probability density function as:

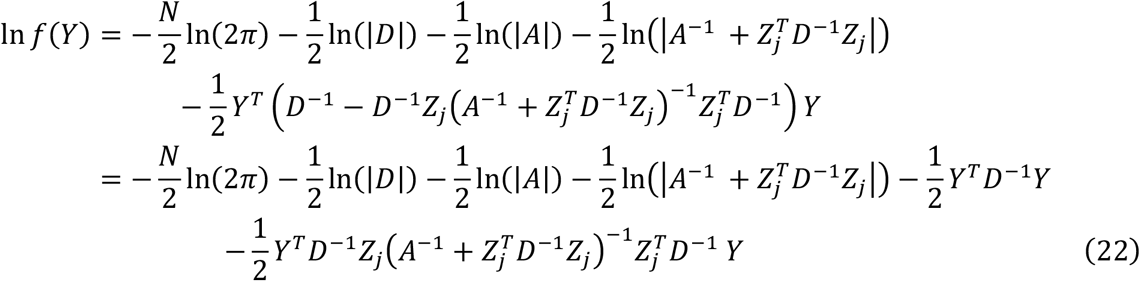

To calculate the Bayes factor, we’ll need the likelihood under the null model, In *f*_0_(*Y*), with *β* set to 0:

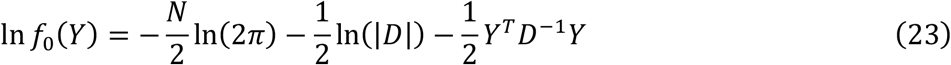

we’ll define a function which returns the Bayes factor for this model in terms of *Z*_*j*_, *Y, A, D* to make the next section more clear.

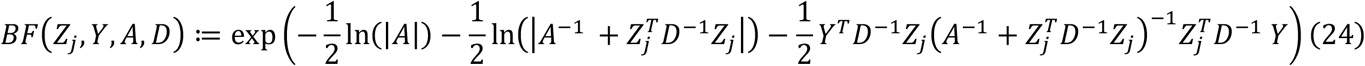

### Multi-ancestry single effect Bayesian linear regression

Multi-ancestry single effect Bayesian linear regressions allow us to estimate ancestry specific effect sizes and the probability that each variant is causal under the assumption that only one variant is truly causal (hence “single effect”). We can fit this model by calculating multi-ancestry single variable Bayesian linear regressions for each variant.

Consider the following model, which now includes all *P* variants:

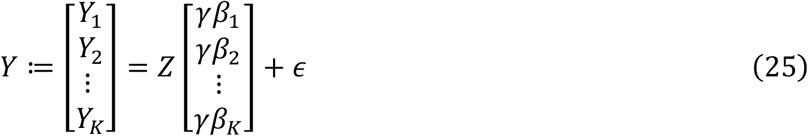

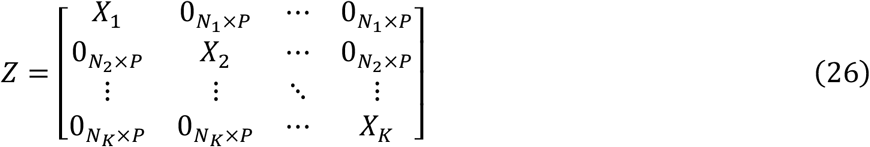

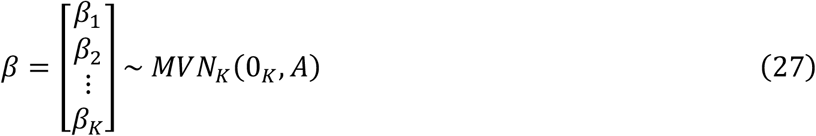

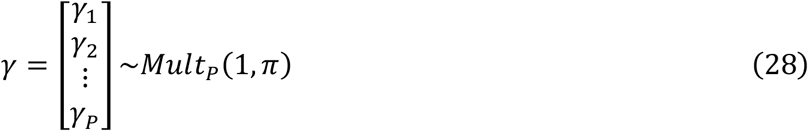

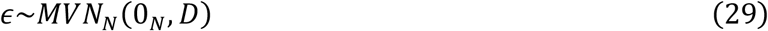

note that *β*_*k*_ is a scalar, so *γβ*_*k*_ is a is a *P* × 1 matrix.

We’re specifically interested in estimating posterior distributions for *γ* and β|*γ*_*j*_ = 1. We can calculate these quantities by fitting *P* multi-ancestry single variable Bayesian linear regressions:

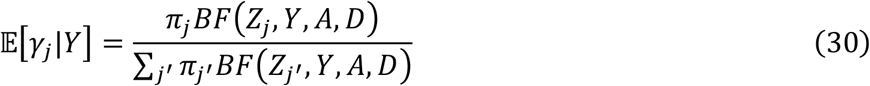

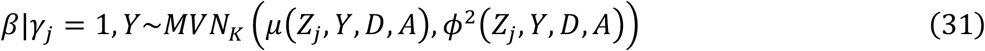

To make things easier later, we’ll define a multi-ancestry single effect Bayesian linear regression function which returns our posterior estimates (separated by ancestry) as follows:

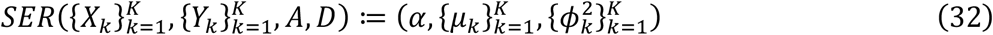

where α = [𝔼[*γ*_1_|*Y*], …, 𝔼[*γ*_*P*_|*Y*]{ is the *P*-vector of posterior inclusion probabilities, *μ*_*k*_ = [*μ*(*Z*_1_, *Y, A, D*)_*k*_, …, *μ*(*Z*_*P*_, *Y, A, D*)_*k*_] is a *P*-vector of effect size posterior means for ancestry *k* (conditional on each variant being the causal variant), 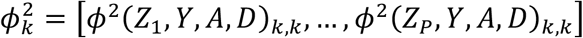 is a *P*-vector of effect size posterior variances for ancestry *k* (conditional on each variant being the causal variant), and 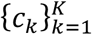 denotes an ordered list. Here we’re denoting *ϕ*^2^(*Z*_1_, *Y, A, D*)_*k,k*_ as the *k*th diagonal element of *ϕ*^2^(*Z*_1_, *Y, A, D*) and *μ*(*Z*_*P*_, *Y, A, D*)_*k*_ as the *k*th element of *μ*(*Z*_*P*_, *Y, A, D*).

It will be useful later to have defined a function which returns the likelihood under this model.

Using the law of total probability, we can write:

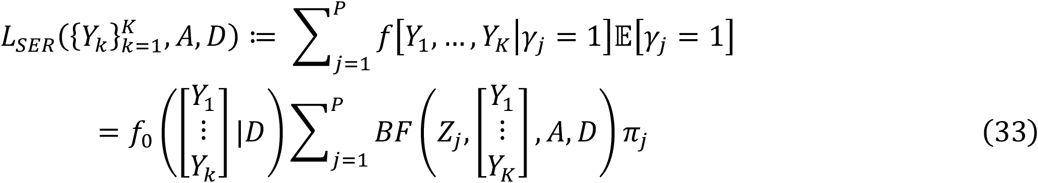

### Iterative Bayesian stepwise selection for MultiSuSiE

MultiSuSiE accounts for multiple causal variants by summing over multiple multi-ancestry single effect Bayesian linear regressions, each which assume and capture a single causal variant. To fit this model, we use iterative Bayesian stepwise selection, which entails sequentially estimating each single effect model while residualizing the phenotype with respect to all other single effects.

To fit the model with multiple causal variants given in *MultiSuSiE overview (*equations 1-5), we use the algorithm presented in Box 1:

**Box 1:** Pseudocode for MultiSuSiE using iterative Bayesian stepwise selection.

|  |  |
| --- | --- |
| while ($F_i - F_{i-1} > .001$): | # Check convergence |
| 1. for $l$ in $1, 2, \dots, L$: | |
| 1a. $\{r_k^{(l)}\}_{k=1}^K = \{Y_k - \sum_{l' \neq l} X_k (\alpha^{(l')} \circ \mu_k^{(l')})\}_{k=1}^K$ | # Residualize association statistics |
| 1b. $A^{(l)} = EM\left(\alpha^{(l)}, \{\mu_k^{(l)}\}_{k=1}^K, \{\phi_k^{(l)^2}\}_{k=1}^K, \rho\right)$ | # Update effect size priors |
| 1c. $\left(\alpha^{(l)}, \{\mu_k^{(l)}\}_{k=1}^K, \{\phi_k^{(l)^2}\}_{k=1}^K\right) = SER\left(\{X_k\}_{k=1}^K, \{r_k^{(l)}\}_{k=1}^K, A^{(l)}, D\right)$ | # Estimate effect sizes |
| 2. $\{\sigma_k^2\}_{k=1}^K = \left\{ERSS\left(Y_k, \alpha^{(l)}, \{\mu_k^{(l)}\}_{l=1}^L, \{\phi_k^{(l)^2}\}_{l=1}^L\right) / N_k\right\}_{k=1}^K$ | # Update noise priors |
| 3. $D = diag([\sigma_1^2]_{\times N_1}, [\sigma_2^2]_{\times N_2}, \dots, [\sigma_K^2]_{\times N_K})$ | # Update noise priors |
| 4. $F_{i-1} = F_i$ | # Update objective function |
| 5. $F_i = ELBO\left(\{Y_k\}_{k=1}^K, \left\{\{\mu_k^{(l)}\}_{k=1}^K\right\}_{l=1}^L, \left\{\{\phi_k^{(l)^2}\}_{k=1}^K\right\}_{l=1}^L, \{\alpha^{(l)}\}_{l=1}^L, \{A^{(l)}\}_{l=1}^L, D\right)$ | # Update objective function |

where *F*_*i*_ is the value of the objective function at iteration *i*, 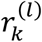 are phenotypes for ancestry *k* with effects from all components other than component *l* residualized out, α^(*l*)^ is a vector of causal status probabilities for component *l*, 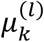 is a vector of posterior effect size means for ancestry *k* in component *l* (conditional on each variant being causal), *EM* is a function which returns an updated effect size variance prior, 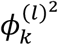 is a vector of posterior effect size variances for ancestry *k* in component *l* (conditional on each variant being causal), *ERSS* is a function which returns the expected residual sum of squares in ancestry *k*, and *ELBO* is a function which returns the evidence lower bound. To estimate posterior inclusion probabilities, we adopt the approach of Ref^13^ and combine estimates of α^(*l*)^ across single-effect models:

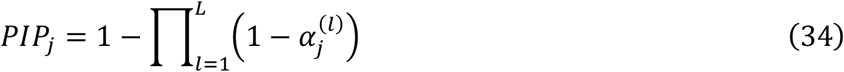

where 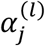 is the *j*th element of α^(*l*)^.

In <u>Prior updates</u> we’ll discuss *EM* and *ERSS* from Box 1 and in <u>ELBO calculation</u> we’ll discuss *ELBO* from Box 1.

### Prior updates

The effect size prior distribution for each component-ancestry pair and the residual noise prior for each ancestry are estimated from the data using empirical Bayes procedures. To update the effect size priors, we set the effect size variance parameter to match our current effect size estimates. To update the residual noise prior, we estimate how much phenotypic variance remains after accounting for the current estimated effects.

Each single effect model has a distinct effect size prior covariance matrix given by equation 4 and reproduced below:

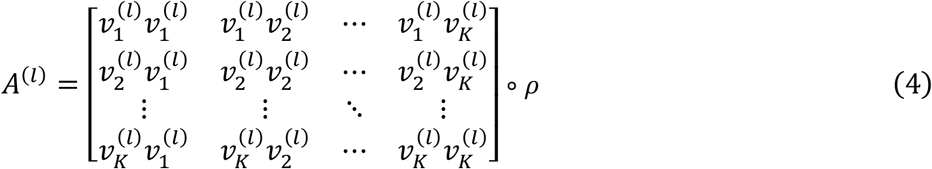

in which 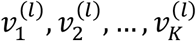 are updated using an expectation-maximization algorithm, and *ρ* is treated as fixed. The expectation step corresponds to step 1c from Box 1 and the maximization step corresponds to step 1b from Box 1.

Specifically, define 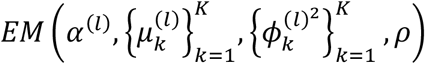 from step 1b as the function that returns *A*^(*l*)^ with 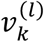 set to 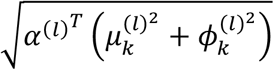. This quantity does not maximize the joint probability density function for *β*^(*l*)^ ∼ *MVN*_*K*_(0_*K*_, *A*^(*l*)^), but does maximize the probability density function for 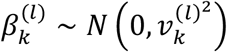, treating effects from each ancestry as independent. Surprisingly, with two ancestries, we observed poor performance when we use an MLE estimator based on β^(*l*)^ ∼ *MVN*_*K*_ (0_*K*_, *A*^(*l*)^) where we set 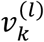 to 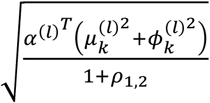 (*ρ*_1,2_ is the cross-ancestry correlation of per-allele effect sizes between ancestries 1 and 2).

After a user-specified number of iterations, at each iteration, EM additionally compares the single effect regression likelihood using our current estimates of 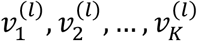 to the single effect regression likelihood where we set each of 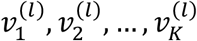 to 0, one at a time, and the single effect regression likelihood where we set all of 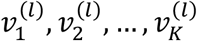 to 0, simultaneously. EM then returns the values of 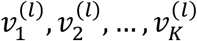 which maximized the likelihood across these *K* + 2 options.

To derive the MLE for a single ancestry, neglecting the cross-ancestry correlation of effect sizes:

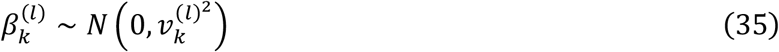

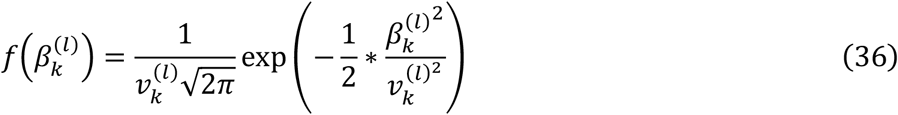

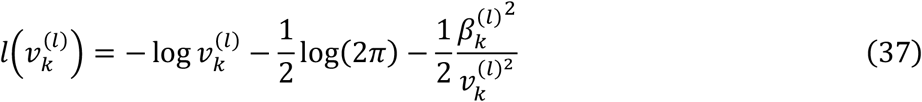

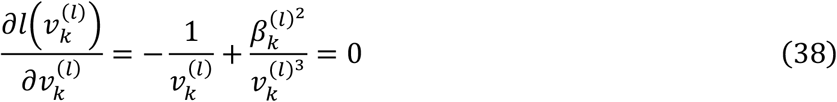

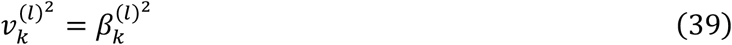

but we don’t know the true value of 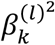 so we plug in 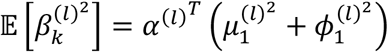 from step 1c.

To derive the MLE for two ancestries, allowing for cross-ancestry effect size correlations:

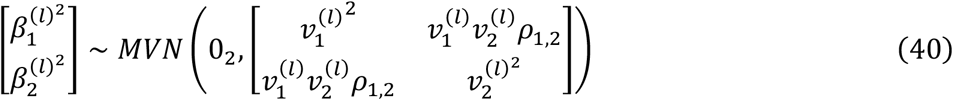

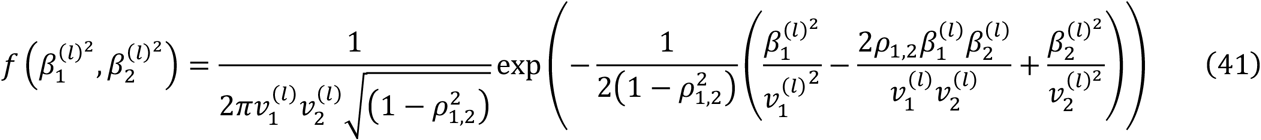

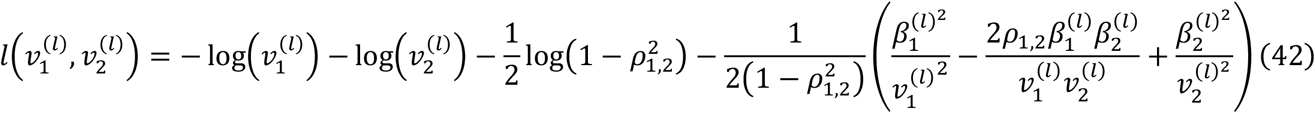

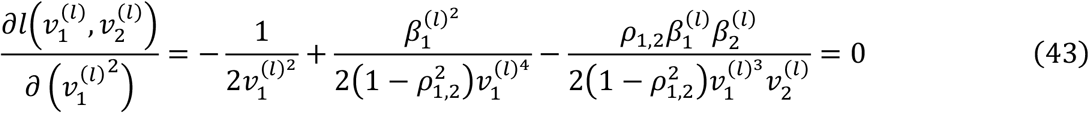

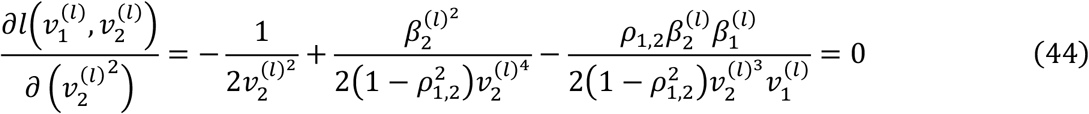

which gives us 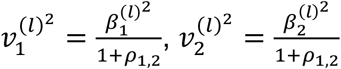 if 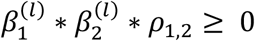 and 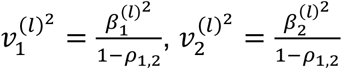 otherwise. We initially used 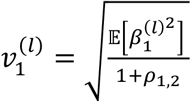 and 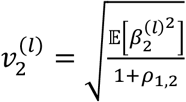to update *A*^(*l*)^ but observed poor performance and chose to treat effects as independent across ancestries when updating *A*^(*l*)^.

To update 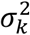, we use the expected residual sum of squares for ancestry *k* (see Ref^13^ for a derivation):

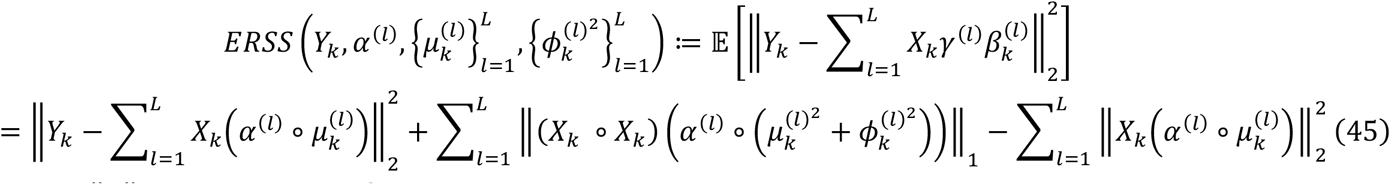

where ‖*x*‖_*c*_ is the *c*-norm of *x*.

### ELBO calculation

The evidence lower bound (ELBO) is a lower bound on the model log likelihood. We use the change in the ELBO to assess convergence.

The ELBO can be written as:

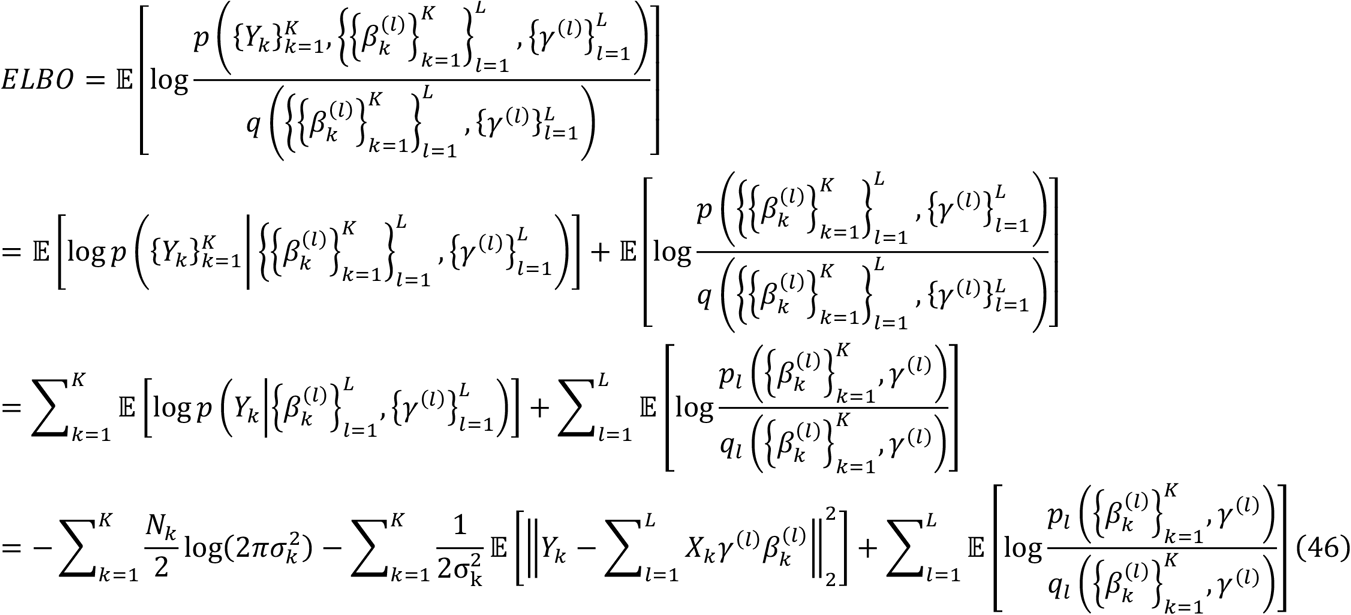

where *p* is probability density function under the generative model or prior and *q* is the probability density function under our estimate of the posterior distribution. *p*_*l*_ and *q*_*l*_ are the corresponding functions for the *l*th single effect regression. *p, q, p*, and *q* are conditioned on 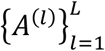 and *D*. 𝔼 is the expectation taken with respect to *q*. The third equality holds under a variational approximation that the posterior distribution factorizes as 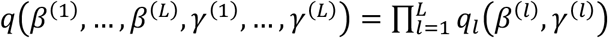.

To compute the second term we can use the ERSS function defined above. To compute the third term, we’ll use an argument from Ref^13^ in an abbreviated form. We can write the ELBO for a single effect regression model as:

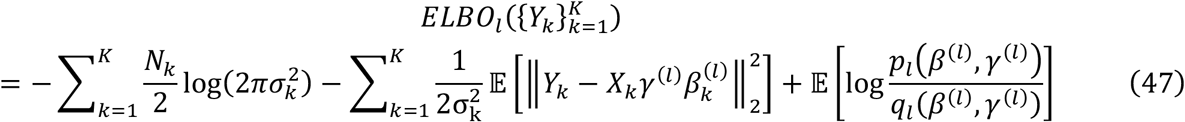

Because we’re able to calculate the exact posterior distribution for *q*_*l*_ (fixing the other *L* – 1 effect estimates), the KL divergence between *q*_*l*_ and the true posterior is equal to zero, and as a result, 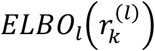 is equal to the log-likelihood of the residuals under the single effect regression model. We can use this to simplify calculation of the third term in the overall ELBO:

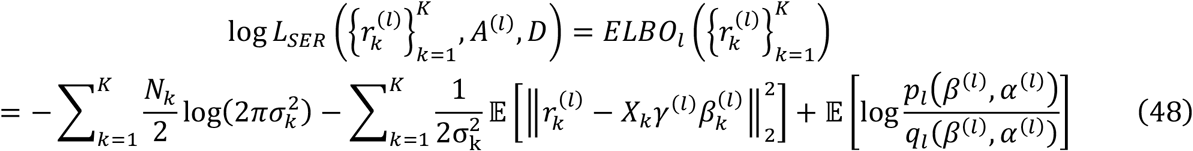

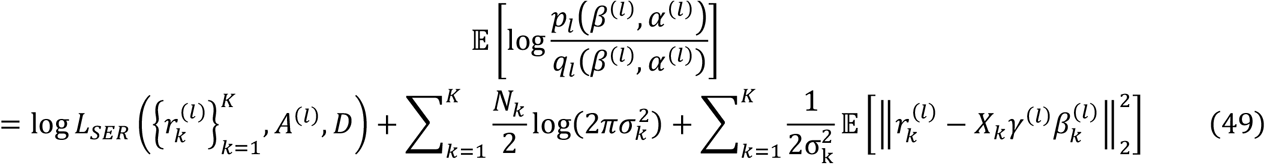

We’ve discussed how to calculate the first term in <u>Multi-ancestry single effect Bayesian linear regression</u> (equation 33) and the third term in <u>Prior updates</u> (equation 45). This implies that we can define a function which computes the overall ELBO,

*ELBO* 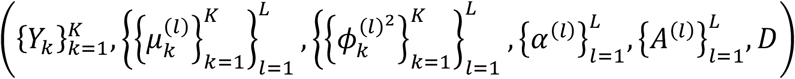, which is used in Box 1.

### Implementation with summary statistics

So far we’ve discussed fitting MultiSuSiE using individual-level phenotype and genotype data. However, fitting MultiSuSiE using summary statistics is relatively straightforward and can reduce runtime and memory requirements. The computational complexity of MultiSuSiE with individual level data is *O*(*MLNP*), where *M* is the number of iterations until convergence (using a refactored algorithm equivalent to Box 1 and fixing the number of ancestries). Below, we describe an approach which requires only summary statistics and has computational complexity of *O*(*MLP*^2^), neglecting the cost of computing LD matrices; computing LD matrices has complexity of *O*(*NP*^2^), so the overall complexity is *O*(*MLP*^2^ + *NP*^2^). Despite this, the summary statistics-based procedure has much lower maximum memory requirements and makes large-scale fine-mapping of loci with tens of thousands of variants and hundreds of thousands of samples feasible.

When fitting multi-ancestry single variable Bayesian linear regressions, individual level data are only used in the form of 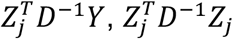, and *Y*^*T*^*D*^−1^*Y*, all of which can be written as functions of 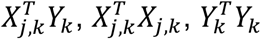, and 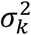:

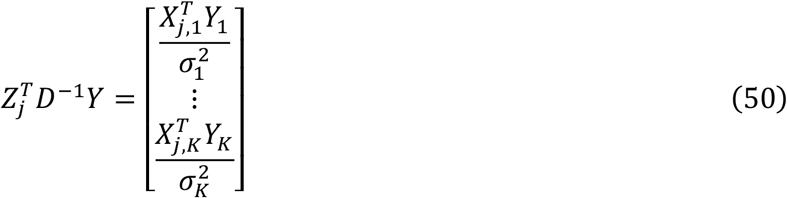

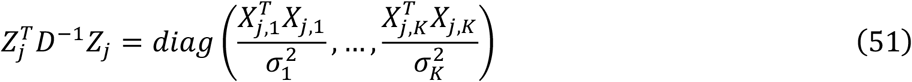

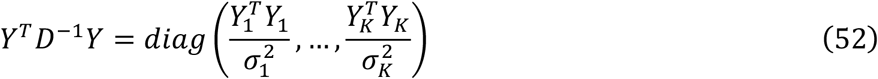

This allows us to define functions analogous to those from <u>Multi-ancestry single variable Bayesian linear regression</u> and <u>Multi-ancestry single effect Bayesian linear regression</u> which perform all relevant single effect calculations using 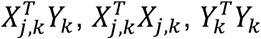 and 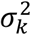, instead of individual level data. We’ll denote these functions as 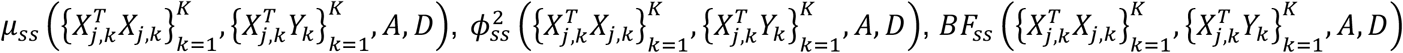, and 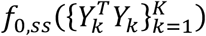. As described in Ref^18^, 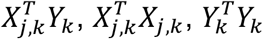, and 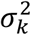 can be recovered from standard GWAS summary statistics (specifically, linear model marginal variant effect sizes, linear model marginal variant effect size standard errors, in-sample LD, trait sample variance, and GWAS sample size).

Therefore, we can fit multi-ancestry single variant linear models with standard summary statistics. Furthermore, multi-ancestry single effect Bayesian linear regressions only require the results of all *P* multi-ancestry single variable Bayesian linear regressions, so we can fit multi-ancestry single effect Bayesian linear regressions using only summary statistics. This allows us to define 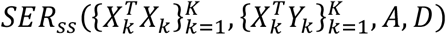 which returns effect estimates, 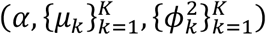, analogous to 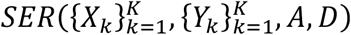, and 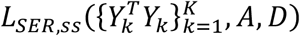, which returns the single effect regression likelihood, analogous to 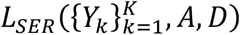.

To implement MultiSuSiE with summary statistics, we follow the approach of Ref^18^ by tracking and residualizing variant effects on 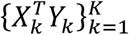, rather than individual level phenotypes, 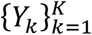. Box 2 describes the adapted iterative Bayesian stepwise selection algorithm for multiple ancestries with summary statistics.

**Box 2:** Pseudocode for MultiSuSiE using iterative Bayesian stepwise selection with association statistics.

|  |  |
| --- | --- |
| while ($F_i - F_{i-1} > .001$): | # Check convergence |
| 1. for $l$ in $1, 2, \dots, L$: | |
| 1a. $\{\psi_k^{(l)}\}_{k=1}^K = \{X_k^T Y_k - \sum_{l' \neq l} X_k^T X_k (\alpha^{(l')} \circ \mu_k^{(l')})\}_{k=1}^K$ | # Residualize phenotypes |
| 1b. $A^{(l)} = EM(\alpha^{(l)}, \{\mu_k^{(l)}\}_{k=1}^K, \{\phi_k^{(l)^2}\}_{k=1}^K, \rho)$ | # Update effect size priors |
| 1c. $(\alpha^{(l)}, \{\mu_k^{(l)}\}_{k=1}^K, \{\phi_k^{(l)^2}\}_{k=1}^K) = SER_{ss}(\{X_k^T X_k\}_{k=1}^K, \{\psi_k^{(l)}\}_{k=1}^K, A^{(l)}, D)$ | # Estimate effect sizes |
| 2. $\{\sigma_k^2\}_{k=1}^K = \{ERSS_{ss}(Y_k^T Y_k, X_k^T Y_k, X_k^T X_k, \alpha^{(l)}, \{\mu_k^{(l)}\}_{l=1}^L, \{\phi_k^{(l)}\}_{l=1}^L) / N_k\}_{k=1}^K$ | # Update noise priors |
| 3. $D = diag([\sigma_1^2]_{\times N_1}, \dots, [\sigma_K^2]_{\times N_K})$ | # Update noise priors |
| 4. $F_{i-1} = F_i$ | # Update objective function |
| 5. $F_i = ELBO_{ss}(\{Y_k^T Y_k\}_{k=1}^K, \{\{\mu_k^{(l)}\}_{k=1}^K\}_{l=1}^L, \{\{\phi_k^{(l)^2}\}_{k=1}^K\}_{l=1}^L, \{\{\psi_k^{(l)}\}_{k=1}^K\}_{l=1}^L, \{A^{(l)}\}_{l=1}^L, D)$ | # Update objective function |

where 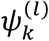 are residualized association statistics for ancestry *k*, with effects from all components other than component *l* residualized out, *ERSS*_*ss*_ is a function which returns the expected residual sum of squares in ancestry *k*, and *ELBO*_*ss*_ is a function which returns the ELBO. To complete our description of the algorithm, we should discuss *ERSSss* and *ELBOss*. As shown in Ref^18^, the expected residual sum of squares can also be computed in terms of association statistics:

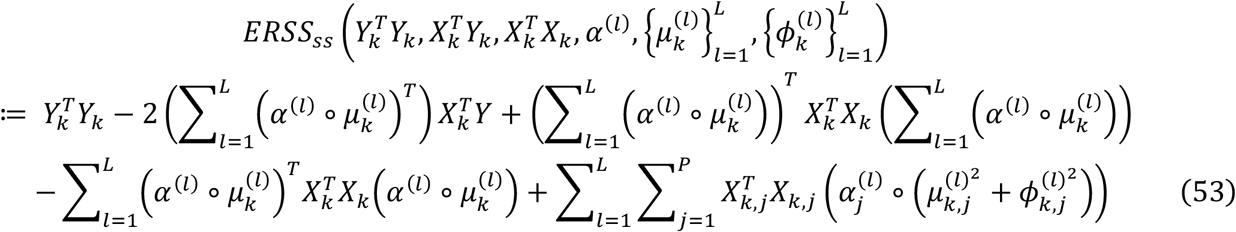

where 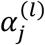 is the *j*th element of 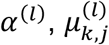 is the *j*th element of 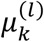, and 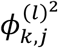 is the *j*th element of 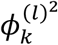.

Calculating the ELBO using only summary statistics requires functions that return the expected residual squares and single effect model likelihoods, both of which can be implemented using summary statistics as discussed above.

### Additional implementation details

GWAS summary statistics may be poorly calibrated for variants with low allele counts^41^. MultiSuSiE zeros out columns and rows of 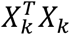 and rows of 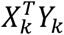 for variants with an expected minor allele count in ancestry *k* (estimated from genotypic variance, assuming Hardy Weinberg equilibrium) of 20 or less.

### Ancestry definition

We used discrete continental ancestry classifications as provided by All of Us (“ancestry_pred_other” column from the srWGS auxiliary file). In short, All of Us genetic data was projected onto the first 16 principal components of a dataset with genetic ancestry annotations containing 1000 Genomes^86^ and The Human Genome Diversity Project^87^ data. A random forest was then trained on the annotated dataset, using principal components as features, and used to predict genetic ancestry for each All of Us sample. See Ref^88^ for additional details. Admixture proportions were calculated by interpolating between minimum and maximum PC1 values of all African or European ancestry individuals.

### Simulations - generative model

We simulated 10 3Mb quantitative trait GWAS loci on chromosome 11, similar to previous work^12^, using empirical African-ancestry and European-ancestry LD (17,262-23,634 variants per locus; Supplementary Table 1) from the All of Us v7 short read whole-genome sequencing Allele Count/Allele Frequency threshold Plink dataset. For data processing details including sample and variant quality control and filtering, see Ref^88^. We restricted to variants with minor allele frequency (MAF) > 0.01 in either Afr47k or Eur47k and missingness < 0.05 in both Afr47k and Eur47k. Due to the high cost of running simulations on the All of Us Researcher Workbench, we opted to download LD matrices and run simulations on the Harvard Medical School O2 high performance compute cluster. We defined cohorts of unrelated individuals from specific ancestries: Eur94k (*n*=94,082), Afr47k (*n*=47,041), Eur47k (*n*=47,041), Afr23k (*n*=23,520), and Eur23k (*n*=23,520).

Smaller cohorts were nested within larger cohorts of the same ancestry. For each cohort, we computed in-sample LD on the All of Us Researcher Workbench with LDstore2^89^ (setting compression high). To avoid constraints on downloading large files from the All of Us Researcher Workbench, we then converted LD matrices to numpy NPY format, encoding values as float16 to reduce file size, split matrices into 100MB chunks, downloaded chunks one at a time, and reconstructed the original LD matrix. We then simulated summary statistics directly from LD matrices using the RSS likelihood^30^, 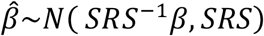, where β is a vector of per-allele causal effect sizes measured without error, 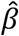 is a vector of simple linear regression effect sizes, *S* is a diagonal matrix with simple linear regression standard errors on the diagonal, and *R* is an in-sample LD matrix. We used a regularized LD matrix, *R* ∗ (1 − λ) + λ*I*, where λ is the smallest value such that 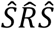 is invertible for both Afr47k and Eur47k (not considering monomorphic variants or variants in complete LD, λ ≤ 0.0^2^6 for all loci) and assumed that all effects were small so that the standard error for a variant with sample size *n* and genotype vector *X*_*j*_ could be approximated by 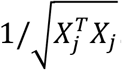.

At each 3Mb locus, we randomly selected 5 causal variants in the central 1Mb. Causal variants had a *p*_*shared*_ probability of having causal effects in both ancestries, a 1 − *p*_*shared*_/^2^ probability of having a causal effect only in African-ancestry individuals, and a 1 − *p*_*shared*_/^2^ probability of having a causal effect only in European-ancestry individuals. Next, we sampled per-allele causal effect sizes using a multivariate normal distribution with mean 0 and correlation *ρ* for variants with causal effects in both ancestries, and a normal distribution with mean 0 for variants with causal effects in only one ancestry. We then scaled all effect sizes at each locus such that the locus-wide heritability of the more heritable ancestry was equal to 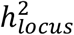 (at each locus, the ratio of heritabilities across ancestries varies due to differences in causal variant MAF). Effect size sampling was restarted if the sum of causal genotype variance was less than 0.095 in either ancestry (0.095 is the variance of a single variant with minor allele frequency of 0.05). In main simulations, we set 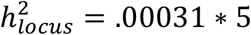 (such that on average, at *N*=94,401, the expected chi-squared statistic for a causal variant with no LD corresponds to a p-value of 5*10^−8^), *p*_*shared*_ = 0.764, and *ρ* = 0.93 (which results in an overall mean per-locus cross-ancestry correlation close to 0.75). In low heritability simulations, we decreased 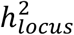 3-fold and in high heritability simulations we increased 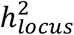 3-fold. In simulations where cross-ancestry differences in causal effects were either entirely due to differences in causal variant identity, or entirely due to differences in per-allele effect sizes at shared causal variants, we used a numerical optimization algorithm to identify a value of *p*_*shared*_ or *ρ* (and set *ρ* or *p*_*shared*_ equal to 1) such that the overall mean per-locus cross-ancestry correlation of causal effect sizes across all simulations was equal to 0.75. In simulations restricted to a subset of variants with similar MAF distributions in Afr47k and Eur47k, we divided variants into 51 MAF bins (2 equal width bins from 0 to 0.01 and 49 equal width bins from 0.01 to 0.50) and, for each locus, randomly selected variants from each ancestry and bin to maximize the total number of variants included, while constraining the number of variants in each bin to be equal across ancestries.

### Simulations – fine-mapping

All methods were run using summary statistics and in-sample LD. We simulated each 3Mb locus 1000 times. All simulations were fine-mapped using each cohort-method pair, except for Afr47k+Eur47k with MESuSiE (fine-mapped each 3Mb locus 500 times), PAINTOR (fine-mapped 1 replicate-window pair), and MGfm (fine-mapped 1 replicate-window pair). For Figure 1c, Supplementary Figures 6-8, and Supplementary Tables 3-5, we fine-mapped an additional 3,000 simulations per locus (4,000 in total).

We applied MultiSuSiE to Afr47k+Eur47k and Afr23k+Eur23k using the susie_multi_rss function with rho = numpy.array([[1, .75], [.75, 1]]) and low_memory_mode = True.

We implemented single-ancestry SuSiE fine-mapping using the MultiSuSiE Python package to decrease computational costs. We compared our implementation to the standard susieR implementation by fine-mapping Eur94k across 100 simulations for each 3Mb window (using estimate_prior_method=“EM”, standardize=FALSE, min_abs_corr=0, float_type=np.float64) and found that the estimated PIPs were nearly identical (Pearson correlation= 0.999999999, maximum difference in PIP= 0.000785).

SuSiEx was run with keep_amibg=True, and multi-step=True. Note that multi-step=False by default, but was enabled in Ref^21^. SuSiEx was run using the same set of variants as MultiSuSiE, filtering variants with minor allele count less than 20 for each ancestry. To run SuSiEx-unfiltered, we additionally set min-purity=0 and pval_thresh=1. We combine SuSiEx PIPs across single effect models using 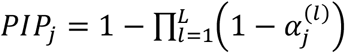, where 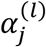 is the PIP of variant *j* in single effect model *l*.

MESuSiE was run using the meSuSie_core function from the MESuSiE R package with default parameters. MESuSiE was run using the same set of variants as MultiSuSiE, censoring, for each ancestry, variants with minor allele count less than 20. MESuSiE-intersection was run by filtering the input GWAS summary statistics and LD matrices to variants with MAF > 0.01 in both Afr47k and Eur47k.

MCVmeta was implemented by meta-analyzing GWAS summary statistics across ancestries using a custom implementation of inverse variance based meta-analysis^42^. For each variant, ancestries with a minor allele count less than 20 were assigned a weight of 0. LD matrices were meta-analyzed across ancestries using the method given in Ref^33^. Due to missing genotypes, the meta-analyzed LD matrix is not exactly equal the correlation matrix calculated using a concatenated genotype matrix 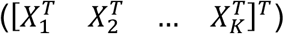. We then ran single-ancestry SuSiE on the meta-analyzed GWAS summary statistics and LD matrix with the parameter settings given above.

SCVmeta was implemented by running single-ancestry SuSiE with L=1 on meta-analyzed GWAS summary statistics, giving the identity matrix as LD input.

PIPmeta was implemented by applying single-ancestry SuSiE to each ancestry separately and meta-analyzing the results as 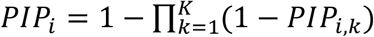, where *PIP*_*i,k*_ is the PIP of variant *i* estimated using SuSiE in ancestry *k*. This approach is justified as follows:

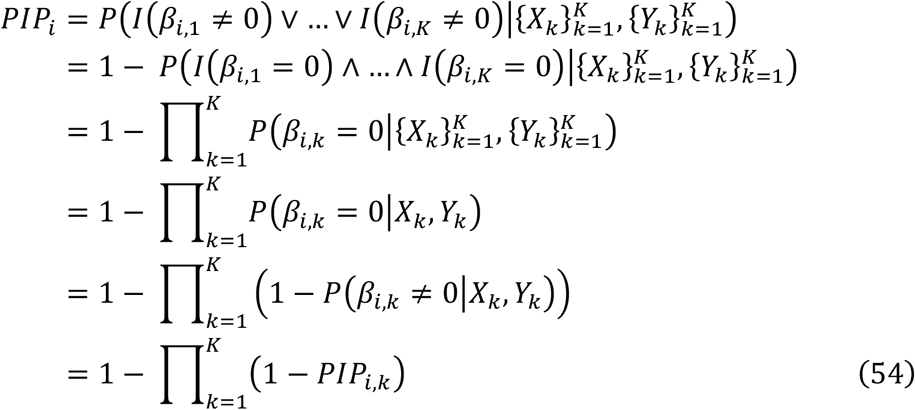

Where *β*_*i,k*_ is the causal effect size of variant *i* in ancestry *k*. The third and fourth equalities hold under the assumption that causality status is independent across ancestries. Although this assumption is false, our post-hoc analysis method outperforms or performs comparably to three alternative post-hoc analysis methods (Supplementary Figure 26).

MGfm was run using the MGFMwithJAM function from the MGflashfm R package. To reduce computational runtime, we set maxcv_stop=5.

PAINTOR was run using the PAINTOR_V3.0 (v3.1) package with two distinct parameter settings. First, we ran PAINTOR with -enumerate 2 and the method failed to complete in 48 hours. Next, we ran PAINTOR with -mcmc and reported the runtime in Table 2.

To calculate the running time of each method, we ran each method on 100 simulated replicates of each 3Mb locus. Each method, except for MGfm and PAINTOR, was benchmarked using hundreds of single-core slurm jobs, only including successfully completed fine-mapping runs (occasionally, MESuSiE non-reproducibly failed with error code Signals.SIGILL: 4). To calculate maximum memory usage for each method, we used the MaxRSS value reported by the sacct command to find the maximum RAM utilized by each slurm job used for the fine-mapping method.

The statistical significance of power differences between pairs of methods was assessed using a continuity corrected McNemar test. Error bars correspond to 95% Agresti-Coull binomial confidence intervals.

### All of Us Summary Statistics

We fine-mapped 14 traits from the All of Us short read whole-genome sequencing v7 Allele Count/Allele Frequency threshold Plink dataset. For genetic data processing details including sample and variant quality control (QC) and filtering, see Ref^88^. We restricted to variants with minor allele frequency (MAF) > 0.01 in either Afr47k or Eur47k and missingness < 0.05 in both Afr47k and Eur47k.

To select traits, we started with 5 quantitative traits derived from manually collected physical measurements and 29 quantitative traits derived from electronic health records (EHR) with reported sample size greater than 140,000 and heritability > 0.04 (as reported by Pan-UKBioBank^90^). For each manually collected physical measurement trait, within each individual, we averaged repeated measurements. For each EHR trait, within each individual, we selected the measurement with the most common combination of the “standard_concept_name”, “unit_concept_name”, “operator_concept_name”, “measurement_type_concept_name”, “visit_occurrence_concept_name” fields for that trait. We then dropped individuals based on the “operator_concept_name”,”standard_concept_name”, “unit_concept_name”, “measurement_type_concept_name”, “unit_source_value” fields to increase measurement consistency. For all traits, we iteratively dropped outliers (defined as individuals more than six standard deviations from the trait mean) until no outliers remained. To calculate waist-to-hip ratio, BMI adjusted, we iteratively dropped outliers for waist circumference and hip circumference separately, calculated the quotient of waist circumference and hip circumference, iteratively dropped outliers again, and residualized BMI on the quotient using simple linear regression. Post QC, we dropped all traits with African-ancestry sample size less than 18,500. We then quantile transformed each trait within ancestry and sex at birth stratum. Finally, to arrive at the list of 14 traits given in Table 1, we greedily selected traits based on European-ancestry heritability as estimated by Pan-UK Biobank such that all pairs of traits had absolute phenotypic Pearson correlation less than 0.2.

Next, we computed GWAS summary statistics using Plink2.0 for each trait-cohort combination including PC1-10 (projections onto PCs from 1000 Genomes and Human Genome Diversity Project samples), sex at birth, genomic data collection site, age (at time of phenotype measurement), age^2^, sex*age, and sex*age^2^. Plink2.0 was run using the –linear omit-ref, -- variance-standardize, --vif 1000 flags. For each EHR trait, we subselected individuals of European ancestry to match missingness levels in individuals of African ancestry.

### Real trait fine-mapping

We defined “hit variants” as variants with *p*<5*10^−6^ and cohort-specific MAF > 0.01 in Afr47k+Eur47k (using meta-analyzed summary statistics, including variants with MAF > 0.01 in either Afr47k or Eur47k), Afr47k, or Eur94k. We then defined 3Mb overlapping loci with 1Mb between start points, spanning the genome, starting at a genomic coordinate of 1, similar to Ref^12^. We fine-mapped each locus-trait pair that contained a hit variant in the central 1Mb, or that was located at the beginning of a chromosome and contained a hit variant in the first 1Mb. We excluded hit variants contained in 3 regions of long-range LD (chr6 27-34Mb, chr8 7-13Mb, chr11 45-58Mb), similar to Ref^12^, adjusted for different reference genome builds.

We fine-mapped each nominated locus-trait pair using 6 cohorts (with MultiSuSiE or SuSiE), and 4 methods (with Afr47k+Eur47k). All loci were fine-mapped with GWAS summary statistics and LD computed using LDStore2. To reduce computational cost, we did not exclude individuals with missing phenotypes while computing LD matrices, resulting in mismatch between GWAS and LD sample sets. Through simulation studies we determined this choice is unlikely to affect FDR or power (Supplementary Figure 21). All methods were run using the same parameters as described in *Simulation – fine-mapping*. Estimates of 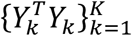, which are used by MultiSuSiE, were recomputed after residualizing out GWAS covariates. Variants fine-mapped by multiple loci were assigned PIP from the locus that contained the variant within the central 1Mb. If they did not fall within the central 1Mb of any locus, there were assigned the minimum PIP from all windows in which they were fine-mapped. To assess the statistical significance of the difference in the number of fine-mapped variants across pairs of methods or cohorts, we used a two-sided test on the z-score from a genomic block-jackknife with 200 blocks.

To assess functional enrichment of fine-mapped variants, we first selected 39 binary functional annotations from the baseline-LF v2.3 or baseline-LD v2.2 models^46–48^ excluding annotations derived from large genotyping or whole-genome sequencing datasets to avoid biasing functional enrichment in favor of European-ancestry cohorts. We then converted functional annotation BED interval files from hg19 to hg38 using LiftOver and dropped annotations that contained less than 1% of variants with MAF > 0.05 in Afr47k. Next, we calculated the mean functional enrichment of fine-mapped variants (*P*(*a*_*i*_ = 1|*PIP*_*i*_ > .5)/*P*(*a*_*i*_ = 1), where *a*_*i*_ equals 1 if variant *i* is in annotation *a* and 0 otherwise and *PIP*_*i*_ is the *PIP* of variant *i*) for all cohorts, annotations and, traits (taking the reciprocal for Repressed_Hoffman). We then greedily selected annotations to maximize the functional enrichment of fine-mapped variants averaged across Eur47k and Afr47k such that no pair of annotations had absolute Pearson correlation greater than 0.5 (calculated using variants with MAF > 0.05 in Afr47k). This process was repeated, greedily selecting annotations such that no pair of annotations had an absolute Pearson correlation greater than 0.2, yielding a set of 11 annotations, which were used for all functional enrichment analyses. When restricting to the top x variants with highest PIP, the functional enrichment of fine-mapped variants was calculated as *P*(*a*_*i*_ = 1*PIP*_*i*_ ≥ *PIP*_(*M*–*x*–1)_)/*P*(*a*_*i*_ = 1) where *PIP*_(*p*)_ is the *p*th smallest PIP for a cohort or method, *M* is the total number of variants, and *x* is the minimum number of variants with PIP > 0.5 across cohorts compared. To assess the statistical significance of the difference in functional enrichment of fine-mapped variants between a pair of methods or cohorts, *M*_1_ and *M*^2^, we used two-sided test on the z-score from a genomic block-jackknife with 200 blocks (each containing equal numbers of variants in Afr47k+Eur47k) on ∑_*a,t*_ *FE*_*a,t,M*1_ − ∑_*a,t*_ *FE*_*a,t,M*2_ where *FE*_*a,t,M*_ is the functional enrichment of fine-mapped variants for annotation *a*, trait *t*, and method or cohort *M*. To assess the statistical significance of the difference in functional enrichment of *χ*^2^ association statistics between cohorts, we used a two-sided paired t-test, treating trait-annotation pairs as independent.

## Code availability

A Python implementation of MultiSuSiE is available at https://github.com/jordanero/MultiSuSiE.

## Acknowledgements

We are grateful to Xihong Lin, Po-Ru Loh, Xilin Jiang, Arun Durvasula, Elizabeth Dorans, Katie Siewart-Rocks, Chaimaa Fadil, Gaspard Kerner, Buu Truong, Hui Li, and Phillip Nicol for helpful discussions. We thank All of Us participants for making this research possible. We also thank All of Us for providing access to the data used in this research. This research was conducted using the UK Biobank resource under application no. 16549 and funded by National Institutes of Health (NIH) grants R01 MH101244, R37 MH107649, R01 HG006399, U01 HG012009 and F31 HG013040. The funders had no role in study design, data collection and analysis, decision to publish or preparation of the manuscript.

